# Mapping climate change and health into the medical curriculum: co-development of a “planetary health – organ system map” for graduate medical education

**DOI:** 10.1101/2021.11.23.21265688

**Authors:** Hayden Burch, Benjamin Watson, Grace Simpson, Laura J. Beaton, Janie Maxwell, Ken Winkel

## Abstract

**Purpose:** Within the context of a review of a Doctor of Medicine graduate curriculum, medical students partnered with faculty staff to co-develop a novel curriculum resource exemplifying the integration of planetary determinants of health into existing medical curricula.

**Method:** We undertook qualitative methodologies involving a planetary health literature review and curriculum mapping exercise in three parts between April 2018 - May 2021. In part one, a student focus group sought students’ perceptions on opportunities for climate-change related teaching. Part two involved two 5-hour workshops that mapped planetary health principles to classical organ systems-based teaching areas. Part three consisted of curriculum mapping expert review.

**Results:** Participatory workshops involved 26 students and positioned students as leaders and partners in curriculum development alongside academics and clinicians. Final synthesis produced a comprehensive infographic rich document covering seven organ systems plus healthcare’s ecological footprint, the role of medical students and opportunities for applied skills and behaviours.

**Conclusions:** The student-staff co-production method adopted here promotes higher order relational and extended abstract reasoning by students, the ultimate task of any higher education. This approach, and the open access resource generated, provides an integrated and novel planetary health framework, supporting students to be leaders for a sustainable future.

**Practice Points:**

[1] This project provides a methodology to overcome barriers to curriculum-wide integration of planetary determinants of health and a template to move beyond stand-alone planetary health workshops or population health case studies.
[2] Student and educator co-development of planetary health teaching and learning resources promotes higher order relational and extended abstract reasoning by students, the ultimate task of any higher education.
[3] Integrating planetary health supports emerging clinicians in all areas of medicine to be leaders for a sustainable future.

## Introduction

In 1910, the publication of the Flexner Report, with its promotion of a standardised and scientific basis for the teaching of medicine, marked a revolution in medical education. Just over 100 years later the emergent, existential threat posed by human induced climate change, argues for another planetary scale transformation in medical training. Just as that report “*kept steadily in view the interests of two classes…the youths who are to study medicine and to become the future practitioners, and, secondly, the general public, which is to live and die under their ministrations*” (1), so too must this new, holistic framing address the same enduring constituents. Moreover, it must respond to the Oslerian critique of the Flexner approach (2) - “*teacher and student chased each other down the fascinating road of research, forgetful of those wider interests to which a hospital must minister*”. Thus, whilst the United Nations declares a “*climate emergency*”, Flexnerian-minded, reductionist science, research-preoccupied medical schools remain glacially slow at addressing what The Lancet has described as “*the biggest global health threat of the 21st century*” (3).

Nevertheless, in response to this bifocal challenge to both serve students and wider interests of the public, an international coalition of healthcare educators and students have begun to articulate a pedagogical response to the health impacts of the Anthropocene. Specifically, planetary health and sustainable healthcare frameworks, philosophies, consensus statements and early teaching experience have begun to be reported from Europe, North America and Australia (4-10). Whilst this nascent activity is promising, persistent barriers to the implementation of such urgently needed, systemically minded, curriculum initiatives are evident. These include lack of faculty leadership, lack of content expertise and its sequestration within global health and epidemiological subjects as well as limited or non-existent access to contextually appropriate educational resources (11, 12).

Consequently, the practical implementation of planetary health concepts into predominantly biomedical curricula is frequently piecemeal, unidirectional and often poorly relates to clinical practice (13, 14). Indeed, students, who often have difficulty connecting the bidirectional factors affecting the upstream systemic forces with downstream patient trajectories, are ill-equipped to apply this knowledge in their professional practice and may not appreciate the need for advocacy.

At the University of Melbourne Medical School in Australia, students interested in environmental health identified the review of a Doctor of Medicine (MD) graduate curriculum as an opportunity to address the ‘planetary health’ gap in their teaching. The students partnered with Medicine, Dentistry and Health Sciences faculty staff to map opportunities and an evidence-base for integrating teaching of the planetary determinants of health within the classical ‘organ-systems’ curriculum framework.

The primary objectives were to generate a curriculum map that (i) aligns the first-year curriculum more closely with existing Australian Medical Council’s (AMC) Graduate Student Attributes, (ii) exemplifies the integration of planetary health as a cross-cutting theme into an existing curriculum, with particular focus on the mechanistic impacts of climate change on patients and practice; and (iii) specifies opportunities to integrate principles of sustainable healthcare. This strategy, and output, described and available here, extends a familiar ‘organ-system’ reductionist curriculum structure into a post-Flexnerian student-staff co-developed planetary health framework.

## Methods

We conducted a planetary health literature review and curriculum mapping exercise between April 2018 - May 2021. The mapping project was conducted in three parts. Our mixed methodology is outlined in **Table 1**.

**Table 1.**
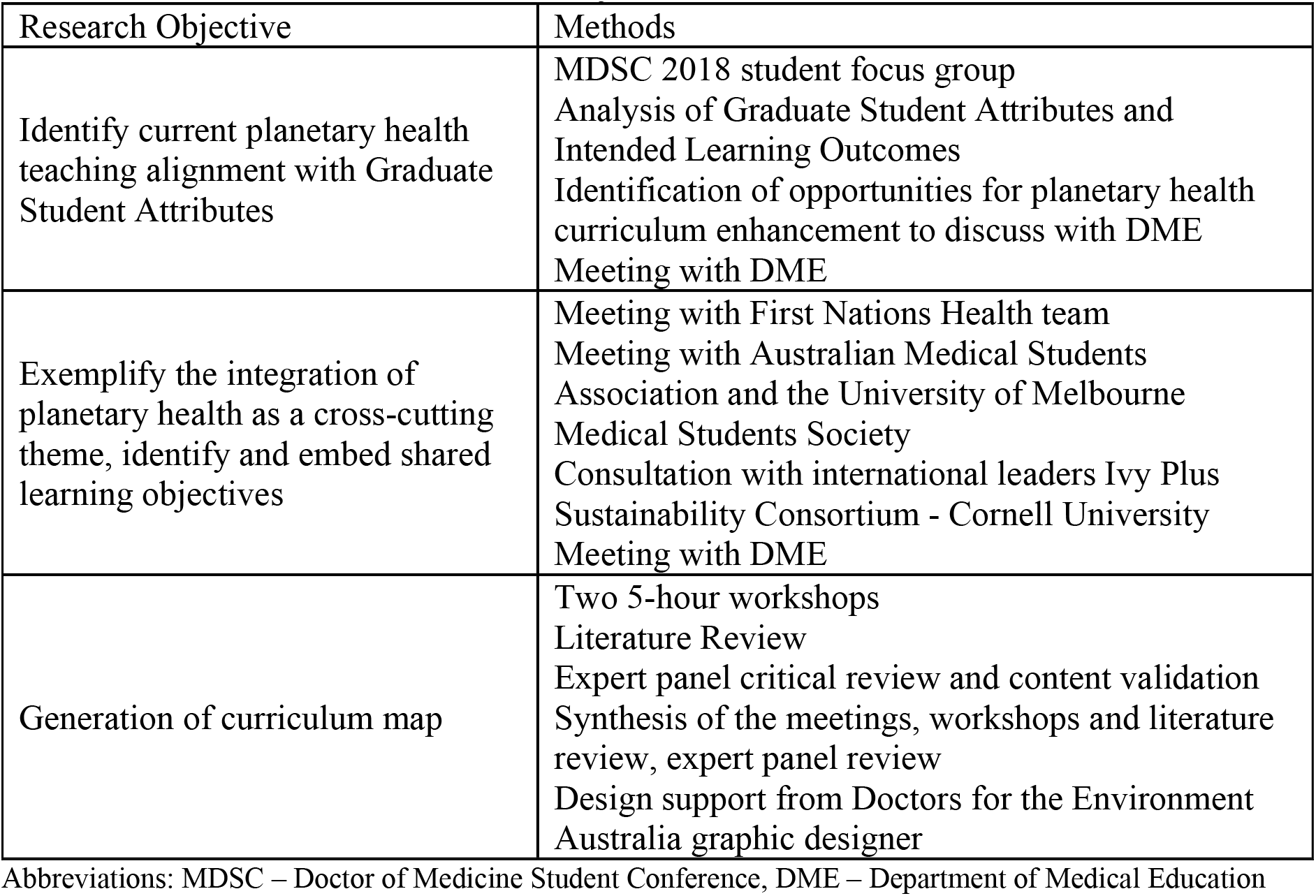
Methods used to meet research objectives

| Research Objective | Methods |
| --- | --- |
| Identify current planetary health teaching alignment with Graduate Student Attributes | MDSC 2018 student focus group<br>Analysis of Graduate Student Attributes and Intended Learning Outcomes<br>Identification of opportunities for planetary health curriculum enhancement to discuss with DME<br>Meeting with DME |
| Exemplify the integration of planetary health as a cross-cutting theme, identify and embed shared learning objectives | Meeting with First Nations Health team<br>Meeting with Australian Medical Students Association and the University of Melbourne Medical Students Society<br>Consultation with international leaders Ivy Plus Sustainability Consortium - Cornell University<br>Meeting with DME |
| Generation of curriculum map | Two 5-hour workshops<br>Literature Review<br>Expert panel critical review and content validation<br>Synthesis of the meetings, workshops and literature review, expert panel review<br>Design support from Doctors for the Environment Australia graphic designer |

Abbreviations: MDSC – Doctor of Medicine Student Conference, DME – Department of Medical Education

In part one, a student focus group at the University of Melbourne MD Student Conference (MDSC 2018) sought first year (MD1) to final year (MD4) students’ perceptions on opportunities for climate change related health teaching in the Melbourne Medical School (MMS) curriculum (unpublished data). Eleven students representing all four years of the MD attended. Following presentation of focus group data to the Department of Medical Education (DME), we formed the ‘Planetary Health Curriculum Taskforce’ to further map the intersections between the University of Melbourne MD1 curriculum and planetary health. The Curriculum Taskforce consisted of three medical students and two medical faculty academics from the University of Melbourne.

To situate our mapping outcomes in their practical application at MMS, and, to exemplify the cross-cutting opportunities of planetary health and embed shared learning objectives, we held meetings with key stakeholder group representatives prior to and throughout this period. For example, engagement with the First Nations Health team inspired application of the Social Determinants of Health model and identification of overlap for First Nations and planetary health MD1 learning outcomes. We also met with the MMS DME to discuss the applicability of our mapping, and, corresponded with international leaders from Cornell University as a member of the Ivy Plus Sustainability Consortium (made up of sustainability officers from the Ivy League universities) (15).

Part two involved two 5-hour mapping workshops conducted in May and June 2019. Each workshop was facilitated by the Curriculum Taskforce. Workshop objectives were to i) conduct a literature search on the health impacts of climate change and ii) generate a spreadsheet that systematically mapped literature search content. To achieve these objectives, we applied three pedagogical models:

i. *Model 1:* Organ systems (e.g. cardiovascular system)
ii. *Model 2:* Social Determinants of Health (16)
iii. *Model 3:* Major health consequences of climate change (World Health Organization) (17)

We integrated across literature search content using Model 1 headings (e.g. cardiovascular system) with Model 2 columns (relevant social determinants of health) and Model 3 rows (relevant major consequences of climate change). As an example, literature relating to exacerbations of acute heart failure (Model 1 - cardiovascular system) was tabulated according to relationships with social determinant factors such as impeded access to healthcare and patient age (Model 2) in the context of increasing bushfires, worsening heatwaves and air pollution levels (Model 3).

Principles of sustainable healthcare, such as promoting increased awareness and understanding of health co-benefits of emission reductions (e.g. low carbon transport and dietary choices that decrease morbidity and mortality through reduced air pollution levels, increased physical activity and plant-based diets) (18) were also included in alignment with the existing MMS Intended Learning Outcome: to *understand the principles of practising medicine in an environmentally responsible way* (19). We also mapped healthcare’s ecological footprint, the role of medical students and opportunities for applied skills and behaviours.

Workshop participants were recruited via the closed Facebook pages *University of Melbourne Doctors for the Environment* and *University of Melbourne Medical School Common Room*. An additional five participants were contacted directly via Facebook Messenger. Given student perceptions and experience likely differ across stages of their education (20), a broad cohort was purposefully sampled. We created an A4 electronic poster outlining workshop descriptions, date, time, location and a link to register. A $50 cash prize was offered as an incentive and advertised as randomly awarded to a participant who attended both workshops. All participants registered using Google Forms and provided consent for publication. Formal ethics approval was not sought prior to initiation of this project. Twenty-six medical students from the MMS, and those intercalating to pursue the Master of Public Health (MPH) as a combined MD-MPH at the School of Population and Global Health, voluntarily registered to participate in either one or both workshops (**Table 2**).

**Table 2.** Characteristics of part two literature review and curriculum mapping workshop participants

| MD year level | MD1 | MD2 | MD3 | MD/MPH | MD4 |
| --- | --- | --- | --- | --- | --- |
| **Number of participants** | 6 | 6 | 7 | 2 | 5 |
| **Age range (yrs)** | 21 - 22 | 22 - 24 | 23 - 29 | 26 - 31 | 23 - 31 |
| **M:F ratio** | 1:5 | 1:5 | 4:3 | 2:0 | 3:2 |

Preceding the workshops, the Curriculum Taskforce conducted a background literature review searching the Medline database for keywords ‘planetary health’, ‘climate change’ and ‘health’ to generate a group of core articles and associated references. Six key articles were selected (21-26) based on journal, citations, relevant content, and Curriculum Taskforce consensus regarding the relevance and seminal nature of the publication.

The student-leader presented a 30-minute workshop introduction to outline the workshop structure and objectives on both days. Workshop participants were divided into small groups of 3-4 and allocated an organ system. Each group conducted a content analysis of the 6 core articles with a focus on the relevance to their designated organ-system (Model 1). Further research articles were subsequently obtained through reference snowballing (27) and searching the Medline database for climate change and organ system-specific search terms. Participants were rotated within groups once per session and each group rotated organ-systems three times per workshop to provide students with an opportunity to contribute to different organ systems and collaborate with different participants. Rotation also functioned as a peer-review process with an initial 30 minutes assigned to review and critique of the previous group’s work. Facilitators roved between all groups.

Part three consisted of curriculum mapping review and editing from October – November 2019. Seven clinicians with teaching and research experience in climate change and health were voluntarily recruited from the Doctors for the Environment Australia Victorian Committee (**Table 3**). A mixture of specialist and generalist clinicians were recruited to ensure broad clinical experience. Each clinician was allocated two organ system blocks and instructed to review and edit content for clinical relevance, strength of evidence and quality of writing.

## Results

We developed an evidence-informed planetary health – organ system map (the complete *Planetary Health – Organ System Map* is available as **Supplemental Digital Appendix 1**), with planetary health as a cross-cutting theme, to exemplify the integration of planetary health learning outcomes for first year MD students into the existing organ systems-based model of teaching. Through engagement with stakeholders, we were able to integrate additional common learning outcomes, particularly related to First Nations or Aboriginal and Torres Strait Islander health.

Our initial mapping workshops generated a shared online Microsoft Excel spreadsheet. We completed mapping for 7 of the 11 organ system blocks: cardiovascular, respiratory, renal, gastrointestinal, neuroscience, reproduction and intersystem. The remaining blocks (endocrine, metabolism, locomotor and exercise) were deemed scarce in peer-reviewed content in addition to constraints on project time and human resources.

We then synthesised and edited workshop content into paragraph format with organ system chapters (Model 1), consequences of climate change as chapter sections (Model 3) and determinants of health guiding paragraph structure (Model 2). The first report version of the mapping was available at the end of August 2019 including a summary of relevant Graduate Student Attributes and Intended Learning Outcomes at the conclusion of the mapping report. The final synthesis, inclusion of images, figures and executive summaries was completed by Curriculum Taskforce student members with design support from Doctors for the Environment Australia. The final mapping report was completed in May 2021.

## Discussion

There is growing international medical recognition, regulatory obligation and student-led demand that planetary health concepts, including the health impacts of climate change and the principles of sustainable healthcare, are embedded into medical education and practice (7, 10, 12, 28-30). In Australia, the Medical Deans of Australia and New Zealand established a Climate Change and Health Working Group which has developed graduate outcome statements and learning objectives that incorporate the principles of planetary health (31, 32).

Internationally, the curriculum principles that are emerging to address this challenge include alignment with global health priorities that address the socio-economic and environmental determinants of health under the UN Sustainable Development Goals framework, indigenous eco-health-centric leadership, promotion of systems thinking and change, active student-staff collaboration, and experiential, practice-based learning that cultivates interprofessional teamwork, advocacy and leadership development (4-11). Research from the Centre for Environmental Policy at Imperial College London specified three key strategies for successful integration: i) integrate sustainable healthcare as a cross-cutting core theme, ii) facilitate educators and students sharing knowledge and teaching one another in this emerging field, and iii) relating teaching to clinical practice (12).

One of the intermediate steps, towards broader integration of climate change and sustainability content into medical education, is to link existing classical organ-systems focused preclinical studies with the protean impacts of climate change on the bodies of individual patients. A 2019 New England Journal of Medicine interactive perspective provides a useful open access resource in that direction (33). More recently, Emory University students and staff reported their experience in the incorporation of climate content into the pre-clinical, pathophysiology centred medical school curriculum (34). Whilst their report includes some examples of climate change-organ-system linkages and learning objectives, it is neither systematic in process nor pedagogically structured under one or other of the preceding international frameworks.

Our work, in seeking to align curricula with AMC Graduate Student Attributes has applied the emergent pedagogic principles to jointly produce an integrated planetary health and organ system curriculum map for first year teaching at the MMS. In doing so, we demonstrated the potential opportunities for traditional biomedical curricula to integrate themes relating to planetary health across multiple organ-systems and learning objectives. This approach aligns with recent understanding that planetary health is a theme akin to ethics or leadership that should spiral through the core curriculum (12). In this context our mapping of health strategies that mitigate greenhouse gasses and produce co-benefits to health may suit integration in subsequent year-levels in the context of leadership or ethics discussion.

Central to our approach was the application of the three selected models, which generated mapping output that identified and explored the bidirectional relationships between broad public and planetary health content, and person-centred biophysiological mechanisms of disease. This approach was enhanced by defining our scope to focus on clinical relevance.

Although sustainable healthcare education has been included in medical curricula at some schools in the United Kingdom for over a decade (9), this project provides a methodology to overcome barriers to curriculum-wide integration and provide a template to move beyond stand-alone planetary health workshops or population health case studies. As one example, there is opportunity to teach students about the biophysiological mechanisms of thromboembolism formation - in the context of learning about heatwave events and increased 2.5 particulate matter air pollution (PM_2.5_) impacting on dehydration and coagulation pathways. Further studies are likely warranted to assess the extent to which integrated knowledge might be understood, recognised and applied in their future clinical practice.

A key challenge in developing our multi-model mapping was addressing how clinically relevant literature best aligned with each model. This was highlighted in mapping areas that overlapped i.e., literature regarding dehydration pathways in pregnant or fertile-aged women may have been addressed within either the cardiovascular or reproductive blocks. Likewise, the sequalae of dehydration may also be explored in chapter sections regarding heatwaves / ozone and aeroallergen levels or vulnerable shelter and human settlements / migration. This overlap may present an opportunity for consolidation of knowledge across the academic year.

Importantly, these points of uncertainty created discussion that enhanced collaborative teaching and learning between students, faculty academics and expert clinicians. Empowering students to be leaders and partners in teaching and developing education material is known to improve their own effectiveness as learners, and confidence in teaching (35). It also explicitly promotes higher order relational and extended abstract reasoning by students (Blooms’ revised taxonomy Levels 4-6), the ultimate task of any curriculum (36).

Simultaneously, given the emergent nature of planetary health pedagogy our research methodology and output may assist medical educators in addressing known broader challenges to integrating sustainable and planetary healthcare teaching (12). For example, as this methodology enables students to share their knowledge and experiences with teachers outside of formal classroom boundaries, it may support the evident lack of teachers proficient in sustainable and planetary healthcare teaching within medical faculties. Alternatively, our mapping output might address the need for learning resources that are centralised, accessible and frequently peer reviewed. A remaining challenge, given the assessment driven nature of medical education, is the development of ‘assessment for learning’ inclusive of planetary health teaching within medical education. We have not completed a review of the assessment blueprint. However, we propose this as a valuable exercise for internal medical education assessment teams, as assessment often strongly correlates with students’ prioritisation of content (37). Moreover, we would suspect that team and portfolio-based Work-Integrated Learning assessments would enhance both student engagement and contextually appropriate learning (constructive alignment) (38).

Our planetary health curriculum map was developed specifically by University of Melbourne students with experience and remit to maintain relevance to their MD program. It is likely that other local and international medical schools apply similar organ-system frameworks of teaching and learning that would benefit from this body of work. Participants in the Curriculum Taskforce were enthusiasts for planetary health education, and this may have influenced the mapping themes and content that were generated. Future work may be strengthened by including students and faculty that may prioritise planetary health differently in the context of a busy curriculum.

Ultimately the role of medical professionals in providing leadership, advocating for sustainable healthcare and adopting evidence-based strategies for management of planetary health related risks to health and healthcare infrastructure, operations and personnel, has been widely acknowledged by professional and educational bodies (8, 39-42). This planetary health curriculum map, and the methodology employed, presents an opportunity for leaders in medical education who are seeking a model of integration with existing curricula. This approach thus extends a familiar ‘organ-system’ reductionist curriculum structure into a post-Flexnerian student-staff co-developed planetary health framework, supporting students to be leaders for a sustainable future.

## Supporting information

Supplemental Digital Appendix 1

## Data Availability

All data produced in the present study are available upon reasonable request to the authors

## Acknowledgments

The authors wish to thank members of Doctors for the Environment Australia (DEA) for their clinical and research expertise and graphic support to produce our final curriculum mapping resource.

## Disclaimers

None.

## Previous presentations

None.

