## Supplemental Digital Appendix 1 for "Mapping climate change and health into the medical curriculum: co-development of a “planetary health – organ system map” for graduate medical education"

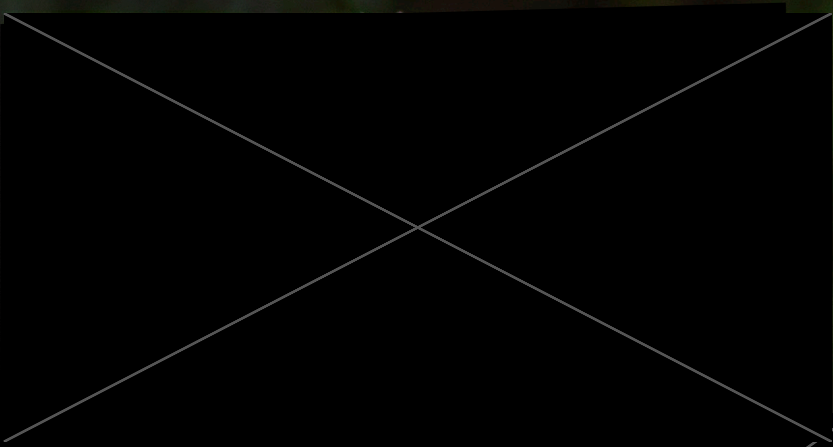

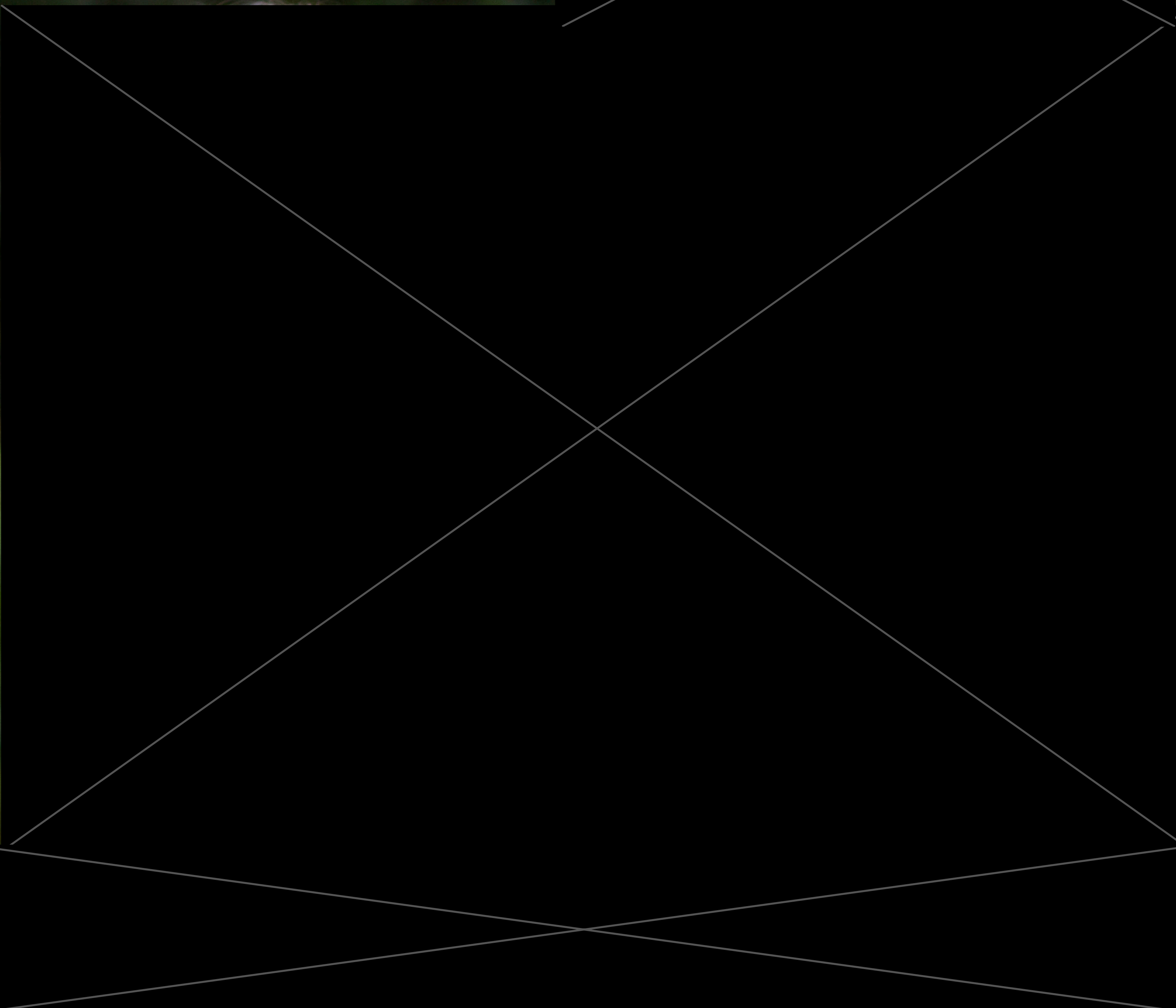

© Doctors for the Environment Australia Inc.

This work is copyright Doctors for the Environment Australia Inc. All material contained in this work is copyright Doctors for the Environment Australia Inc. except where a third party source is indicated. Permission to use third party copyright content in this publication can be sought from the relevant third party copyright owner/s.

This work is licensed under a Creative Commons Attribution 4.0 International License. To view a copy of this license visit http://creativecommons.org.au/.

You are free to distribute, remix, adapt, and build upon the material in any medium or format, particularly for educational and non-profit services, so long as you attribute Doctors for the Environment Australia and the authors.

Citation: Burch H, Watson B, Simpson G, Beaton L. J, Maxwell J, Winkel K. Mapping climate change and health into the medical curriculum: co-development of a "planetary health-organ system map" for graduate medical education. Melbourne, Australia: Doctors for the Environment Australia; 2021.

Images, unless specified otherwise, are copyright of Shutterstock (www.shutterstock.com).

Cover photo by Jesse Thompson / Doctors for the Environment Australia

Respiratory diseases are impacted by climate change, metered dose inhalers contribute towards healthcare's greenhouse gas emissions and depending on the clinical situation alternatives can be considered.

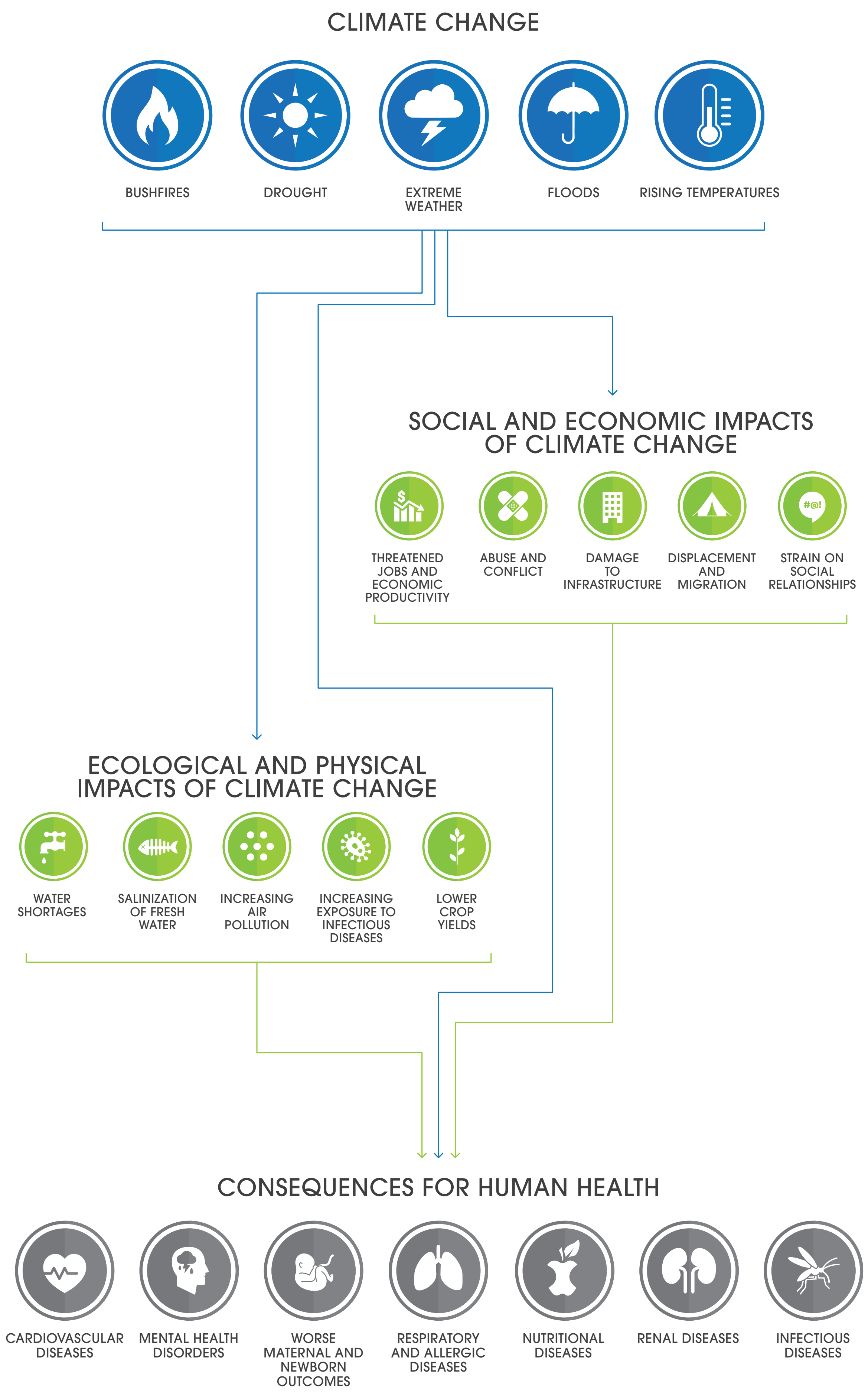




### Climate Change Poses the Greatest Threat to Human Health

Human civilisation has flourished in pursuing health, economic and development gains over recent centuries. However, these achievements in human activity have been made at a cost—the ongoing health of human civilisation and the state of the natural systems on which it depends.[1]

The exploitation of nature's resources, degradation of life support systems and increasing global pollution all undermine the health of the planet and its life forms.[1, 2] Ecosystems and biodiversity are showing rapid decline as human actions threaten more species with global extinction than ever before.[3] Climate change, as one critical domain of planetary health, is a major health emergency[4-6] undermining the last 50 years of gains in development and global health.[7] The consequences threaten every dimension of human health and are catastrophic to societies' long-term survival.

Valuing planetary health is central for the medical profession, who have a responsibility to protect and advance the health and wellbeing of individuals, communities and populations.

Paradoxically, healthcare itself, which exists to protect health, is a large contributor to ecological decline.[8-11] In order to provide high quality care and improved health—without exhausting natural resources or causing severe ecological damage[12]—understanding of how healthcare delivery impacts on planetary health is important.

Diagram (opposite page) authors own based on Hughes and McMichael 2011 and Whitmee et al. 2015.

### Overview

Climate change is a health emergency posing significant threats to health and to the healthcare sector.

As a result, there is growing demand for planetary health concepts, including the health impacts of climate change and the principles of sustainable healthcare, to be embedded systematically into medical education and practice.

Student members of Doctors for the Environment Australia (DEA) undertook a review of their Doctor of Medicine (MD) graduate curriculum. Their objective was to align their education more closely with the existing Australian Medical Council graduate outcome statements (within resource below).

The students partnered with staff across several departments and two schools within the University of Melbourne Medicine, Dentistry and Health Sciences faculty to form the volunteer “Planetary Health Curriculum Taskforce”.

The Taskforce developed ‘Mapping Climate Change and Health into the Medical Curriculum’ as a resource for all Australian medical educators, students and clinicians.

The resource exemplifies how planetary health concepts and knowledge can be integrated into the organ systems framework for medical education. This framework, which compartmentalises content according to the body’s organ systems, is familiar to all Australian medical students and teachers.

Mapping Climate Change and Health into the Medical Curriculum demonstrates how planetary health is a cross cutting theme relevant to every specialty. It may be used as a guide for curriculum development, whereby planetary health can be integrated into existing organ systems-based teaching. Educators can easily find relevant planetary health learning for lectures, tutorials, case-based discussions and clinical scenarios.

With the frequency and severity of extreme weather events predicted to continue to worsen, doctors, whether in training or in practice, must be equipped with the knowledge, skills, values, competence and confidence they need to sustainably assess, manage and treat patients presenting with climate change related illnesses.

This resource supports the development of well-rounded medical students and doctors equipped to practice medicine now and in the future.

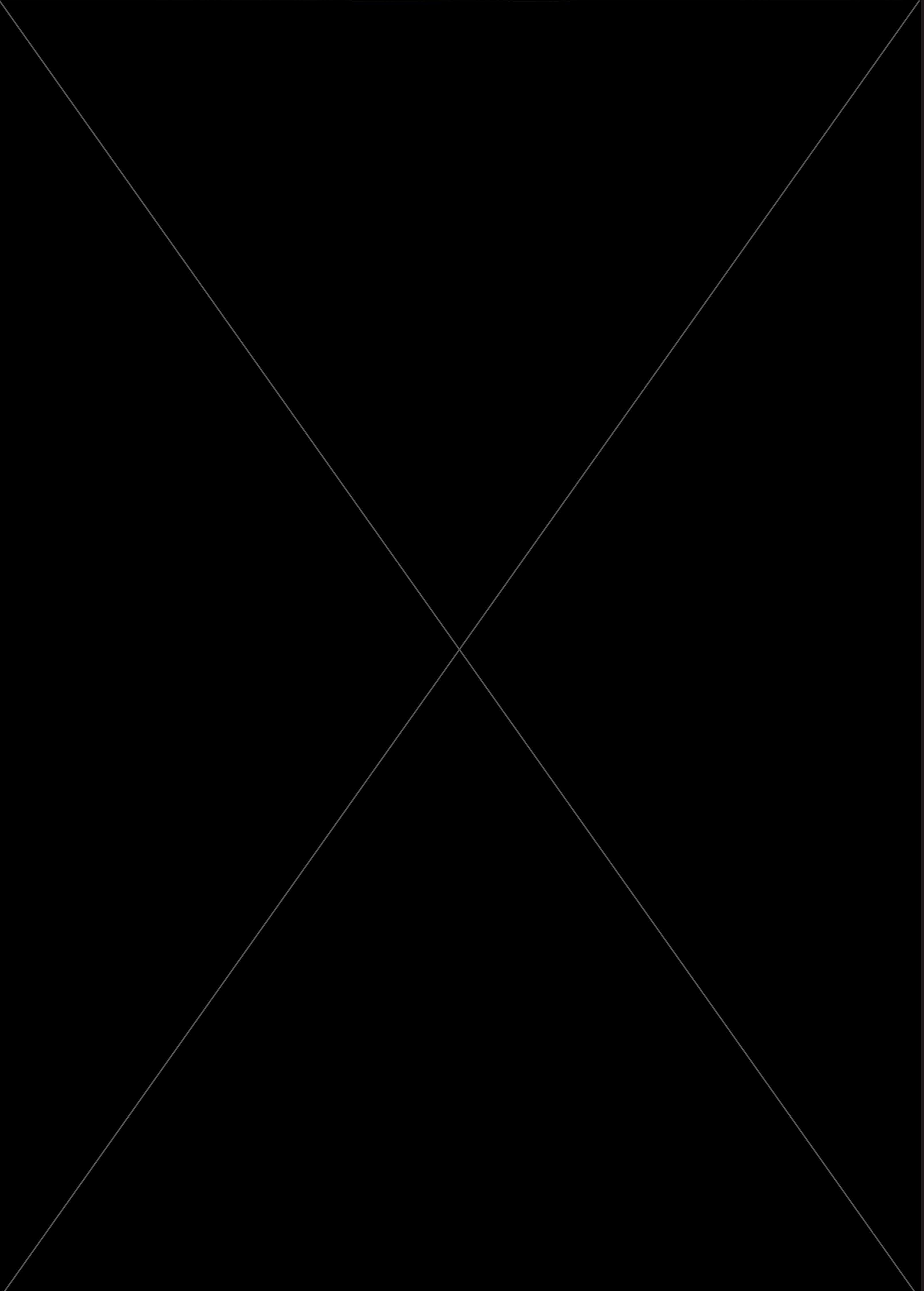

### Table of Contents


1 **Executive Summary**

5 **Curriculum Mapping Framework and Methodology**

9 **Organ System Mapping**

9 Cardiovascular System

19 Respiratory System

29 Renal System

35 Gastrointestinal System

47 Neuroscience

55 Reproduction

63 Intersystem

65 **Healthcare's Ecological Footprint**

69 **The Role of Medical Students**

69 MD1 PCP Final Learning Outcomes

69 MD2 Graduate Student Attributes Developed

73 **Opportunities for Applied Skills and Behaviours**

75 **References**

87 **Appendix**


Image source: Jesse Thompson /
Doctors for the Environment Australia

### Executive Summary

The health effects of climate change are already being felt and are projected to become worse if greenhouse gas emissions continue. These impacts will affect human health directly, indirectly, and through societal responses.

1. Direct health impacts: heatwaves, bushfires and smoke, storms, floods and drought may lead to acute cardiorespiratory events such as acute coronary syndromes and storm asthma, and psychological conditions such as post-traumatic stress disorder.

2. Indirect health impacts: the decline in air quality, food and water quality and quantity, and change to ecosystems contribute to worsening of disease as a result of financial consequences, poor access to food, water, healthcare, and exacerbation of psychological conditions like anxiety and depression.

Unchecked climate change will continue to increase the frequency and severity of extreme weather events across Australia. During the 2019/2020 Australian Black Summer bushfires, the bushfire smoke alone was estimated to be responsible for 417 excess deaths, 3151 additional hospital admissions for cardiovascular and respiratory diseases and 1305 asthma presentations to emergency departments.

Overall, climate change will increase inequality, as people with fewer material, social and health resources will be more vulnerable to the adverse impacts of climate change. Vulnerability to climate change ultimately depends on geographic, social, economic, cultural and biological factors.

Groups most at risk include:

- Socio-economically disadvantaged
- People with disabilities and chronic disease
- Remote Aboriginal and Torres Strait Islander communities
- Pregnant women and their unborn children
- Children
- Older people
- Rural communities
- Industries and their workers

Indigenous Australians are disproportionately more vulnerable to the health impacts of climate change. The capacity of Indigenous Australians to respond is undermined by the ongoing dispossession and loss of access to traditional lands, waters and natural resources. These factors can be compounded by challenges of poverty, prevalence of chronic disease and intergenerational disadvantage.[13] This leads to significant implications for the level and types of services needed to address the health burden experienced by Indigenous peoples due to climate change. Conversely, understanding of Caring for Country is a central planetary health concept and is linked to a broad range of health and environmental benefits. Indigenous Australians should be acknowledged in their capacity to provide expert, time-tested ways of knowing that allow all Australians to engage and reflect on the relationship of planetary and human health.

On current emissions trajectories, Australian doctors should expect to treat a threefold increase in heatwave-related deaths in Melbourne and Brisbane and fivefold in Sydney over the period 2013 to 2080, compared with current heat-related mortality.[14]

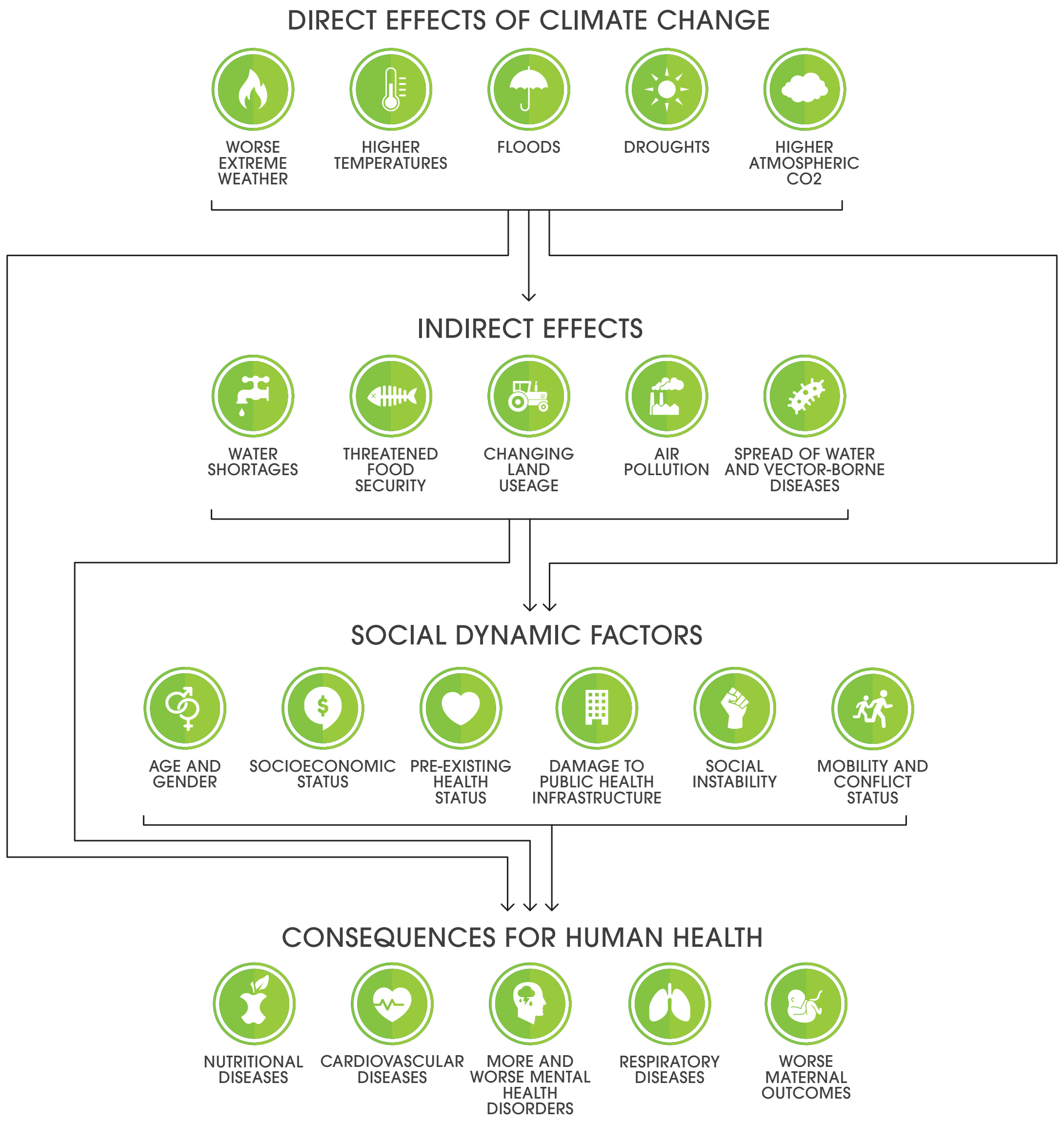


Authors own based on Whitmee et al. 2015 and Watts et al. 2015.

### Climate Drivers and the Health Impacts for Australians

| CLIMATE DRIVER | HEALTH IMPACTS |
|---|---|
| **Extreme weather events** – more severe and frequent bushfires, floods and storms. | Injuries and deaths from heat, smoke inhalation, burns, drowning in flooding, infectious disease transmission through floodwater, exposure to pollutants. Trauma and mental health impacts (distress, grief, behavioural health disorders, acute and chronic anxiety or depression). |
| **Extreme heat** – hotter days, more frequent, severe, prolonged heatwave events. | Heat-related mortality and morbidity, increased exacerbations of pre-existing cardiovascular, respiratory, renal and psychiatric conditions, especially in vulnerable populations such as elderly, children, mentally unwell and those with existing chronic diseases. |
| **Outdoor air quality** – worsening particulate matter, ozone and aeroallergens (pollens). | Respiratory tract infections and exacerbation of chronic diseases (e.g. ischaemic heart disease, chronic obstructive pulmonary disease and asthma). |
| **Water security** – drought, bushfires and flooding, increasing entry of contaminating debris, human/animal waste into waterways and drinking water supplies. | Increasing rural and regional mental health consequences including suicides, particularly amongst agricultural producers experiencing drought.Increased water-borne diarrheal and intestinal diseases (e.g. campylobacter, cryptosporidiosis), wound and blood stream infections. |
| **Food security** – drought, altered crop yields and altered land availability, higher temperatures favouring pathogen proliferation. | Increasing food-borne illness and diarrhoeal diseases (e.g. salmonella, campylobacter, cholera, harmful algal blooms). Malnutrition and increasing risk of high-energy, low-nutrient diets predisposing to metabolic syndrome. |
| **Change to vector-borne ecology** – increasing proliferation, increasing biting activity, increasing latitude and altitude habitat. | Increasing exposure and infections with arboviruses and other vector disease such as: dengue, Ross River fever, Murray River encephalitis, Barmah Forrest virus, Mycobacterium ulcerans. |
| **Sea level rise, ocean temperature and acidification changes** – migration and loss of fish stock, forced migration from low-lying nations and competition for dwindling resources. | Worsening conflict, mental health issues, health-related problems of climate induced displacement. Loss of heritage and cultural lands (e.g. Torres Strait Islands). Food and water insecurity for Australians. |

Climate change is affecting the health of Australians, now and in the future. These health consequences are from multiple threats, such as increasingly severe bushfires and floods impacting on cardiovascular, respiratory and psychological diseases, to sea level rise and ocean acidification risking food security and loss of cultural lands and identity. For a more comprehensive look at how greenhouse gas emissions affect the determinants of health and specific disease outcomes, see organ block chapters. Sources: (14-18)





### Health Co-benefits of Greenhouse Gas Mitigation

| ACTION | HEALTH AND POPULATION BENEFITS | ENVIRONMENTAL BENEFITS |
|---|---|---|
| **Active transportation.** More physical activity i.e. walking or cycling and reducing vehicle usage. | Reduced risk for major non-communicable diseases - obesity, ischaemic heart disease, stroke, diabetes, and renal disease. Positive effects on mental health, improved symptoms of stress, anxiety, and depression.<br><br>Decreased inhalation of air pollutants - improved asthma symptoms; reduces rates of lung cancer, renal disease, heart disease and stroke.<br><br>Decreased health expenditure and workdays lost due to illness. | Decreased greenhouse gas and fine particulate emissions from vehicles.<br><br>Reduction of vehicle kilometres travelled also reduces the concentration of ground level ozone precursors. |
| **Plant-based dietary modifications** | Diets rich in plant-based foods, such as fruit, vegetables, and unprocessed cereals, reduce the risk of various gastrointestinal and other cancers.<br><br>Reduced animal products (particularly red meats) decrease the risk of ischaemic heart disease and colorectal cancer. Reduced saturated fat intake decreases the risk of obesity, diabetes and heart disease.<br><br>A balanced reduction in protein intake may slow chronic kidney disease progression in patients with pre-existing kidney disease. | Plant-based foods have significantly lower emissions and water consumption per gram of protein than animal products. |
| **Improved understanding of Caring for Country** | Can promote more frequent exercise, lower rates of obesity, diabetes, renal disease and cardiovascular disease, and less psychological stress. | The harvesting and cultivation of bush foods have a significantly lower carbon footprint than contemporary agricultural approaches - in part due to being native to the Australian climate. |
| **Increased green spaces and urban forestry** | Increased exposure to natural and green space settings, particularly in urban areas has shown multiple benefits for mental health and neurological functioning.<br><br>Increased absorption of pollutants by trees along busy roads decreases inhalation of air pollutants and improves asthma symptoms. | Improved air quality by decreasing levels of pollutants. |

Co-benefits describe the actions and behaviours that are mutually beneficial to the climate, individuals and populations. Understanding how these actions contribute toward multiple health benefits is crucial for doctors who seek to practice medicine in an environmentally responsible way. For a more comprehensive look at the individual and population co-health benefits of greenhouse gas mitigation, see chapter blocks. Sources: (19-38)

### Curriculum Mapping Framework and Methodology

The project objectives were to generate a planetary health organ system curriculum map that:

1. Aligns first year organ-system based curricula with existing Australian Medical Council (AMC) graduate outcome statements;[39]

2. Exemplifies the integration of planetary health as a cross-cutting theme into an existing systems-based curriculum, with particular focus on the mechanistic health impacts of climate change that relate teaching to clinical practice; and

3. Specifies opportunities to integrate principles of sustainable healthcare.

The project was conducted in three parts.

In part one, a student focus group at the Melbourne Medical Doctorate Student Conference (MDSC 2017) sought first-to-final year students' perceptions on opportunities for climate change related health teaching in the Melbourne Medical School curriculum (Appendix I). Following the workshop, engaged students and staff formed the Planetary Health Curriculum Taskforce. Over the course of eighteen months the Curriculum Taskforce engaged the Department of Medical Education, faculty First Nations health team and international leaders in planetary health teaching from the Ivy Plus Sustainability Consortium (made up of sustainability officers from the Ivy League universities).

Part two consisted of two single-day workshops conducted in May and June 2019 (Appendix II). Twenty-six medical students from the University of Melbourne were voluntarily recruited by the Taskforce student leaders to complete a literature search on the health impacts of climate change. Three distinct models were applied (outlined below). The Medline database reputable for medical research was used as the primary search engine, using keywords relating to climate change and specific organs. Further studies were found by reference snowballing (the review of article reference lists).





Workshop content was then synthesised by the Curriculum Taskforce.

We completed mapping for seven of the eleven organ system blocks: cardiovascular, respiratory, renal, gastrointestinal, neuroscience, reproduction and intersystem. The remaining blocks (endocrine, metabolism, locomotor and exercise) were deemed scarce in peer-reviewed content in addition to constraints on project time and human resources.

Part three consisted of curriculum mapping review and editing from October – November 2019. Seven clinicians were recruited voluntarily from the Doctors for the Environment Australia Victorian Committee (Appendix III). All participants were clinicians with teaching and research experience in climate change and health and included a mixture of specialist and generalist clinicians to ensure broad clinical experience. Each clinician was allocated two organ system content blocks for review.

Final synthesis, development of key findings and editorial additions, such as images and figures, were made by the student leader of the Curriculum Taskforce and sourced throughout the mapping process.

The mapping process was completed using the following three models.

#### Model 1: Organ Systems

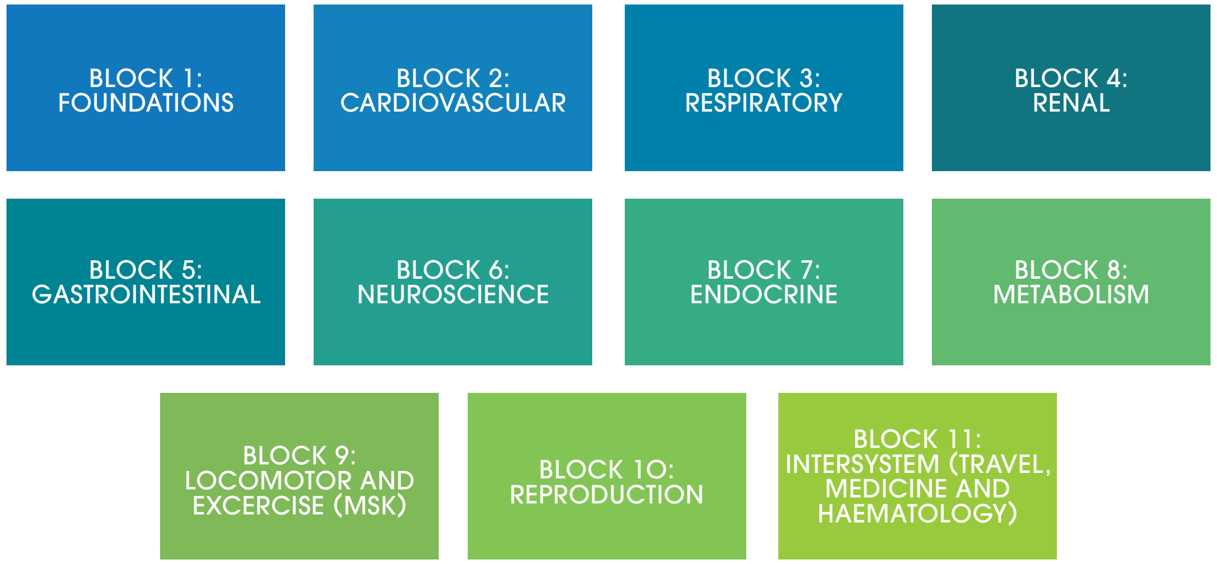


The University of Melbourne Medical School teaches the first year of the MD course using 11 teaching blocks, based primarily on organ-systems. This framework is familiar to most medical providers. Literature search findings relating to climate change and health were initially grouped according to each organ system. MSK = musculoskeletal.

### Model 2: Social Determinants of Health

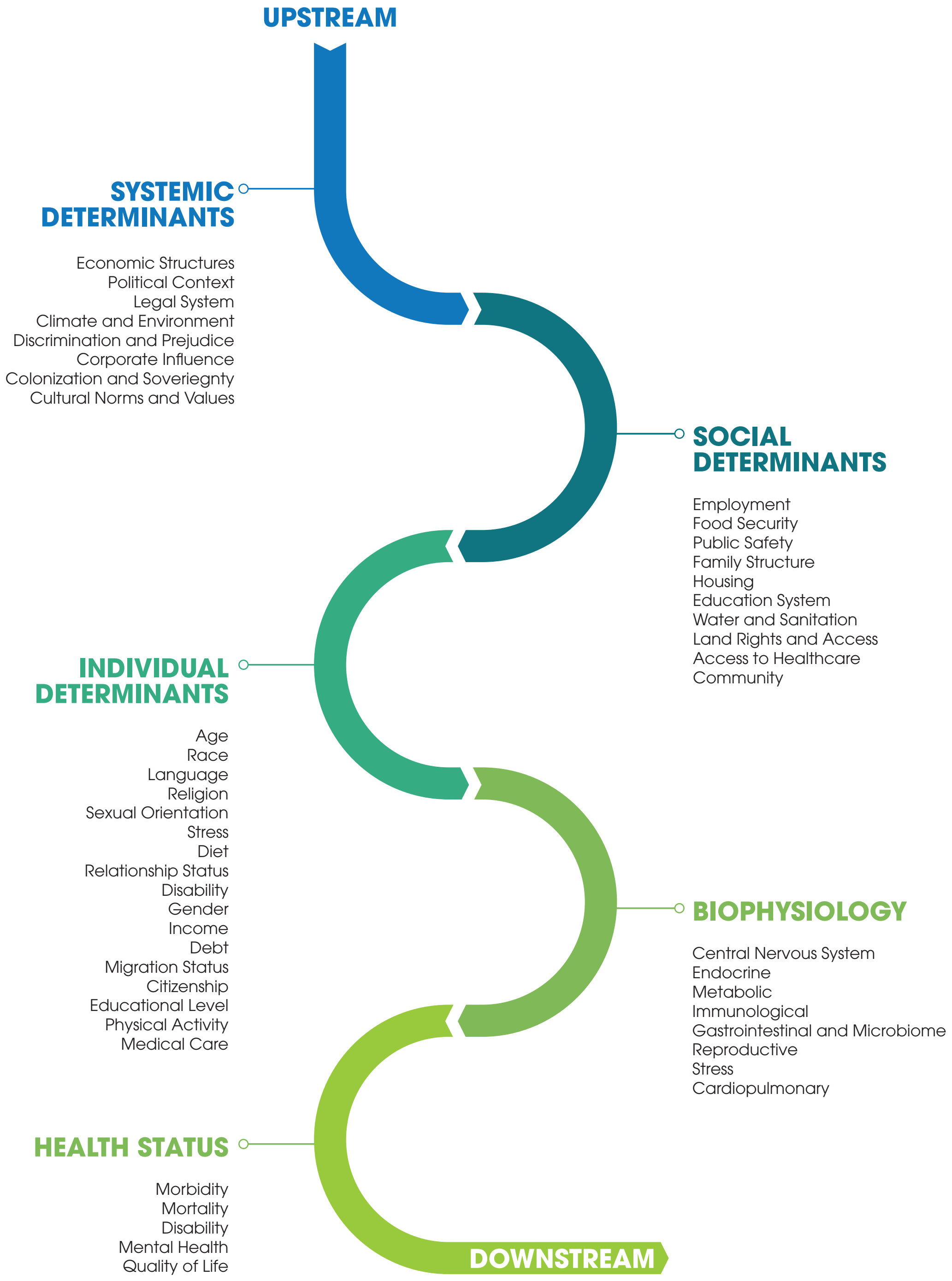


The widely used Determinants of Health framework was overlayed on to the organ systems model, allowing workshop participants to detail the systemic, social, individual and biophysiological relationships that link changes in climate to each organ system and specific disease process.





### Model 3: Major Health Consequences of Climate Change (WHO)

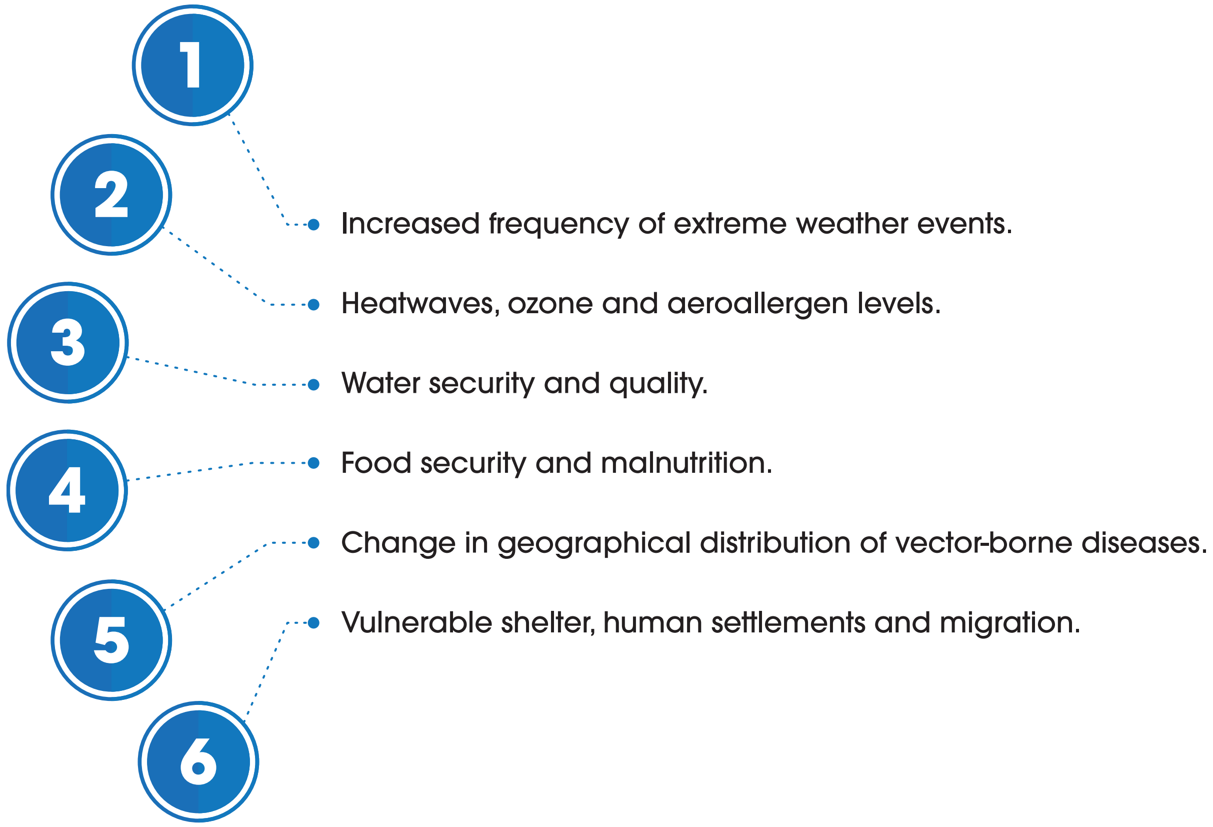


The six major consequences of climate change defined by the WHO provided distinct pathways for specific organ systems/ disease findings to be mapped. Climate change brings increasing temperatures, rising seas, and more frequent incidence of extreme weather events that impact on human health. Severe events include bushfires and associated harmful smoke, and flooding that can increase risks of water-related illnesses as well as vector-borne illnesses. Climate change impacts food production – both in terms of increased drought cycles reducing yields and diminished micronutrients in staple crops; and through the pollutants that are tied to carbon emissions (and climate change) that are also detrimental to human health. For example, today, there is greater mortality globally due to air pollution than because of HIV, malaria, and tuberculosis combined.

**Integration example:** we synthesised content into organ system chapters (Model 1), consequences of climate change as chapter sections (Model 3) and determinants of health guiding paragraph structure (Model 2). As an example, literature relating to increased incidence of acute myocardial infarction (Model 1 - cardiovascular system) was tabulated according to relationships with social determinant factors such as impeded access to healthcare and patient age (Model 2) in the context of increasing bushfires or worsening heatwaves and air pollution levels (Model 3).

### Organ System Mapping Block: Cardiovascular System

This block maps the relationships between climate change consequences and cardiovascular disease. The cardiovascular health benefits from mitigating greenhouse gases are also explored.

#### 1. INCREASED FREQUENCY OF EXTREME WEATHER EVENTS

Extreme weather events affect cardiovascular health through several pathways. Directly, the associated psychosocial stress and trauma of the event and anxiety over event recurrence are associated with an increased incidence in myocardial infarction,[40] sudden cardiac death,[41] and development of stress-related cardiomyopathy.[42] Indirectly, displacement and loss of services related to disasters is frequently associated with interruptions of medical care and access/availability of medications for chronic medical conditions.[43] Additionally, stretching of limited medical resources puts populations with chronic cardiovascular conditions at risk for life-threatening disease exacerbations. The impacts to infrastructure, possible displacement from homes and community, and reduced access to healthy food and clean drinking water may also lead to cardiovascular disease.

##### BUSHFIRE AND BUSHFIRE SMOKE

Smoke from bushfire has significant and measurable short-term impacts on cardiovascular morbidity and mortality. The extent of longer-term impacts is still not clearly understood. Bushfire smoke consists of a complex mix of particulate matter (PM) and gases which can be absorbed into the bloodstream, triggering vascular endothelial dysfunction via increased oxidative stress and systemic inflammation.[44, 45] Particulate matter 2.5 microns or smaller ($PM_{2.5}$) is associated with a variety of pathophysiological changes including deranged coagulation, blood vessel dysfunction and atherosclerotic disease,[46] compromised heart function, deep venous thromboses[47] and pulmonary embolism.[48]

Increased cardiovascular mortality rates, out-of-hospital cardiac arrests, ischemic heart disease, acute myocardial infarction and angina have been reported in both Melbourne and Sydney on high bushfire smoke days attributed to increased $PM_{2.5}$ concentrations.[49-54] A 2015 Victoria-wide study reported a 6.98% [CI95 1.03% - 13.29%] increase in risk of out-of-hospital cardiac arrests and 2% increased risk of ischaemic heart disease related emergency department attendance related to a specified $PM_{2.5}$ concentration increase.[51] A Melbourne-specific study analysed 8434 paramedic-attended out-of-hospital cardiac arrests and found an increased risk of 3.61% [CI95 1.29 – 5.99%] in out-of-hospital cardiac arrest associated with exposure to $PM_{2.5}$ particulates on either the day before or the day of cardiac arrest.[55]

> “Increased cardiovascular mortality rates, out-of-hospital cardiac arrests, ischemic heart disease, acute myocardial infarction and angina have been reported in both Melbourne and Sydney on high bushfire smoke days.”

During the 2019/2020 Australian Black Summer bushfires there were 3151 additional cardiovascular and respiratory hospital admissions, 1305 asthma presentations to emergency departments and 417 excess

#### KEY POINTS

- **Extreme weather events worsened by climate change places populations with chronic cardiovascular conditions at risk for life-threatening exacerbations of disease.**
- **Air pollution ($PM_{2.5}$ particulates) from bushfire smoke, hotter temperatures and vehicle exhausts trigger acute deranged coagulation and vascular dysfunction leading to coronary artery and cerebrovascular disease.**
- **Heat stress and dehydration due to increasing temperatures places an acute demand on the cardiovascular system. Compensatory increased heart rate impairs diastolic coronary artery perfusion predisposing to cardiac ischaemic events.**
- **Out-of-hospital mortality rates increase by up to 7% on high bushfire smoke days.**
- **The elderly, people with co-existing heart disease (heart failure, ischaemic heart disease) and those with reduced sweating capacity due to medications are particularly vulnerable to more frequent and severe heatwaves.**
- **Prescribing appropriate active transport and plant-based dietary modifications can reduce cardiovascular and stroke risks whilst reducing greenhouse gas emissions.**

deaths attributable to bushfire smoke alone in Queensland, New South Wales, the ACT and Victoria.[56] On a global scale, outdoor air pollution inclusive of bushfire smoke is being increasingly recognised as causing significant cardiorespiratory harm.[57]

##### LONG TERM EXPOSURE TO AIR POLLUTION AND REPEATED BUSHFIRE SMOKE

Repeated exposure to fine particle air pollution and other toxic compounds can be cumulative over a lifetime. Long-term health effects from low-level exposure to PM and toxic compounds in bushfire smoke are still largely unknown. However, the ESCAPE project established a 13% increase in non-fatal acute coronary events from the long-term exposure to $PM_{2.5}$ at 5 mg/m³ elevation, in addition to increasing the risk of ischaemic heart disease (IHD) and ischaemic stroke.[58] Additionally, long-term exposure to high concentrations of $PM_{2.5}$ is linked to an almost linear increase in risk of both ischaemic and haemorrhage stroke (20% [1.20, 1.15 to 1.25], and 12% [1.12, 1.05 to 1.20] respectively).[59]

#### 2. HEAT: HEATWAVES, OZONE AND AEROALLERGENS

Increasing frequency, severity and duration of heatwaves can play a major role in the exacerbation and development of cardiovascular disease. This is through both acute and chronic processes, and by direct and indirect mechanisms. The indirect effects

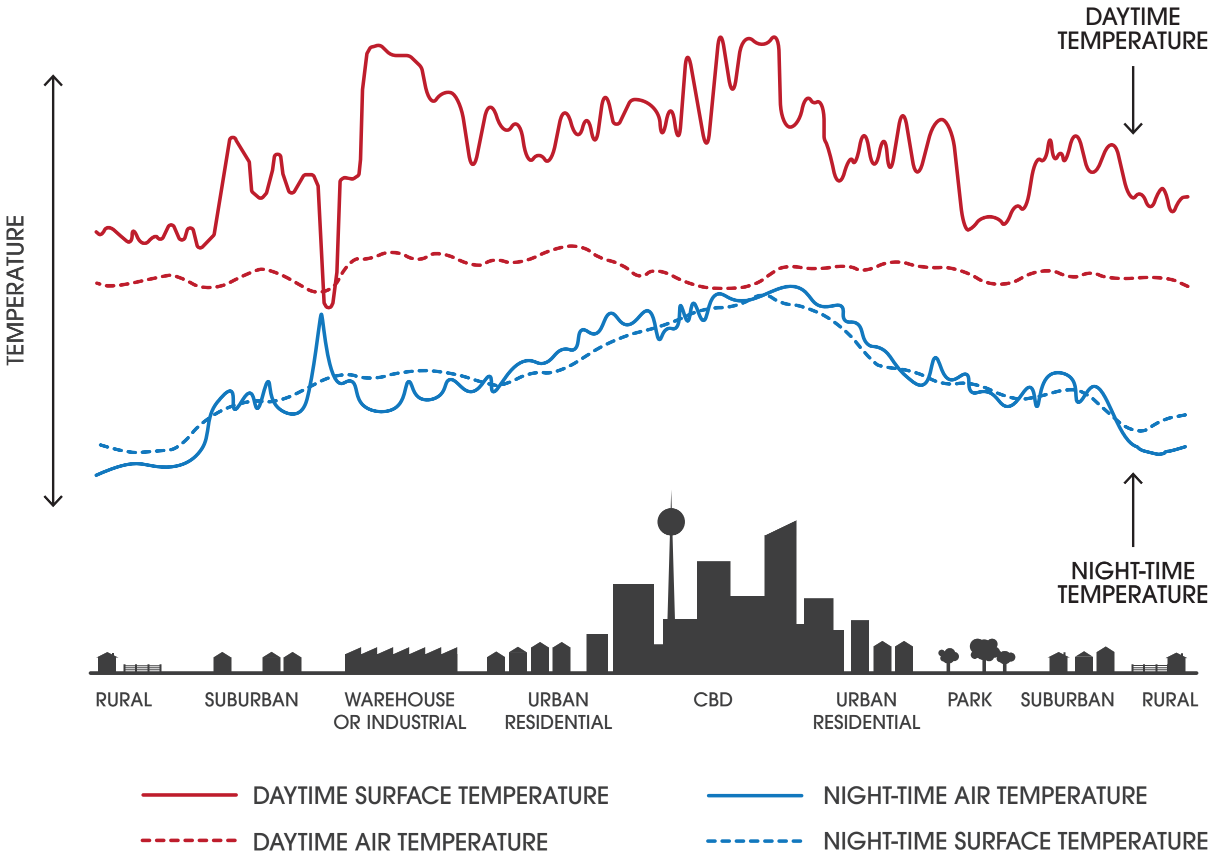


The Heat Island Effect describes why cities are often significantly hotter than surrounding areas, with implications for the health of urban populations. Structures and materials such as buildings and paved surfaces absorb and re-emit the sun's heat, whilst also providing significantly less shade and moisture than trees, vegetation, and water bodies. The dips and spikes in surface temperatures over the pond area show how water maintains a nearly constant temperature because it does not absorb the sun's energy the same way as buildings and roads. Authors own based on EPA 2020.





of heatwave cause the majority of excess mortality and morbidity and are far more difficult to identify, thus heatwaves are often labelled as 'silent killers'.[60]

The direct effects attributable to heatwaves include heat stress, heat exhaustion and heat stroke, as well as their sequelae such as organ failure and cardiac arrest.[40] On hot days if the body is unable to cool itself sufficiently, core temperature rises and can lead to heat stress.[18] Mild signs of heat stress may include dizziness, weakness or fatigue. The most severe form of heat stress is heat stroke, which can lead to cardiovascular consequences and be fatal.

Heat stress is normally associated with an acute cardiovascular response. Heated blood from the core circulation is shifted to the peripheral circulation. This acute cardiovascular response places demand on the cardiovascular system that is met through recruitment of cardiac reserve, increased cardiac output (via increased heart rate), sweat production and blood flow to the skin.[18] In context of heatwaves, whole-body heating can increase heart rate and the cardiac output by up to 7-10 l/min.[40] As heart rate increases to meet thermoregulatory requirements during hot weather, diastolic function becomes impaired. This reduces coronary artery perfusion and predisposes to acute coronary ischemia.

These mechanisms are further augmented by increased dehydration, decreased central blood volume and reduced arterial pressure,

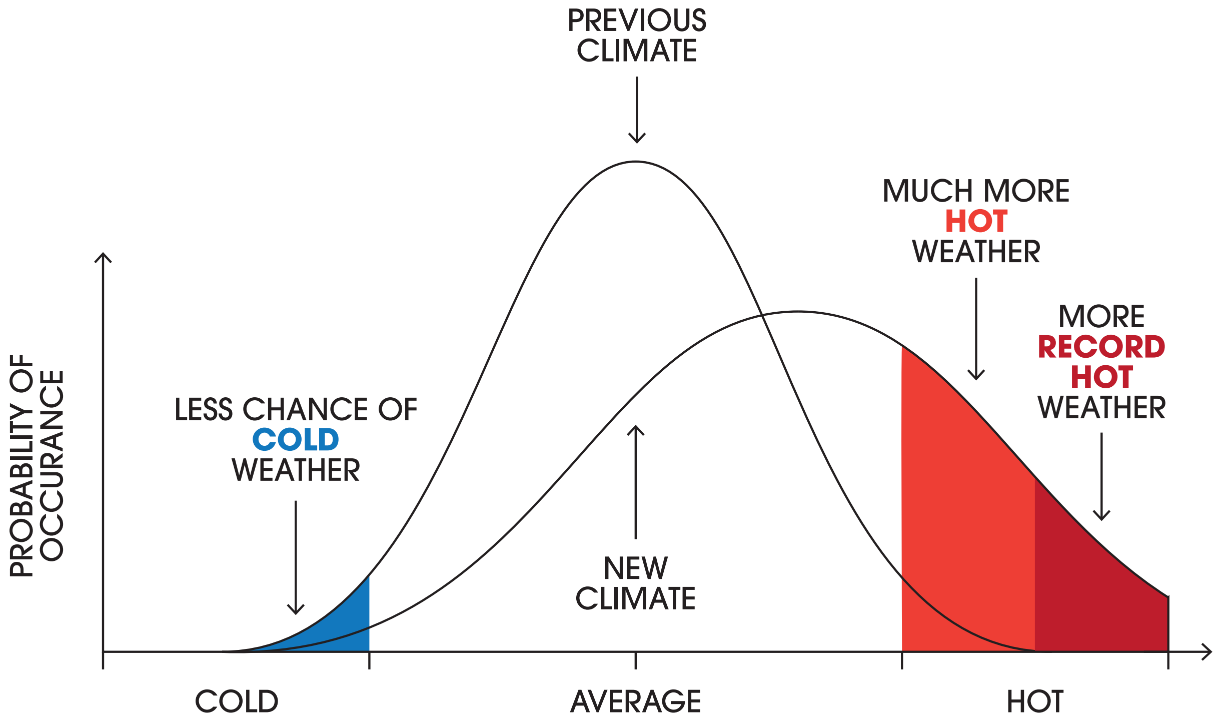


Rising greenhouse gas emissions result in much more frequent and intense hot weather (both average and extreme temperatures). There may be clinical implications for patients with pre-existing cardiovascular diseases, such as ischemic heart disease, and for pharmacological management. Authors own based on the Bureau of Meteorology 2020.

### Mechanisms for Heatwave-associated Acute Myocardial Infarction

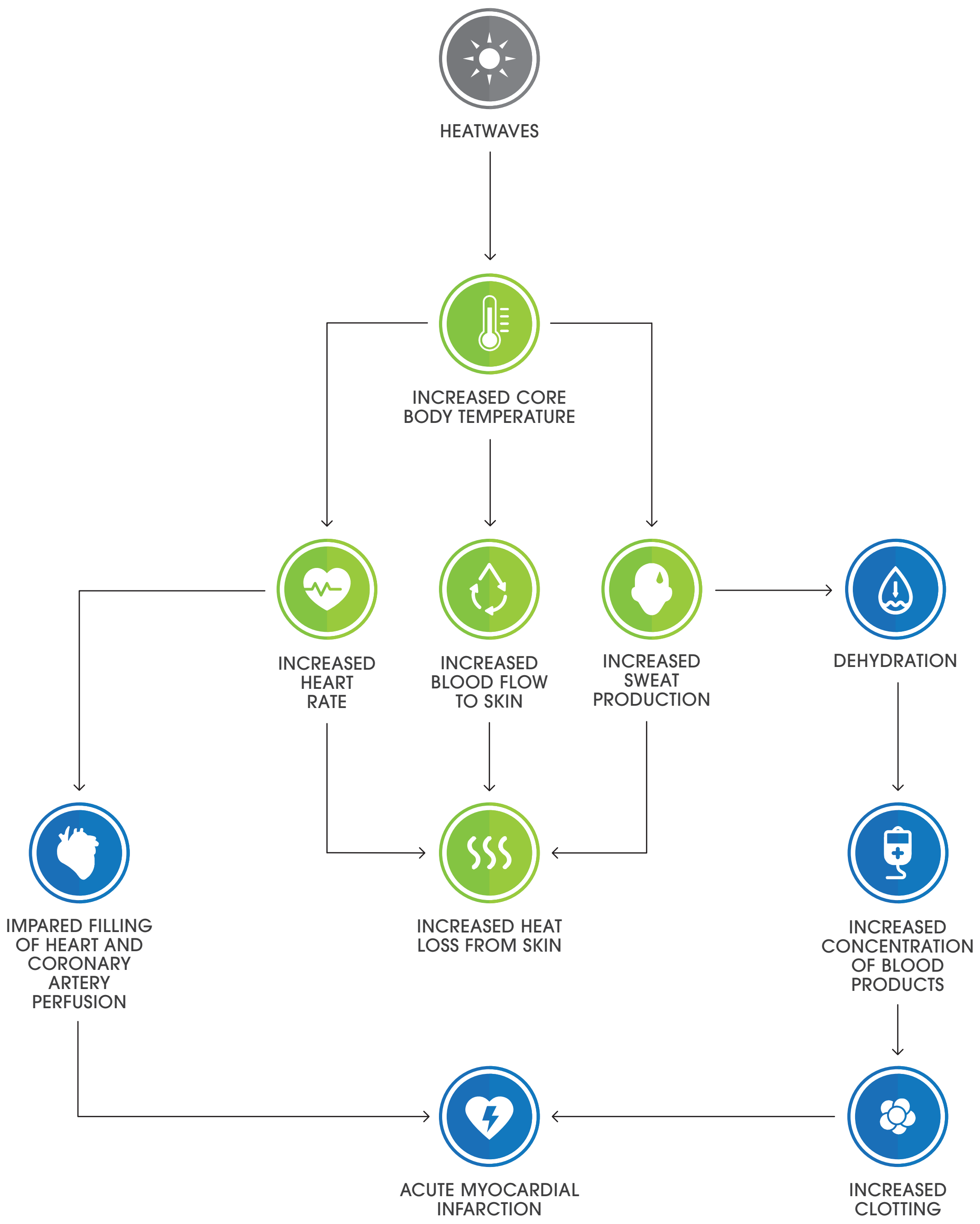


Increasing frequency, severity and duration of heatwaves can play a major role in the exacerbation and development of acute myocardial infarctions and other cardiovascular diseases. The body's core thermoregulatory responses exacerbate poor coronary artery filling. Dehydration and loss of blood volume contribute to a pro-coagulation state. These factors predispose to acute coronary syndromes. Authors' own.





particularly on standing. Reduced blood volume leads to an increase in red blood cell count, blood viscosity, neutrophils and platelet counts, along with an increase in plasma cholesterol levels without a compensatory protective change among lipoprotein fractions.[40] These factors predispose to a marked increase in arterial thrombosis, ischemia and increased mortality in hot weather.

#### CARDIOVASCULAR MEDICATION IMPLICATIONS

In context of increasing heat, medications that interfere with salt and water balance, such as diuretics, predispose vulnerable individuals to heat stroke and cardiovascular consequences.[61] A retrospective study on 94 adult patients admitted with heat-related illness following the heat wave of February 1993 in South Australia identified diuretic medications as a risk factor at presentation for symptom severity and morbidity.[62] The current evidence does not support the issuing of clear-cut recommendations regarding the use of cardiovascular drugs, but it nevertheless indicates a need for increasing the monitoring of patients and adjusting doses or temporarily suspending cardiovascular drugs accordingly, with special attention being given to diuretics, beta-blockers, renin-angiotensin-system inhibitors and anti-cholinergic drugs.[40]
In addition, those with co-morbid mental health diseases are at risk as psychotropic drugs interfere with the body's ability to regulate temperature, particularly serotonic antidepressants. Individuals being treated with these drugs could be at increased risk of heat-related illness during extreme heat events.

#### VULNERABLE GROUPS

People carrying out heavy manual labour are at risk due to the additional heat created by muscle work.[40] As temperatures increase, the physiological limits of the cardiovascular system to maintain efficient oxygenation of muscles will likely limit productivity. Therefore to stay healthy, outdoor workers must reduce their productivity with potentially serious economic losses.[63]

The elderly, people with co-existing heart disease (heart failure, ischaemic heart disease) and those with a reduced ability to sweat due to medications are particularly vulnerable. Those in lower socioeconomic groups may be reluctant to use air conditioning because of cost.

The most significant effect of high temperature on mortality occurs on the day of exposure and up to three days after exposure.[64] A predicted heat wave should prompt a rapid response to prevent heat-related deaths in vulnerable sub-groups such as the elderly. Older people are more likely to live alone, have reduced social contacts, experience co-existing chronic illness, and have a lower socioeconomic status with limited financial resources, all of which are significant risk factors for heat-related effects.[18]

Vulnerable groups living in cities are at risk due to the urban heat island effect, which occurs because of a decreased amount of vegetation and increased areas of dark surfaces in urban environments, in addition to the heat produced from vehicles and generators.[65] This results in urban centres that are several degrees warmer than surrounding areas. This effect is generally more prominent during the night than the day. It is estimated that 60% of the global population will live in cities by 2030, greatly increasing the total human population exposed to extreme heat.[66]

#### ADAPTIVE STRATEGIES

As general advice, vulnerable groups are encouraged to increase their fluid intake, including drinking without waiting for thirst. People at risk should avoid the hottest environments, wear loose-fitting clothes and take frequent showers or baths.

It is also recommended that physical activity be reduced during hot weather, and exposed people should be made aware of the symptoms of heat exhaustion and heat stroke.[67] A list of recommendations is available from the Victorian Health Department's Better Health Channel.[68]

#### 3. WATER SECURITY AND QUALITY

As a consequence of dehydration associated with other climate change related illnesses, cardiovascular sequalae may be exacerbated. These include bacterial gastroenteritis (likely to increase in incidence from contamination of human water supplies during flooding) with pathogens such as cryptosporidium and giardia and mosquito-borne diseases (Dengue fever, Ross River fever).[69, 70]

#### 4. FOOD SECURITY AND MALNUTRITION

##### HEAT, FOOD SUPPLY AND CARDIOVASCULAR DISEASE

Rising temperatures are leading to drought, damaged crops and low yields that undermine food and water supplies. Decreased food and water supply leads to rising costs that tend to disproportionately disadvantage people on lower incomes. This may contribute to reduced access to nutritious food and consequently predispose individuals to obesity and cardiovascular disease.[71]

Although the net effect of climate change on pollinators remains uncertain, a reduction in animal pollination would decrease yields of numerous pollinator-dependent food crops central for food and micronutrients.[72] A decline in global pollination, for example from reduced bee numbers unable to survive increasing heatwave events, would result in reduced dietary intake of fruits, vegetables, nuts, and seeds, increasing the risk of heart disease, stroke, diabetes, and certain cancers in adults.[72]

##### SEA-LEVEL RISE IMPLICATIONS

An increase in sea-level rise is leading to increased sodium intake through the food chain. Increasing salinisation of underground freshwater aquifers[72, 73] combined with increasing ambient temperatures are depleting and concentrating groundwater resources. As a result, reduced crop yields from saline contamination may lead to higher food prices and more food shortages. The use of saltier water for crop irrigation is already leading to increased sodium uptake within both plant and animal food sources.[74, 75] The increasing consumption of dietary sodium is a key risk factor for primary hypertension.[76, 77]

#### 5. CHANGE IN GEOGRAPHICAL DISTRIBUTION OF VECTOR BORNE DISEASES

The incidence of certain vector-borne and zoonotic diseases indirectly increase the risk for cardiovascular disease. Infections that are likely to involve dehydration pathways are most associated with cardiovascular sequelae. In Australia these are Dengue fever and Ross River fever, particularly as the incidence of tropical disease moves south with increasing ambient temperatures. In the developing world, approximately 10% of strokes are related to exposure to certain vector borne and zoonotic diseases,[78] many of which are climate sensitive. In particular, Chagas disease is a major cause of stroke worldwide. Twenty million people globally have chronic Chagas, which is an independent risk factor for stroke in Latin America,[79] and a leading cause of heart failure in South America.[80]

#### 6. VULNERABLE SHELTER AND HUMAN SETTLEMENTS AND MIGRATION

Children, women of childbearing age, and the circumstances of people experiencing irregular migration, people fleeing conflict, refugees,





asylum seekers and internally displaced people place them at high risk of developing and exacerbating cardiovascular diseases.[81]

These risks may also apply to Australians who are displaced and living in temporary shelter as a result of flooding or bushfire events. Risks during irregular migration and/or in temporary shelter include threats to physical safety, sexual and gender-based violence, inadequate nutrition/access to food and clean water, lack of access to health services, distress caused by separation from families and trauma related to the cause of displacement. Additionally, people living in protracted temporary shelter or refugee situations face long term unemployment, inadequate or lack of access to health services and interrupted schooling. These determinants of health are central to an individual's and population's risk for developing cardiovascular disease.

#### CARDIOVASCULAR HEALTH CO-BENEFITS OF GREENHOUSE GAS MITIGATION

There are known cardiovascular health co-benefits from strategies that reduce greenhouse gas emissions. For example, prescribing appropriate active transport and plant-based dietary modifications can reduce cardiovascular and stroke risks whilst reducing greenhouse gas emissions.[38]

A plant-based diet contributes to protecting cardiac health by reducing red meat consumption which is a known risk factor for ischaemic heart disease.[37] Animal products have much higher greenhouse gas emissions per gram of protein than plant-based alternatives.[32] For example, beef and lamb have emissions per gram of protein that are about 250 times higher than legumes.[24]

The term 'active transport' refers to modes of transport that involve more physical activity to get around, rather than the use of private motor vehicle. Active transport encompasses walking, running and cycling, often in combination with public transport, such as buses, trains, underground trains or boats. Daily exercise can be performed other than by active transport, but for many people incorporating exercise as part of their daily commute is the simplest and most organic method. There are a number of measures that could be taken to facilitate greater use of active transport. For example, building better roads suited to cyclists would achieve health improvements worth 10–25 times the cost of road improvements.[82]

> "Increased active travel can reduce greenhouse gas emissions, vehicle air pollution and address physical inactivity, all risk factors for ischaemic heart disease, stroke, and diabetes."

Increased active travel can reduce greenhouse gas emissions, vehicle air pollution and address physical inactivity, which contributes to more than 3 million deaths a year worldwide[2] and is a risk factor for major non-communicable diseases such as ischaemic heart disease, stroke, and diabetes.[37] Each extra hour a person spends in a car per day is associated with a 6% increased risk of obesity.[26] A study in Copenhagen showed that, after adjustment for age, sex, educational level, leisure time activity, body mass index, blood lipid levels, smoking and blood pressure, individuals who cycle to work had a 28% lower relative risk for all-cause mortality (0.72) than those who did not.[83]

> Caring for Country has been linked to a broad range of cardiovascular and environmental benefits that positively impact Indigenous peoples' health and wellbeing.

#### DISEASE PREVENTION STRATEGIES

Primary preventive measures can also limit the expected effects of climate change on cardiac health. Increased patient awareness of the health consequences of climate change on their cardiovascular health, increased access to sustainably air-conditioned environments (ie. air-conditioners powered by renewable electricity), increased physician and hospital preparedness for responses to acute heat-related events and heat-wave alert response systems are all important to emphasise and establish.[84] It is also important to note that such measures are focussed on helping the community to adapt to the consequences of climate change, some of which are now unavoidable, rather than addressing the upstream causes.

#### IMPROVING CARING FOR COUNTRY

Improving understanding of Caring for Country has been linked to a broad range of cardiovascular and environmental benefits that positively impact health and wellbeing, particularly for Indigenous peoples'.[36] Among people who took part in Indigenous Cultural and Natural Resource Management (ICNRM) courses, especially when living in their traditional country, significant health benefits were found: more frequent exercise, lower rates of obesity, lower rates of diabetes, lower rates of renal disease, lower rates of cardiovascular disease, and less psychological stress.[36] Aboriginal participants in the study supported the idea that the majority of benefits from ICNRM, both personal health benefits and benefits to landscape health are derived from the sense of wellbeing that comes from maintaining or re-establishing cultural connections to country and the more obvious influences of a more nutritious diet and more exercise.[36]

Traditional caring for country practices that involve harvesting of native flora also can have positive outcomes for the environment. The harvesting and cultivation of bush foods, such as wild wattle seeds, bush tomatoes and native millet, can help propagate and re-establish these species in areas where they might otherwise be out competed or over-predated by exotic and pestilent species.[33] These plant-types also have a significantly lower carbon footprint than contemporary agricultural approaches.



### Determinants of Health for Aboriginal and Torres Strait Islander Peoples

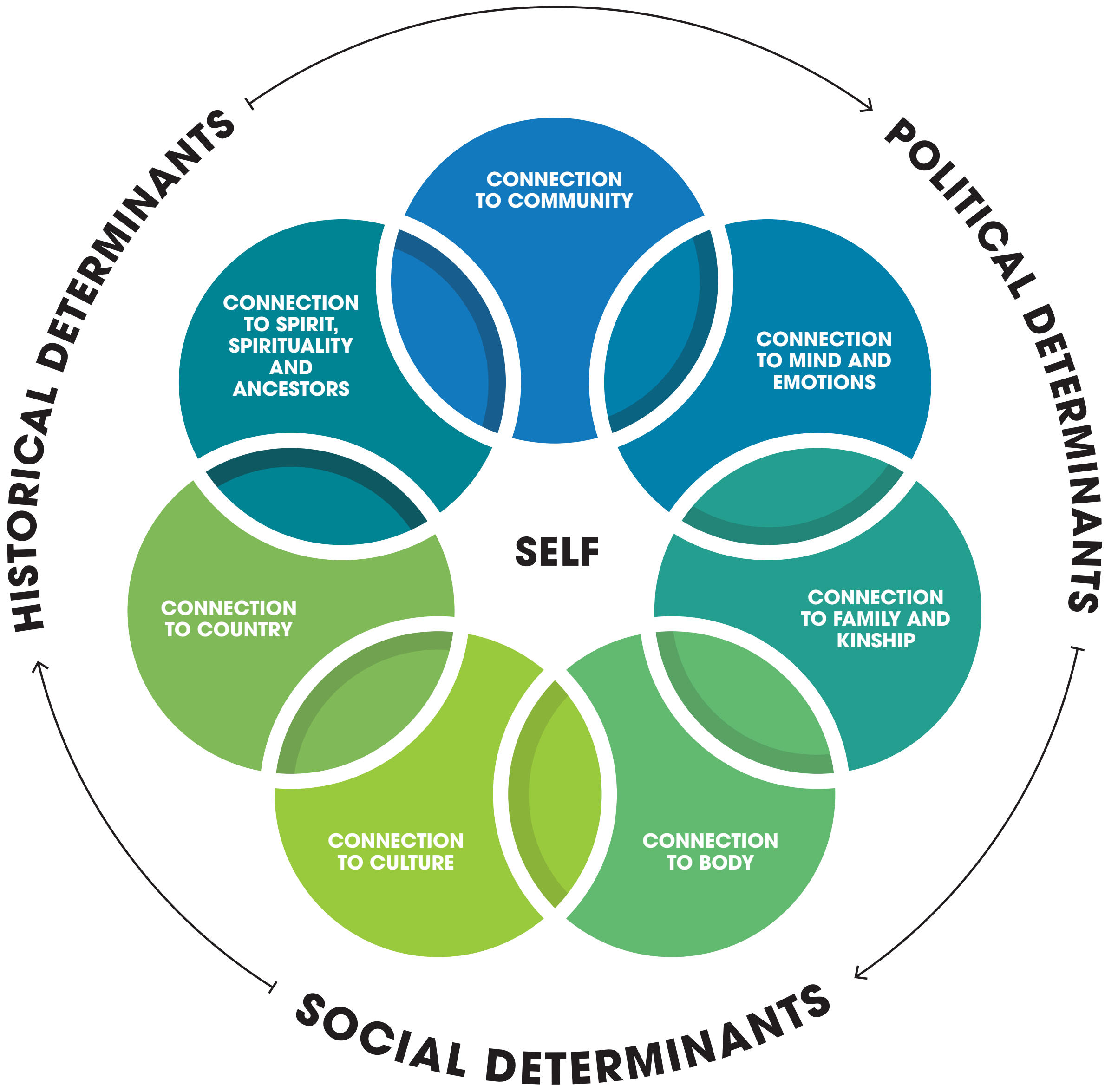


Connection and caring for the environment are recognised as being central to individual wellbeing by the world's oldest cultures. Authors own based on the National Strategic Framework for Aboriginal and Torres Strait Islander Peoples' Mental Health and Social and Emotional Wellbeing 2017.

### Organ System Mapping Block: Respiratory System

This block maps the relationships between climate change consequences and respiratory disease. The respiratory health benefits from mitigating greenhouse gases are also explored. Limited literature search findings were available for sections 3, 4 and 5.

#### 1. INCREASED FREQUENCY OF EXTREME WEATHER EVENTS

Allergic diseases, including asthma, hay fever, rhinitis, and atopic dermatitis are known to increase in prevalence during extreme weather events.[23, 85, 86] Increased exposure to pollens (due to longer growing seasons), moulds (from extreme or more frequent rainfall), air pollution and dust (from droughts) are all exposure pathways to disease.[23] Prolonged drought will lead to more dust and particulate pollution, while increased rainfall might cleanse the air but can create more mould and microbial pollution.[87]

Extreme weather events may also place health infrastructure at risk and affect patient access to healthcare services. A recent study in the Journal of the American Medical Association found that lung cancer patients undergoing radiation were less likely to survive when hurricane disasters disrupted their treatment regimens.[88] Radiotherapy is particularly affected because it requires dependable electrical power and daily treatment.[89] Healthcare infrastructure in particular appears to be more vulnerable than other corporate infrastructure to extreme weather event.[90, 91]

##### BUSHFIRE SMOKE

Bushfire smoke impacts on acute and chronic respiratory conditions via the inhalation of particulate matter, gaseous compounds and carcinogenic chemicals.[52, 92] During a bushfire, large areas of land can be covered in layers of smoke hundreds of kilometres away from the actual fires, potentially involving major cities and exposing millions of people to bushfire smoke. The air pollutant that increases most significantly as a result of bushfire smoke is particulate matter (PM), which is created directly during the combustion process and also formed later from the emitted gases.



Short-term exposure to larger particles with an aerodynamic diameter of 10µm ($PM_{10}$) leads to irritation of the eyes and throat, whereas smaller fine particulates are inhaled deep into the lung parenchyma and are comparable to the smoke from cigarettes.[93, 94] Inhalation of fine particulate matter with aerodynamic diameter of less than 2.5µm ($PM_{2.5}$) is most strongly associated with exacerbation of asthma, chronic obstructive pulmonary disease and chronic bronchitis.

> "Smaller, fine particulates are inhaled deep into the lung parenchyma and are comparable to the smoke from cigarettes."

Across 184 days where metropolitan Sydney was affected by bushfire smoke, an additional 787 respiratory hospitalisations were attributable to fire smoke on those days (in addition to an additional 436 cardiovascular hospitalisations and 197 premature deaths).[95] A Queensland study observed an increase in respiratory hospital admissions of 19% for bushfire days, with effect sizes known to be larger for Aboriginal and Torres Strait Islander peoples, particularly those experiencing chronic obstructive pulmonary disease (COPD).[92]

##### THUNDERSTORM ASTHMA

Associations between thunderstorms and asthma morbidity have been identified in multiple locations around the world. Thunderstorm asthma is characterised by

#### KEY POINTS

- **Air quality and respiratory diseases are affected through several pathways, by both global greenhouse gas concentrations and local air pollution. These include hotter temperatures, bushfire smoke, dust during drought, car and truck exhausts and nearby power stations.**
- **Inhalation of air pollution ($PM_{2.5}$) into lung parenchyma increases exacerbations of chronic obstructive pulmonary disease and chronic bronchitis and the prevalence of allergic diseases including asthma.**
- **Fine particles overcome the respiratory mucosal barrier, triggering inflammation and oxidative stress. Larger air particles ($PM_{10}$) lead to irritation of the eyes and throat.**
- **Lifelong exposure to air pollution and bushfire smoke may predict a 20% increased risk of lung adenocarcinoma, with particulate matter inhaled deep into the lung parenchyma comparable to the smoke from cigarettes.**
- **Increased $CO_2$ concentrations promotes the production of more pollens and stronger pollen IgE-binding capacity – with significant ramifications for allergic airways disease.**
- **The risk of developing asthma in childhood increases by 14% for every 2 µg/m3 incremental increase in chronic exposure to traffic-related particulate matter.**

#### HOW DOES CLIMATE CHANGE IMPACT RESPIRATORY HEALTH?

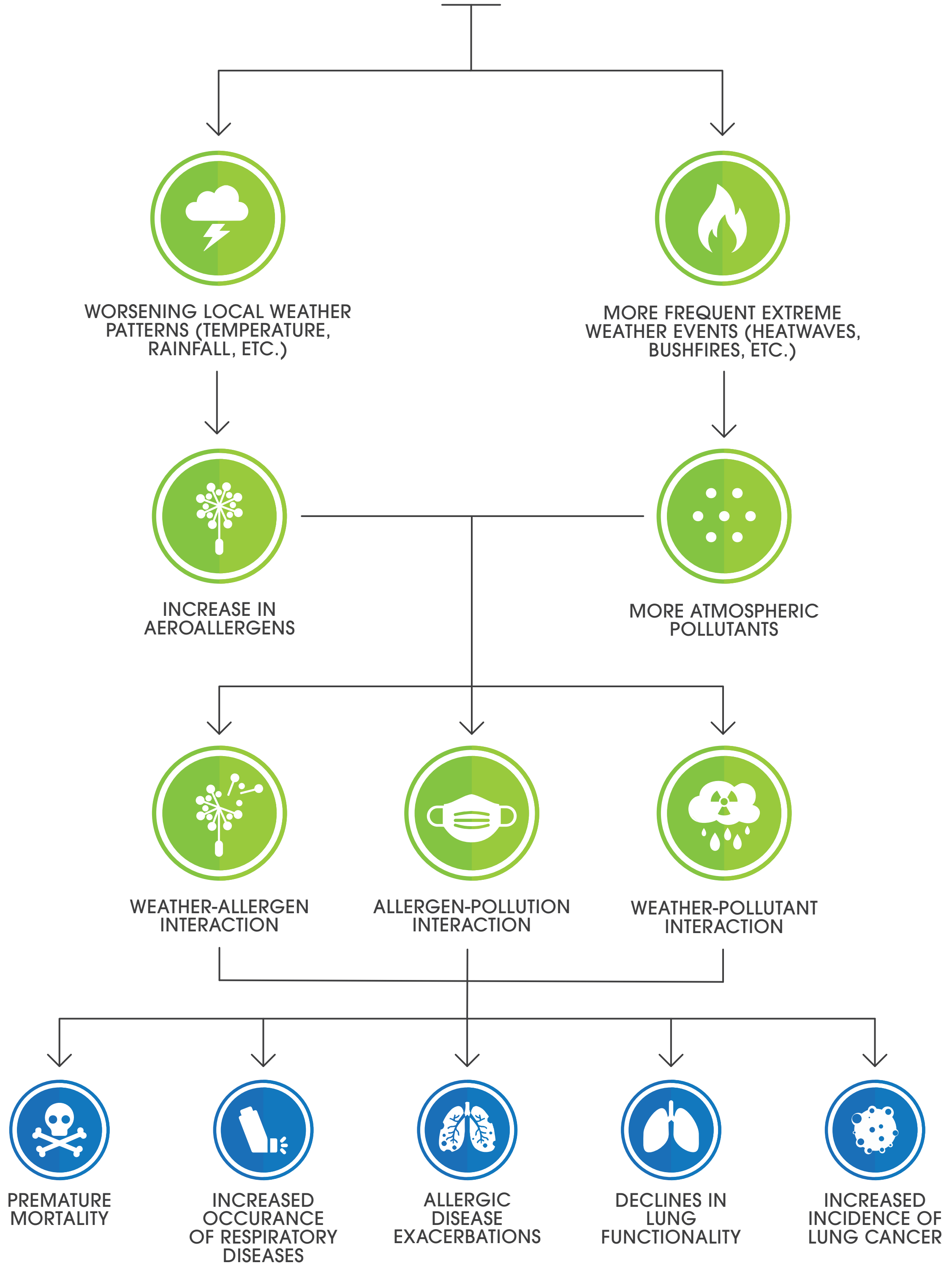


More intense and more frequent extreme weather events such as storms, bushfires and heatwaves are driving higher aeroallergen and air pollution levels. These environmental changes exacerbate chronic respiratory diseases (such as asthma and chronic obstructive pulmonary disease), worsen allergic responses and can contribute to incidence of lung cancer. Authors own based on De Sario and Michelozzi 2013.





asthma outbreaks possibly caused by the dispersion of allergenic particles derived from osmotic rupture of pollen and spores.[23, 96] The thunderstorm-related epidemics are usually limited to late spring and summer when there are high levels of airborne pollen grains. There is a close temporal association between the arrival of a thunderstorm, a major rise in concentration of pollen grains and the onset of asthma epidemics. According to current climate change scenarios, there will be an increase in intensity and frequency of heavy rainfall episodes, including thunderstorms, over the next few decades, which can be expected to be associated with an increase in the number and severity of asthma attacks both in adults and in children.[23] In Melbourne, two large asthma outbreaks caused a major increase in the number of hospital attendances and admissions. This was due to an increase in asthma exacerbation (five- to ten-fold rise) and significant increases in hay fever and rye grass pollen allergy.[97]

##### INDIRECT AND LONG-TERM TMPACTS OF EXTREME WEATHER EVENTS

For people living directly in bushfire or flood regions, psychological reactions such as stress, anxiety and depression can contribute to exacerbation of respiratory diseases.[98, 99] Those most likely to be affected by bushfire smoke include people with existing cardiovascular or respiratory conditions such as asthma, chronic obstructive pulmonary disease and chronic bronchitis, as well as pregnant women, older people and young children.[92] High stress for adults after weather-related disasters can have serious implications for children. For example, an adults' ability to help manage asthma attacks can be impeded. In extreme circumstances, the associated stress barriers might result in neglect.

Ongoing exposure to bushfire smoke poses long-term health risks to all people exposed, with lifelong exposure to wood smoke being

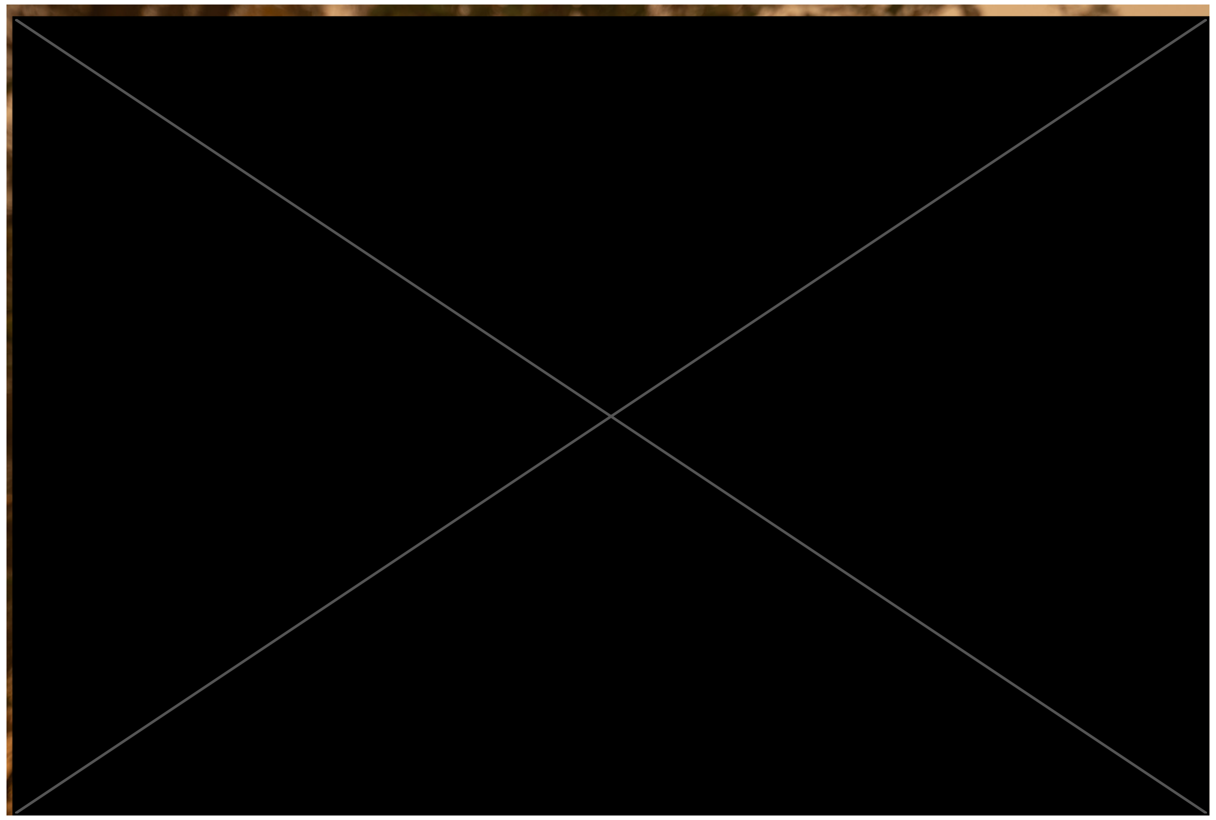

Bushfires in Eden, Australia in December 2019. Pre-existing respiratory conditions such as asthma or COPD and high emotions and stress, are all risk factors for poor respiratory outcomes during extreme weather. Photo source: Joachim Zens / Shutterstock

associated with a 20% increase in risk of lung adenocarcinoma.[93, 100] Bushfire smoke can include carbon monoxide, polyaromatic hydrocarbons, oxides of nitrogen and volatile organic compounds - specifically formaldehyde, acetaldehyde, acrolein, benzene and toluene. Some of these substances act as respiratory irritants and may also be cancer-causing.

#### 2. HEAT: HEATWAVES, OZONE AND AEROALLERGENS

Heat and air pollution contribute directly to deaths from cardiovascular and respiratory disease. Air quality is affected by both global greenhouse gas concentrations in addition to local air pollution mostly from bushfire smoke, car and truck exhausts or power stations. Poor air quality and subsequent respiratory diseases are occurring through several pathways. These include heat mediated increases in production of pollen and mould spores and increases in ambient concentrations of ozone, fine particles, organic gases and dust. These pollutants directly cause respiratory disease or exacerbate respiratory disease in susceptible individuals.

Temperature and air pollution factors that affect the respiratory system include:[101]

- Outdoor temperature
- Air pollution
- Pollen production, pollen season length and allergenicity of certain pollens
- Ground level ozone
- Heat-related stress
- Local air pollution
- Bushfire smoke (detailed above)

The underlying mechanisms are different for the various pollutants. Similarly, the relationship between air pollution, pollen exposure and respiratory allergy is based on an individual's response to air pollution, which depends on the source and components of the pollution, as well as on climatic agents.

##### OUTDOOR TEMPERATURE

Heat and associated air pollution are particularly harmful to populations over 75 years old and those with pre-existing respiratory conditions, such as asthma, allergic rhinitis, bronchitis and COPD. Currently over summer, each degree celsius increase in maximum apparent temperature (a combined indicator of temperature and humidity) above a city-specific threshold is related to a ~7% increase in daily respiratory deaths.[86] A smaller, but still significant effect, has been observed for respiratory hospital admissions (3–5%). The effect of extreme weather events, i.e. heatwaves, is much larger, accounting for an increase in respiratory mortality during each heatwave day ranging from 12.1% to 61.3%. Inhalation of hot and humidified air activates airway sensory nerves (most likely thermosensitive C-fibre afferents), triggering transient bronchoconstriction in patients with asthma (cholinergic reflex). This is augmented if exposed to prolonged heat, where changes to ventilation rate and tidal volume increase the total intake of airborne pollutants.

##### AIR POLLUTION

Air pollution has a significant impact on asthma, in addition to rhinosinusitis, COPD and respiratory tract infections.[86] Fine particles (fewer than 2.5 micrometres in diameter, or $PM_{2.5}$) are small enough to be inhaled deep into the lungs, overcome the mucosal barrier and embed in lung parenchyma. These foreign particles induce airway inflammation and oxidative stress, resulting in decreased lung function and contributing to increased respiratory symptoms (coughing, dyspnoea and sputum production), decreased lung func¬tion, aggravated asthma, development of chronic bronchitis, development of arrhythmias, nonfatal heart attacks, and premature death in people with heart or lung disease.





Particulate matter 2.5 micrometres in diameter also diffuse across the alveolar-arterial membrane, with increased exposure associated with an increased risk of emergency department visits and hospitalizations for cardiorespiratory diseases, especially among adults over 65 years of age.[102] For larger $PM_{10}$ particles, for every 10 mg-m³ increase of $PM_{10}$ and sulphur dioxide there is an associated 0.5–1.5% increase in admissions for asthma and COPD.[102] Finally, air pollution also makes individuals more susceptible to pollen or dust triggers, by inducing airway inflammation and mucosal barrier dysfunction, leading to priming of allergen-induced responses.[103] In a psychosocial context, decreased air quality and pollution are known to affect an individual's ability to work or to attend school via the contribution to respiratory and cardiovascular disease. Thus, extreme temperatures and air pollution tend to adversely impact individuals in the lower socioeconomic groups more so than higher socioeconomic groups, contributing to the cycle of vulnerability. Children (up to 14), adults over 65, smokers, pregnant women and those with existing cardiovascular or respiratory are most susceptible to the health impacts from air pollution.[104]

| CLIMATE CHANGE EVENT | POTENTIAL ENVIRONMENTAL IMPACT | EFFECT ON ALLERGIC DISEASE PREVALANCE |
|---|---|---|
| **Increase in temperature.** | Migration of stinging and biting insects into new environments and increased population of existing insect species.<br><br>Change to crop patterns, with the potential to introduce new allergenic pollens into the atmosphere and new food proteins into local diets. | Sensitisations to new stinging and biting insect species and to foods, with the potential to increase cases of IgE-mediated anaphylaxis. |
| | Earlier and longer pollination seasons.<br><br>Increases in humidity associated with higher temperatures will lead to increased numbers of cockroaches, house dust mites and moulds, thus increasing allergen loads. | New pollen and mould sesitisations, leading to increased prevalence and attacks of allergic rhinoconjunctivitis and asthma; longer pollen seasons leading to increased duration of symptoms. |
| **Increase in precipitation and drought, leading to, damaged crops, lower yeilds, food shortages and less work.** | Population migration. | Development of sensitisation to new allergens, leading to the development of allergic respiratory and skin conditions. |
| **More frequent thunderstorms in spring and summer.** | Thunderstoms cause pollen grains to rupture, increasing the levels of respirable allergens and also lead to an increase in ozone levels. | Increased hospital admissions due to asthma. |

Effects of climate change on the prevalence of allergic diseases. Impacts include increased incidence of IgE-mediated anaphylaxis, thunderstorm asthma and hospitalisations due to more frequent and severe allergic disease. Authors own based on Pawankar et al. 2008 and D'Amato et al. 2011.

##### POLLENS

Increased temperature and carbon dioxide concentrations increase photosynthesis and metabolism by allergenic plants, affecting the timing, duration and volume of particular pollens.[105] As temperature increases, plants produce more pollen for longer periods of time, intensifying the allergy seasons.[106] Earlier and greater rates of flower blooming will occur and increase the subsequent risk of exposure amongst allergenic individuals. Climatic change will also increase the amount of allergenic proteins contained in pollens,[23] as carbon dioxide concentrations alter soil pH and confound protein composition and strength of IgE-binding.[107]

##### GROUND LEVEL OZONE

Ozone ($O_3$) exposure has both a priming effect on allergen-induced responses and an intrinsic inflammatory action in the airways.[107] Approximately 40-60% of inhaled $O_3$ is absorbed in the nasal airways, the remainder reaching the lower airways. Exposure to $O_3$ induces free radical production that leads to epithelial and type 1 alveolar cellular injury and significantly increases levels of inflammatory cells (in particular neutrophils) and mediators such as IL-6, IL-8, granulocyte-macrophage colony-stimulating factor (GM-CSF) and fibronectin amongst asthmatics.[23] This inflammatory response increases epithelial permeability, increases airway hyperreactivity to bronchoconstrictor agents and is associated with reduced lung function and increased exacerbation of asthma and development of chronic bronchitis.

Physical activity whilst exposed to increased concentrations of ground level ozone leads to increased rates of asthma exacerbations, emergency departments visits and admissions for respiratory diseases. This is likely due to increasing lung volumes and respiratory frequency resulting in greater deposition of air pollutants and $O_3$ into the lower airways. Repeated exposure to ground level ozone may also permanently scar lung tissue.[108]

> "A significant proportion of asthmatic episodes are triggered by environmental factors, including ambient air pollutants, allergens and stress due to external environmental events."

##### TEMPERATURE AND STRESS IMPACTS

Hotter temperatures increase cortisol release, impair executive function and behavioural responses, decrease the capacity of both working and short-term memory, reduce sleep quality and disrupt people's physical activity routines. This results in reduced wellbeing and increased psychological distress that may have an impact on an individual's coping responses, behavioural patterns, relationships and adherence with best practice management for conditions such as asthma. Melbourne is projected to experience 12-17 additional days above 35 degrees Celsius per year under the Paris Target compared to 2020. The most vulnerable members of society - the very old, the very young, Aboriginal and Torres Strait Islander communities, and those who work outdoors are most vulnerable to exposure to extreme heat and are disproportionately



represented amongst those experiencing psychological distress and poor mental health during these periods.

##### LOCAL AIR POLLUTION

In Melbourne, car and truck exhausts are large contributors to local air pollution. They release cytotoxic gases and particulates such as nitrogen oxides, sulphur dioxide and particulate matter into the air. Exposure to traffic-related air pollution (TRAP) is a significant contributor to the prevalence of all-cause mortality (14% increase with every 10 µg/m³ increase in $PM_{2.5}$ over 10 years), lung cancer mortality (37% increase with every 10 µg/m³ increase in $PM_{2.5}$ over 10 years) and to existing and new onset of asthma and allergy.[102] Proximity to major roads correlates with exposure to TRAP. Cars and truck exhausts are significant sources of outdoor nitrogen dioxide ($NO_2$).

$NO_2$ exposure is associated with increased emergency room visits, wheezing and medication use among children with asthma. $NO_2$ also enhances the allergic response to inhaled allergens and contributes to increased ground level ozone formation. As a greater proportion of people undertake behaviour modifications during hot conditions, such as the use of vehicles and air conditioning, localised differences in microparticle concentrations occur and amplify broader air pollution concentrations resulting from temperature and carbon dioxide concentration changes. A 2015 meta-analysis found that the risk of developing asthma in childhood increases by 14% for every 2 µg/m³ incremental increase in chronic exposure to traffic-related particulate matter.[109]

#### 6. VULNERABLE SHELTER AND HUMAN SETTLEMENTS AND MIGRATION

Migration involves exposure to a new set of pollutants and allergens as well as changes in housing conditions, diet, and accessibility to medical services, all of which are likely to affect migrants' health. The type and severity of consequences will vary depending on whether migration is forced and/or irregular, and therefore often involving vulnerable shelter,

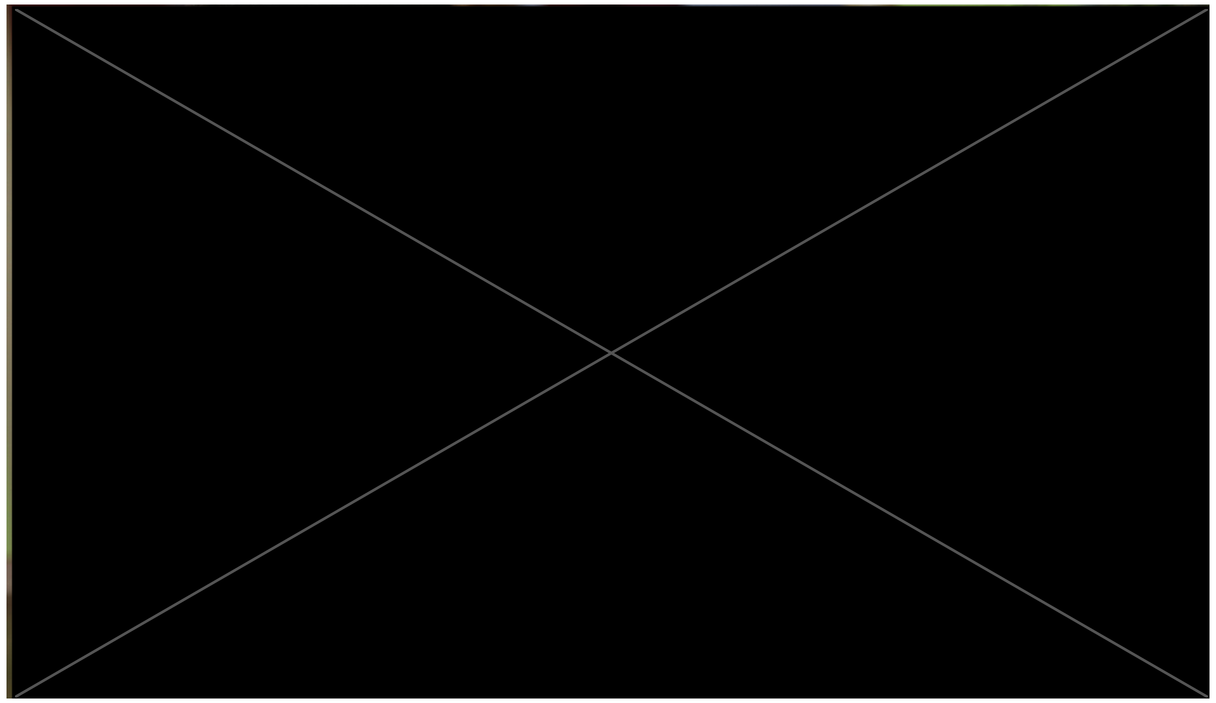

Poor air quality, from more aeroallergens and more air pollution due to climate change, can exacerbate asthma and other respiratory diseases.

or planned and safe migration as a result of climate changes. Forced migration confers risk for airborne diseases such as tuberculosis that are prevalent within refugee and migrant camps.

In regard to migration to developed nations, atopy and asthma are more prevalent in developed and industrialised countries. The effect of migration on respiratory health is therefore age and time dependent where early age and longer time spent in the new environment increase the likelihood of developing allergenic symptoms, such as asthma, rhinoconjunctivitis, or eczema.[110] Migrants should be aware of the potential for developing allergies and/or asthma.

#### RESPIRATORY HEALTH CO-BENEFITS OF GREENHOUSE GAS MITIGATION

Many strategies that mitigate greenhouse gas emissions and local air pollutants also provide multiple co-benefits to respiratory health.[35] For example, in Melbourne, where the main source of air pollution is motor vehicle emissions[20] increasing active travel (walking and cycling) can reduce greenhouse gas and fine particulate emissions. Alternatively, increasing urban greenspace decreases levels of air pollutants that may trigger asthma.[25, 29, 34] It should be noted that it is important to select tree species that are not highly allergenic.[29]

##### DISEASE PREVENTION STRATEGIES

Management of asthma and other respiratory allergic diseases relies on strict control of exacerbation triggers[111] combined with effective behavioural and pharmacological management. A significant proportion of asthmatic episodes are triggered by environmental factors, including ambient air pollutants, allergens and stress due to external environmental events.[23] One challenge for health professionals and patients alike will be ongoing management of changing environmental triggers.

Incorporating the following environmental considerations may support effective asthma control or demonstrate the practice of medicine in an environmentally sustainable way:

General advice for all populations:

- People at risk should also avoid the hottest environments and consider a reduction in physical activity during hot weather or consider pre-medication prior to exposure.
- Weather forecasts and conditions should be monitored.
- Individuals who are highly sensitive to pollens should be provided with appropriate adaptations to their management plans to reduce their exposure time outside.
- Facemasks to protect from fine particulate inhalation and pollens should be considered.
- These components should be incorporated into asthma action plans.

Promoting increased active travel can reduce greenhouse gas, ozone and fine particulate emissions with a myriad of health benefits including reduced rates of asthma, lung cancer, heart disease and stroke.[20] These benefits occur due to reduced local air pollution and from broader climatic stability resulting from reduced vehicle emissions.

Local air quality co-benefits can provide complementary benefits such as reduced healthcare expenditure, fewer lost workdays due to sickness, improved agricultural crop yields and support for progress on related Sustainable Development Goals.



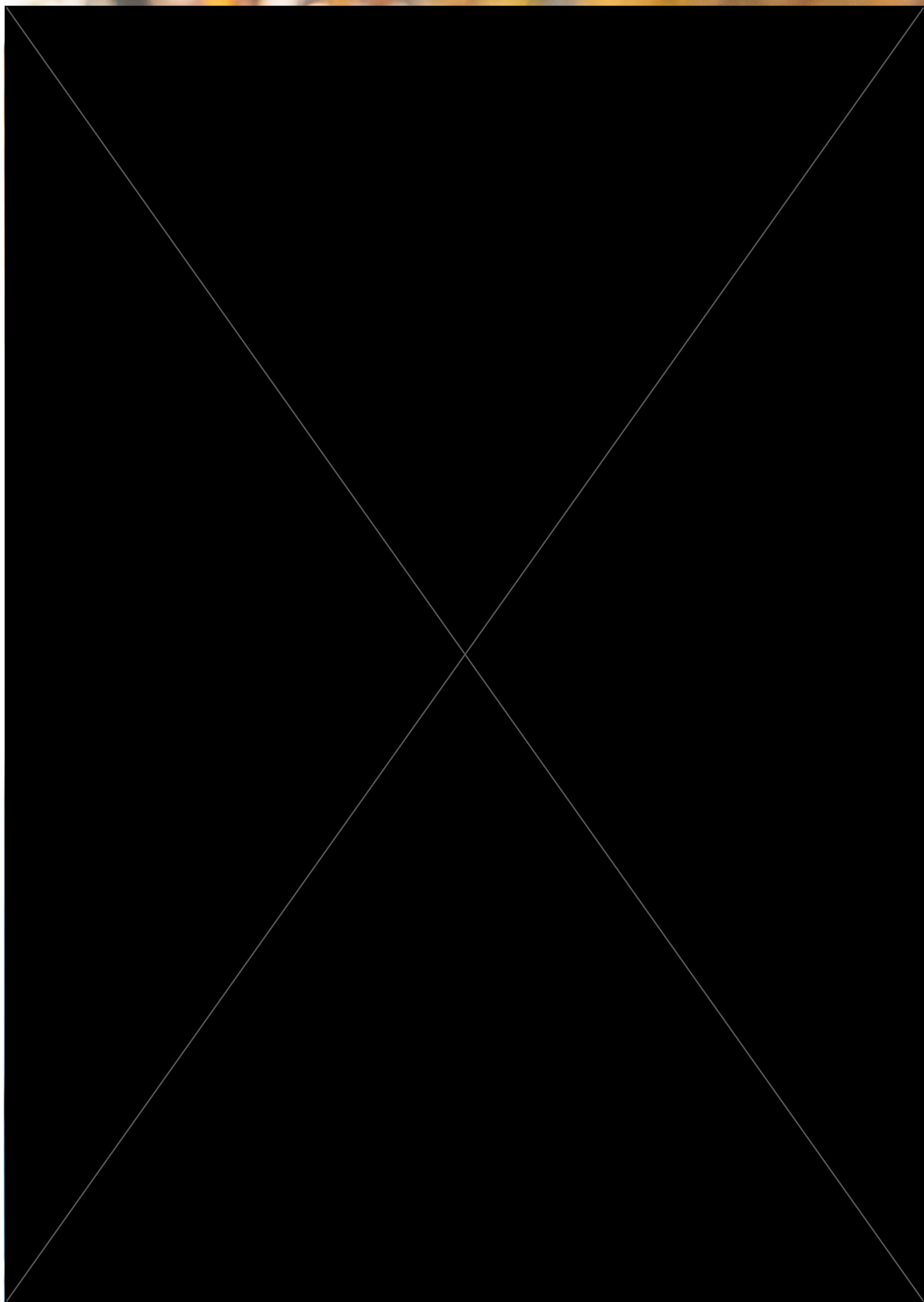

### Organ System Mapping Block: Renal System

This block maps the relationships between climate change consequences and renal disease. The renal health benefits from mitigating greenhouse gases are also explored.

#### 1. INCREASED FREQUENCY OF EXTREME WEATHER EVENTS

Damage or impeded access to healthcare infrastructure due to extreme weather can interfere with delivery of dialysis to patients with end-stage kidney disease.[112, 113] Given that dialysis is a life sustaining therapy, the impact is life-threatening.

Damage or lack of access to health facilities, contamination of water supplies or severe drought can increase the risk of missed dialysis or hospitalisation leading to severe electrolyte imbalances, fluid overload, acidaemia, and potentially death. Furthermore, patients may need to relocate and experience prolonged periods of displacement from family and social supports.[114] Extreme weather events also impact the availability of critical medical care and medications including immunosuppressants for kidney transplant recipients.[115]

#### 2. HEAT: HEATWAVES, OZONE AND AEROALLERGENS

Hotter temperatures have been linked to:[116]

- Acute kidney impairment
- Chronic kidney disease
- Electrolyte imbalances
- Kidney stones
- Urinary tract infections
- Medication considerations (particularly diuretics, b-blockers, and angiotensin-converting enzyme inhibitors) - when patients are taking these medications, they are less able to thermoregulate and therefore cope with heat, and this increases the risk of heat related illness.

##### MECHANISMS

Hotter temperatures/heatwaves > dehydration > acute kidney injury, kidney stones
Recurrent extreme heat exposure > heat stress and dehydration > repeated subclinical acute kidney injury > chronic kidney disease > heatstroke > rhabdomyolysis and heat induced inflammatory injury to the kidney > acute kidney injury.



##### ACUTE KIDNEY INJURY

Extreme heat days increase insensible loss of body water and salt. This loss can lead to fluid deficit, vasoconstriction, reduced kidney perfusion and associated acute kidney injury. Extreme heat can also lead to heatstroke (both clinical and subclinical whole-body hyperthermia) which can lead to rhabdomyolysis or heat-induced inflammatory injury to the kidney and thereby acute kidney injury.[117] Multiple studies have shown increased hospitalisations for AKI during heatwaves.[21, 118-120]

During the severe European heat wave of 2003, acute kidney injury was a prominent cause of excess mortality.[121] Groups that are at particular risk of AKI are those with pre-existing renal disease, the elderly and/or those taking medications that impair renal perfusion (such as diuretics, b-blockers, and angiotensin-converting enzyme inhibitors).

##### CHRONIC KIDNEY DISEASE

There has been recent recognition of epidemics of CKD involving individuals exposed to recurrent extreme heat.[122] These have been observed in Central America, Sri Lanka, India, the Middle East, Africa, and North America.[123] Those affected are primarily young male laborers from rural communities required to perform strenuous work under very hot conditions (e.g. sugar cane harvesters). However, cases are also being observed in women and children. Typically, presentation is with asymptomatic increases in creatinine and minimal proteinuria, which commonly progresses to end-stage kidney disease.

Although the underlying cause of CKD in the above cases remains unproven, there is general agreement that heat stress and dehydration are likely to be key contributors. In terms of mechanism, heat stress, overexertion, and water shortage may lead to any combination of

#### KEY POINTS

- **Hotter weather due to climate change is directly related to both acute and chronic kidney injury from dehydration and insensible salt loss. This is particularly dangerous for individuals taking medications that impair renal function (diuretics, beta-blockers, NSAIDs and ACE-inhibitors).**
- **Dehydration predisposes people to nephrolithiasis, with renal calculi and dehydration both increasing incidence of recurrent UTI.**
- **Rhabdomyolysis and heat induced inflammatory injury to the kidneys has a direct toxic effect on nephrons leading to repeat acute kidney injury and loss of renal reserve, exacerbating CKD.**
- **Altered geographic distribution of malaria and dengue leads to an increased burden of infection and febrile illness which can be complicated by severe AKI associated with a high mortality rate (as high as 45%).**
- **Given that haemodialysis relies on the availability of large quantities of water for each treatment (500L per patient per treatment), water shortages and quality are impacting the provision of dialysis.**
- **Increasing health practitioners' understanding of how hotter temperatures impact patients with kidney disease is significant for developing acute heatwave management plans.**

rhabdomyolysis, hyperosmolarity, hyperthermia, and extracellular volume depletion. These processes can result in acute kidney injury via activation of the vasopressin, aldose reductase and fructokinase pathways, hypokalaemia-induced renal vasoconstriction, uricosuria and urate crystal formation, and/or a reduction in renal blood flow.[115]

In turn, repeated episodes of acute kidney injury can lead to chronic damage, which manifests on biopsy as chronic tubulointerstitial disease, secondary glomerulosclerosis and ischemia.[115] Significant mortality has already been observed in affected areas; the death toll has been estimated at greater than twenty thousand in Central America and chronic kidney disease is now the leading cause of death in Nicaragua and El Salvador.[122]

##### KIDNEY STONES AND URINARY TRACT INFECTIONS

Increased temperatures may also increase the risk of kidney stones. The primary mechanism is thought to be volume depletion, which leads to a compensatory reduction in urine volume with subsequent urinary supersaturation with stone-forming salts.[124] It is projected that with the current rate of temperature rise, the US will see an extra two million lifetime cases of nephrolithiasis by 2050.[125] Risk of urinary tract infections is also projected to increase due to both poor hydration habits and increased incidence of nephrolithiasis.[117]

#### 3. WATER SECURITY AND QUALITY

Water shortage is predicted to increase due to climate change.[1, 75] Poor availability of drinking water can lead to dehydration in susceptible populations, affecting a range of conditions including acute kidney injury, chronic kidney disease, kidney stones, and urinary tract infections. It can also increase the nephrotoxic effects of some drugs,[115, 117] such as NSAIDs or ACEi/ARBs as examples.

Given that haemodialysis relies on the availability of large quantities of water for each treatment (500L of water per patient per treatment), water shortage and quality can also impact the provision of dialysis. This is particularly prominent in rural and remote parts of Australia where baseline water infrastructure is less than metropolitan areas. This makes rural and remote haemodialysis patients significantly more vulnerable, especially during drought periods. Many rural dialysis patients are already being forced to migrate to urban towns in order to access treatment due to water shortages in rural areas.[126]

> “Many rural dialysis patients are already being forced to migrate to urban towns in order to access treatment due to water shortages in rural areas.”

Flooding is also expected to increase due to climate change. Major flooding events increase the risk of risk of diarrhoeal illnesses and rodent-borne infections, such as leptospirosis and hantavirus, which are major causes of AKI in low-income regions.[127]



#### 4. FOOD SECURITY AND MALNUTRITION

An increase in extreme weather events causes a decrease in agricultural productivity and, therefore, decreased agricultural output. This in turn causes a drop in fresh food supplies, causing impacted populations to have an overreliance on unhealthy processed foods. This can increase risk of hypertension, diabetes and obesity, which in turn are all major risk factors for chronic kidney disease.

#### 5. CHANGE IN GEOGRAPHICAL DISTRIBUTION OF VECTOR-BORNE DISEASES

Mosquitoes (responsible for most vector-borne diseases) feed more frequently and produce more offspring during warmer temperatures.[115] Additionally, the parasites and viruses carried by mosquitoes also complete incubation faster within the female mosquito at higher temperatures, increasing the proportion of infective vectors. Because of this, climate is an important determinant of both the incidence and geographical distribution of vector borne diseases. This is further exacerbated by an association between forest loss and malaria prevalence rates across multiple countries.[128] Deforestation increases the incidents of malaria because it provides favourable conditions for the Anopheles mosquito. Pools of water being exposed to sunlight, the creation of artificial ditches and puddles and “tree bowls” (stumps left after logging) are more likely to pool less acidic water, which is conducive to Anopheles larvae development.[128]

Subsequent infection and acute febrile illnesses from vector-borne diseases are a major cause of acute kidney injury in tropical and sub-tropical regions.[115] Malaria and dengue are the two of the most important. In 1-5% of cases AKI

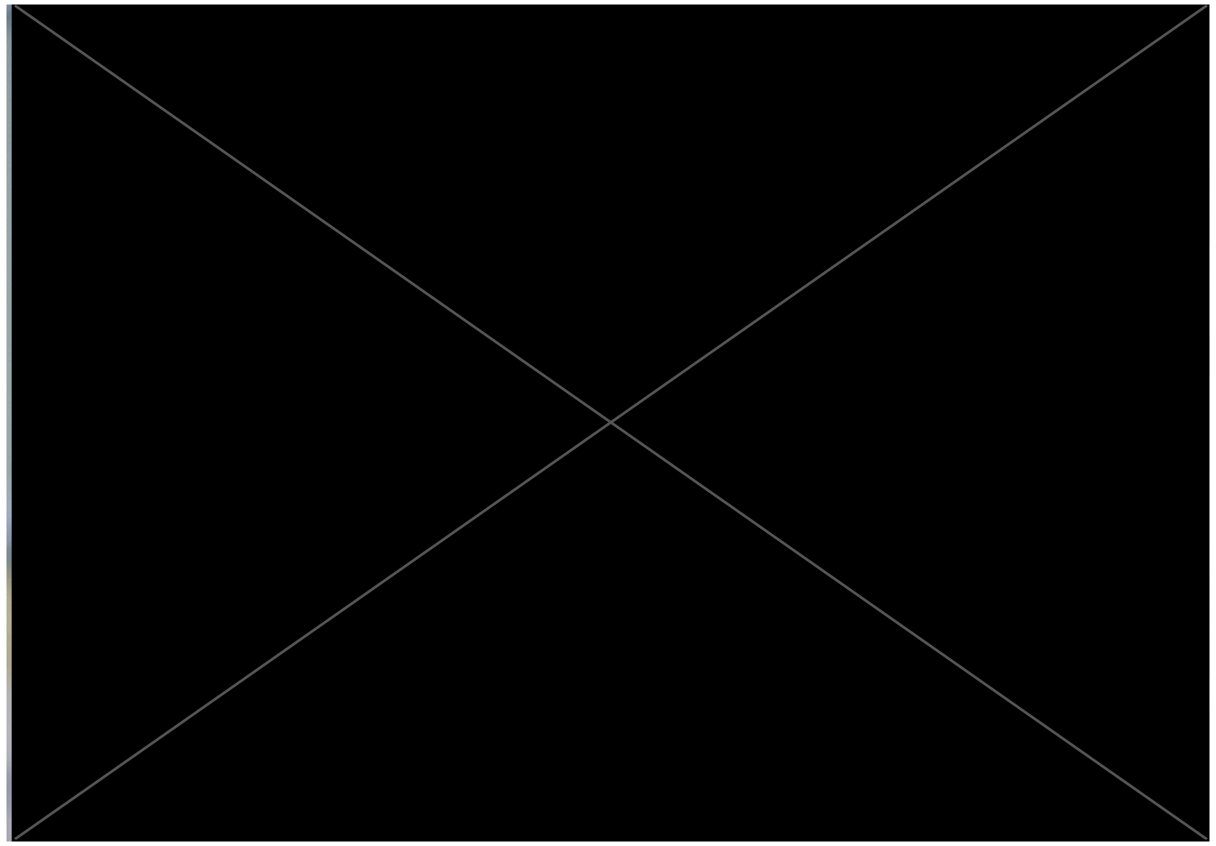

Chronic water shortages are endangering life-saving dialysis treatment for people in rural Australia.

complicates malaria, with incidence rising to 60% in those with severe disease.[129] When AKI does occur mortality rate can reach as high as 45%.[129] Data regarding AKI from dengue are more limited and heterogeneous, though in available studies rates of 11%-36% have been reported in those requiring hospitalisation, with mortality observed in 9-25% of cases.[130]

Global malaria burden is likely to be higher in the future than it would have been without climate change.[131] Rising temperatures have already favoured, and will continue to favour, dengue transmission.[131] In turn, increased rates of acute kidney injury is likely in affected regions.[132]

Zika is another mosquito-borne virus that may pose an increasing threat to humans due to climate change. Infection is generally asymptomatic or causes a non-severe febrile viral illness but can cause congenital abnormalities including microcephaly. Caution has recently been sounded for transplant recipients and other immunosuppressed populations (which includes those with CKD or on dialysis) travelling to affected areas.[133] Concern also exists regarding the potential risk of transmission of Zika virus through solid organ transplantation.

#### 6. VULNERABLE SHELTER AND HUMAN SETTLEMENTS AND MIGRATION

There has been an increase in the number of displaced people fleeing war and disasters in many parts of the world. Forcefully displaced populations are particularly vulnerable to renal disease. The multifactorial pathways include lack of access to healthcare, finance and nutritious food, including physical demands such as heat and extreme weather exposure and increased risk of infections. Further considerations relating to water security and quality see section 3.

#### RENAL HEALTH CO-BENEFITS OF GREENHOUSE GAS MITIGATION

Climate change mitigation strategies that can provide health co-benefits for patients at risk of, or with, kidney disease are:

Strategies that enable a shift from car travel to active transportation (e.g. walking or cycling) would not only reduce the substantial carbon emissions arising from the transport industry, but also air pollution and physical inactivity, which are both important risk factors for a broad range of chronic diseases including kidney diseases.[19, 134]

Initiatives that promote the adoption of more sustainable eating practices (namely, reduced consumption of animal products and increased consumption of vegetables, fruits, legumes, whole grains and nuts) would reduce the 30% of global emissions that is associated with agriculture, while at the same time reducing saturated fat intake and thereby the risks of obesity, diabetes, heart disease and nephrolithiasis.[135] Further, a balanced reduction in protein intake might slow CKD progression in patients with pre-existing kidney disease.[136]

##### DISEASE PREVENTION STRATEGIES

Increasing health practitioners' awareness and understanding of how hotter temperatures impact patients with kidney disease is significant for developing acute heatwave management plans.[21] This could involve education regarding adaptive strategies during heatwaves, such as avoiding exposure to heat, altering fluid restriction levels and altering medication dosage.



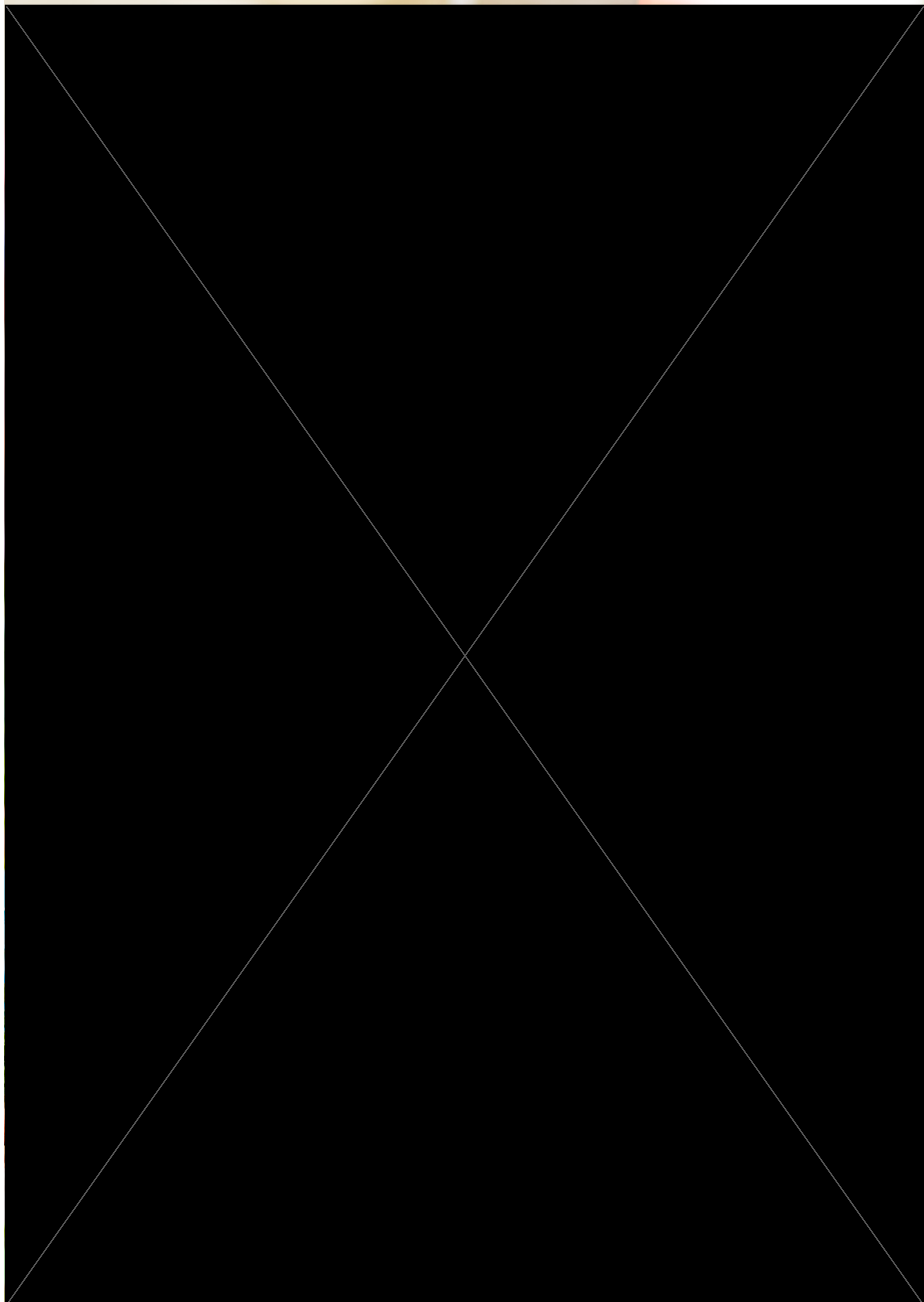

### Organ System Mapping Block: Gastrointestinal System

This block maps the relationships between climate change consequences and gastrointestinal disease. The gastrointestinal health benefits from mitigating greenhouse gases are also explored.

#### 1. INCREASED FREQUENCY OF EXTREME WEATHER EVENTS

Extreme weather events impact upon the gastrointestinal system by affecting food and water security and quality.[137-139] The health considerations include altered nutritional status of patients, micronutrient and electrolyte deficiencies, increasing incidence of food and water-borne gastroenteritis and possible increase in rates of liver disease due to altered diets and self-medicating drinking behaviour post-extreme weather disasters.

If climate change continues unabated, weather patterns will become more extreme and unpredictable, with average rainfall in southern Australia predicted to decline, the time spent in extreme drought conditions projected to increase, and water scarcity, heat stress and increased climatic variability in our most productive agricultural regions, such as the Murray Darling Basin, threatening our food security, economy, and dependent industries and communities.[72, 139]

The consequences of environmental change related to food and water security are explored in sections 2, 3 and 4.

##### LIVER DISEASE

The psychological, physical and economic costs of extreme weather resulting from the loss of human lives, injuries and destruction of properties, for the affected communities are significant. Psychological distress, depression, anxiety, post-traumatic stress disorder (PTSD) and increased alcohol consumption are most commonly reported in the aftermath of natural disasters, particularly bushfires and flooding events in Australia.[140, 141] In some cases, these mental health impacts can be long-lasting, with an Australian Ash Wednesday bushfire survey finding that 42% of participants classified for depression, anxiety or PTSD 1 year after the event, and 23% of participants at 20 months after the event.[140] Similarly, after the 2013 bushfires in the Blue Mountains, almost half of the 189 community members surveyed reported probable PTSD (45%), while 23% reported psychological distress and 16% reported heavy drinking, with men drinking significantly more than women.[140]

The use of alcohol in the aftermath of disasters is frequent and increases as the severity of exposure to traumatic events increases.[141, 142] As far as three-to-four years after the 2009 Black Saturday bushfires in Victoria, people in high, medium, and low-affected fire communities were reporting elevated rates of heavy drinking of 24.7%, 18.7%, 19.6% higher than normal, respectively.[142]

Individual stressors such as job loss, financial strain, the death of a loved one, and relationship breakdown during extreme weather or in the aftermath period are important factors that may also contribute to an increase in a person's alcohol consumption.[141] There are also potential gender differences in responding to bushfires which may impact on how men and women

#### KEY POINTS

- **Climate change is threatening global and local Australian food and water security, contributing to long-term malnutrition, deficiencies in dietary fibre, and nutrient deficiency (e.g. Vit-D, iron, folate, B-12).**
- **The use of alcohol in the aftermath and many years after bushfires is common and increases in use as the severity of exposure to traumatic events increases.**
- **Rising temperatures are consistently associated with increased rates of gastroenteritis and food spoilage.**
- **Increasing frequency and severity of flooding contaminates fresh water sources with pathogens causing diarrhoeal illness (norovirus, rotavirus, adenovirus, campylobacter jejuni and E.coli).**

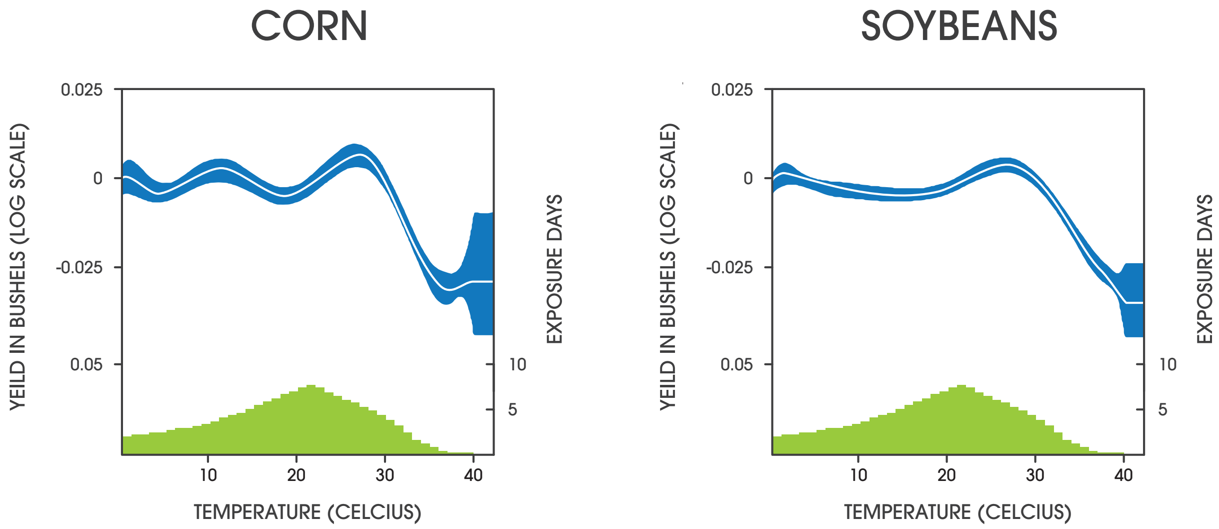


Corn and soybean crops, two of the world's largest sources of caloric energy, are significantly affected by more frequent hotter weather. Temperatures above 29 degrees for corn and 30 degrees celsius for soybean are incredibly harmful for crop yields, with forecast 30-46% decrease in yields by end of the century under the slowest warming scenario. Adapted from Schlenker W, Roberts MJ. Nonlinear temperature effects indicate severe damages to U.S. crop yields under climate change. PNAS 2009; 106: 15594-15598.

#### HOW DO GREENHOUSE GAS EMMISSIONS AFFECT FOOD SECURITY AND NUTRITION?

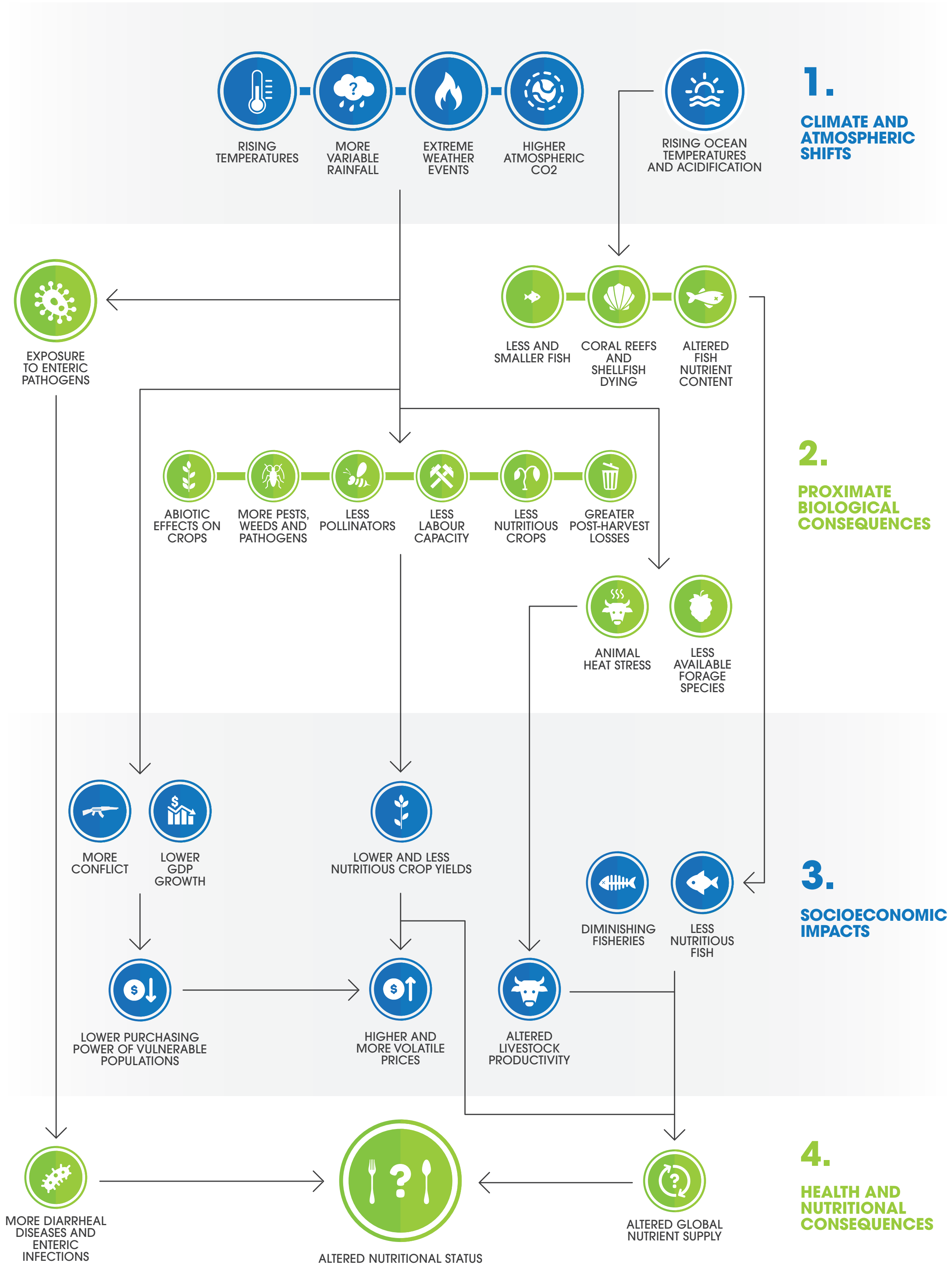


Pathways for impacts of climate change on food systems, food security, and undernutrition. Clinicians may consider the consequences for wound healing, infection immunity, or on post-surgical recovery. Authors own based on Myers et al. 2017.





cope in the aftermath of fires. For instance, a review on bushfires in Australia indicated that men may be more likely to stay and fight the fires as leaving and evacuating may be regarded as 'weak' or 'cowardly', whereas women may be more prone to evacuate early with their children.[143] As a result, the research suggests that this may affect subsequent coping behaviours, with women more likely to experience elevated rates of PTSD and general anxiety, and men more likely to report an increase in alcohol abuse and risk taking behaviours post-fires.[143]

Consequently, increasing frequency and severity of bushfires and flooding across Australia poses significant risk for increasing prevalence and incidence of alcohol-related liver disease. Between 10-20% of heavy drinkers will develop cirrhosis (usually after 10 years or more of drinking), with significant implications for patients and clinicians, such as management of cirrhotic liver and loss of function, bleeding and clotting risk management, nutrition, bleeding oesophageal varices and portal hypertension. Liver disease also presents challenges to other specialties whom operate or prescribe medications such as analgesia or chemotherapy that are processed in the liver.

#### 2. HEAT: HEATWAVES, OZONE AND AEROALLERGENS

Hotter temperatures are consistently associated with gastroenteritis and food spoilage. Hotter temperatures simply enable more rapid bacterial replication. Studies in South Australia, Queensland, Western Australia, New South Wales, Victoria and also in the United States have shown that increasing temperature and heatwaves significantly increase the incidence of Salmonella infections.[144, 145]

Globally, gastroenteritis is the most common cause of death in the 0-5 year age group.[137] An Australia-wide study demonstrated a 2.5–6% increase in the risk of foodborne illness for every degree of temperature rise[146] and an 11% increase in the number of emergency department visits in Sydney due to gastrointestinal infections amongst children under six.[147]

Temperature and ozone directly influence global crop yields. For example, corn and soybean, which are two of the four largest sources of caloric energy produced globally, significantly decrease in yield at temperatures above 29°C and 30°C respectively.[148]

Higher levels of ground level ozone have suppressed maize, wheat and soybean yields and have the potential to affect other crops such as wheat.[72]

Heat stress is a significant determinant of meat and dairy industry productivity. Higher temperatures have been shown to reduce milk yield by 10-25% and up to 40% in extreme heatwave conditions, and negatively affect production in the cattle, pig and poultry meat industries.[72] These issues are further exacerbated by drought.[72]

#### 3. WATER SECURITY AND QUALITY

##### CONTAMINATION OF WATER

A warmer atmosphere can hold more water vapour, contributing to an increase in heavy rainfall events and an increased risk of flash flooding. Worsening extreme weather events will increase the risk of damage to infrastructure for drinking water, wastewater, and stormwater, especially in areas with aging infrastructure. Contamination of freshwater drinking supplies and food sources is a key risk factor for gastrointestinal illness with either human or animal faecal pathogens.[150] Additionally, flooding of landfill sites and industrial areas poses the risk of chemical leakage into water systems with potentially harmful human exposure.

Heavy rainfall and flooding, especially after prolonged dry periods, can result in increased run off from land, stirred up water sediment and contamination of waterways with faecal microorganisms.[149] Agricultural livestock manure and inorganic fertilisers (containing nitrogen and phosphorous in particular) are major sources of water contamination and promote the rapid growth of pathogens and harmful algae.[149]

Waterborne pathogens are estimated to cause 8.5% to 12% of acute gastrointestinal illness cases, affecting between 12 million and 19 million people annually.[149] Eight pathogens, which are all affected by climate, account for approximately 97% of all suspected waterborne illnesses: the enteric viruses norovirus, rotavirus, and adenovirus; the bacteria Campylobacter jejuni, E. coli O157:H7, and Salmonella enterica; and the protozoa Cryptosporidium and Giardia.[149] Cases of cryptosporidiosis and giardiasis in children aged one to nine years also show a peak onset in illness during the summer months.[149]

##### DROUGHT AND POOR CLEAN WATER SUPPLY

At the other extreme, as temperatures increase evaporation rates increase, and so soil moisture decreases. When soils dry out, the severity of droughts increases.

During periods of drought or low rainfall, reliance on untreated water sources and low river or dam flows lead to increased concentration of water-borne pathogens. These lead to greater risk of water-borne infections and vulnerability to diarrheal diseases.[151]

#### Foods Susceptible to Mycotoxin Infections

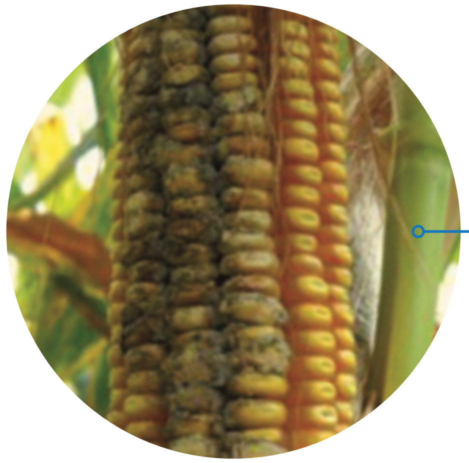

**Climate change will expand the geographical range where mould growth and mycotoxin production occur. Corn is especially susceptible to mould growth and mycotoxin production. Human dietary exposure to these toxins can result in sepsis and death. Aflatoxins (naturally occurring mycotoxins found in corn) are known carcinogens and can also cause impaired development in children, immune suppression, and, with severe exposure, death. Other crops susceptible to contamination by mycotoxins include peanuts, cereal grains, and fruit. In Australia, regulations are designed to prevent mycotoxins entering the food supply though may risk being overwhelmed in the future.**

Above: Authors own based on Crimmins et al. 2016
Opposite page: The percentage decrease in plant nutrition (zinc, iron and protein) amongst wheat, rice, maize and soybean crops as a consequence of increasing CO2 concentrations. Authors own with data from Myers et al. 2014.





#### Percent Change in Plant Nutrition between 380ppm $CO_2$ and 550ppm $CO_2$

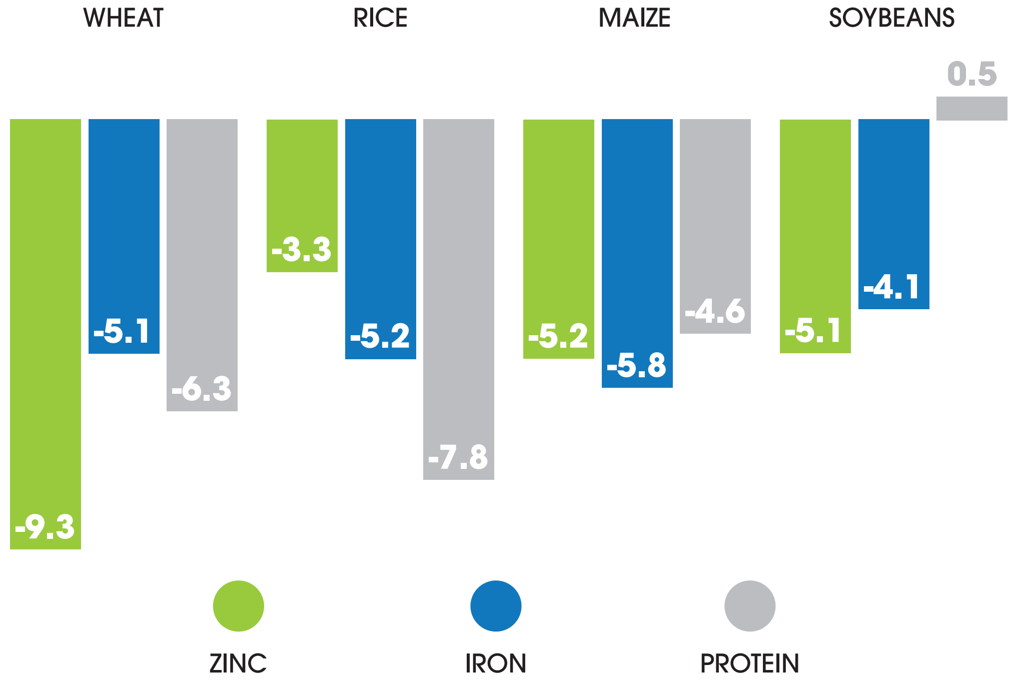


Children, older adults , pregnant women, and immunocompromised individuals have a higher risk of gastrointestinal illness and severe negative health outcomes from contact with contaminated water.[149] Pregnant women who develop severe gastrointestinal illness are at high risk of adverse pregnancy outcomes (pregnancy loss and preterm birth).

##### 4. FOOD SECURITY AND MALNUTRITION

Climate change is making weather patterns more extreme and unpredictable, with serious consequences for Australia's agricultural production, supply chain and food security. Having physical, social and economic access to sufficient, safe and nutritious food that meets an individual's dietary needs and food preferences is critical for an active and healthy life.

Threatened food security places Australians, and global populations, at increased risk of long-term macronutrient and micronutrient deficiencies, gastroenteritis, and gastrointestinal cancers, amongst other co-morbid diseases such as obesity or metabolic disease.

Climate change impacts on food security through:[72, 139, 152]

- Disruption of global and Australian food production systems through crop destruction
- Inhibiting access and supply during bushfire or flooding events

### Impacts of Rising $CO_2$ on the Nutritional Value of Crops

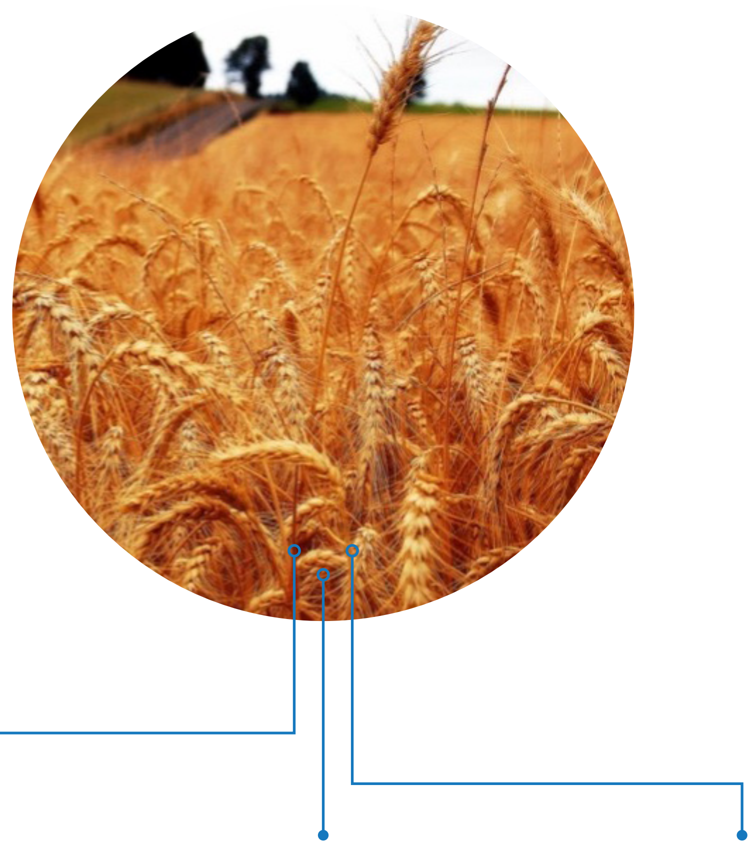

**MINERALS AND TRACE ELEMENTS**
Minerals and trace elements. Rising $CO_2$ levels are very likely to lower the concentrations of essential micro- and macro-elements such as iron, zinc, calcium, magnesium, copper, sulphur, phosphorous, and nitrogen in most plants (including major cereals and staple crops).

**RATIO OF MAJOR MACRONUTRIENTS (CARBOHYDRATE TO PROTEIN)**
It is very likely the rising $CO_2$ will alter the relative proportions of major macronutrients in many crops by increasing carbohydrate content (starch and sugars) while at the same time decreasing protein content. An increase in dietary carbohydrates-to-protein ration can have unhealthy effects on human metabolism and body mass.

**PROTEIN**
Protein content of major food crops is very likely to decline as atmospheric $CO_2$ concentrations increase to between 540 and 960 parts per million, the range projected by the end of this century. Current atmospheric concentrations of $CO_2$ are approximately 450ppm.

Authors own based on Crimmins et al. 2016.



- Creating niches that allow pests and weeds harmful to health to establish themselves
- Compromising crop defences through enhanced pest and pathogen growth and resistance
- Directly altering the nutrient profile, quality and seasonal availability of staple food crops
- Increasing food spoilage due to power outages or contamination due to poor clean water access
- Increasing food prices and promoting reliance on unhealthy processed food alternatives

#### DISRUPTION OF FRESH FOOD SUPPLY

Across Australia there is typically less than 30 days supply of non-perishable food and less than five days supply of perishable food in the supply chain at any one time.[139] Households generally hold approximately 3-5 days supply of food. Such low reserves are vulnerable to natural disasters and disruption to transport from extreme weather. For example, during the 2011 Queensland floods, several towns such as Rockhampton were cut off by floodwaters for up to two weeks, preventing food resupply.

Other examples include:

- Flooding in Townsville in 2019 that saw up to 300,000 cattle killed
- Cyclone Larry that destroyed 90% of the North Queensland banana crop in 2006, affecting supply for nine months and increasing prices by 500%
- Rainfall deficiencies in parts of Western Australia and central Queensland that reduced total national crop production by 12% in 2014/15
- Drought along the east coast of Australia has led to GrainCorp, Australia's largest bulk grain handler, producing grain at less than half the ten-year average and the lowest level in 50 years[153]

As crops and animal food sources become increasingly impacted by severe weather events, fresh foods will become increasingly scarce and expensive with consequences on health.[139] For example, food prices during the 2005- 2007 drought increased at twice the rate of the Consumer Price Index (CPI) with fresh fruit and vegetables the worst hit, increasing 43% and 33% respectively.[139] An increased reliance on processed foods and decreased consumption of fresh foods, particularly among lower socio-economic and remote populations, underpins long-term malnutrition, deficiencies in dietary fibre and various nutrient deficiencies, such as vitamin D, hypocalcaemia, hypokalaemia, iron-deficiency, folate and vitamin B-12 deficient anaemias. In the longer term, estimates on the burden of gastrointestinal cancers attributable to low fruit and vegetable intake range from 5% to 30%,[154-157] with mouth, pharyngeal, laryngeal, oesophageal and stomach cancers most commonly associated.[22]

On an international scale, the shift in recent decades to a more global food market has resulted in a greater dependency on vast food transport and distribution. Consequently, interruption to global food distribution and transport is having significant impacts not only on safety and quality but also on food access for Australians. The effects of climate change on global food supply will differ across Australia based on geographic, social, and economic factors.[158]

#### PEST AND PATHOGEN GROWTH AND RESISTANCE

Increasing temperatures allow for the survival of insect pests over winter (whose growth is otherwise controlled over the winter period) and drive shifts in the geographical range of crop pests and pathogens. In today's environment, insects, pathogens, fungi, and weeds are already estimated to be responsible for reducing the production of major crops by 25–40%.[72] Annual losses due to fungal infestation alone are estimated to reduce global dietary energy availability by 8.5%.[72]

As the environment changes to allow crop pests and pathogens to grow, the increased pest pressure on yields, combined with reductions in the efficacy of pesticides due to use, are likely to result in even greater pesticide use within agriculture. This increases the risk of pesticides entering the food chain.[159] Increased exposure to pesticides could have implications on the safety, distribution, and consumption of livestock and aquaculture products, ultimately with negative consequences for human health.[160]

Worsening drought has been shown to increase crop pests that can further decrease crop yields. For example, drought increases the spread of Aspergillus flavus mould that produces aflatoxin, a substance that contributes to the development of liver cancer in people who eat contaminated corn and nuts.[161]

##### FOOD SPOILAGE

The risk for food spoilage and contamination in storage facilities, supermarkets, and homes is likely to increase due to the impacts of extreme weather events, particularly those that result in power outages which may expose food to temperatures inadequate for storage.[160] In the United States, between 2002 and 2012, extreme weather caused 58% of power outage events. Power outages are often linked to an increase in illness with significant increase in diarrheal illness from consumption of spoiled foods due to lost refrigeration capabilities.[162]

##### NUTRIENT AND MICRONUTRIENT DEFICIENCIES

Although rising carbon dioxide levels stimulates plant growth and carbohydrate (starch and sugar) production, it reduces the nutritional value (protein and minerals) of most food crops.[160, 163, 164] This direct effect of rising $CO_2$ on the nutritional value of crops, in addition to increasing ground ozone levels, represents a potential threat to human health.[72, 160, 163-166]

These nutritional impacts include lower concentrations of protein. As $CO_2$ increases, plants need less protein for photosynthesis, resulting in an overall decline in protein concentration in plant tissues.[163, 164] When grown at the $CO_2$ levels projected for 2100 (540–958 ppm), major food crops, such as barley, wheat, rice, and potato, will exhibit 6% to 15% lower protein concentrations relative to ambient levels in 2015.[163, 164]

Increased $CO_2$ is also very likely to deplete other elements essential to human health (such as calcium, copper, iron, magnesium, and zinc) by 5% to 10% in most plants.[160] The projected decline in mineral concentrations has been attributed to two distinct effects of $CO_2$. First, rising $CO_2$ increases carbohydrate accumulation in plant tissues, which can, in turn, dilute the content of other nutrients, including minerals. Second, high $CO_2$ concentrations reduce plant demands for water, resulting in fewer nutrients being drawn into plant roots.[160] Globally, it is estimated an additional 175 million people will be zinc deficient and an additional 122 million people will be protein deficient by 2050.[72]

As people search for dwindling food resources, the consequence on already strained ocean food sources will increase. Overfishing, in combination with ocean acidification and the collapse of coral reef ecosystems put at risk the fisheries that supply a major component of the diet to hundreds of millions of people worldwide. It is estimated that 90% of global fisheries are currently fully or partially overexploited and at risk of being entirely depleted by 2050.[167] The loss of these fisheries would put over one billion people at risk of nutrient deficiencies.[72]





### Farm to Table: How Climate Change Affects Food Safety and Nutrition

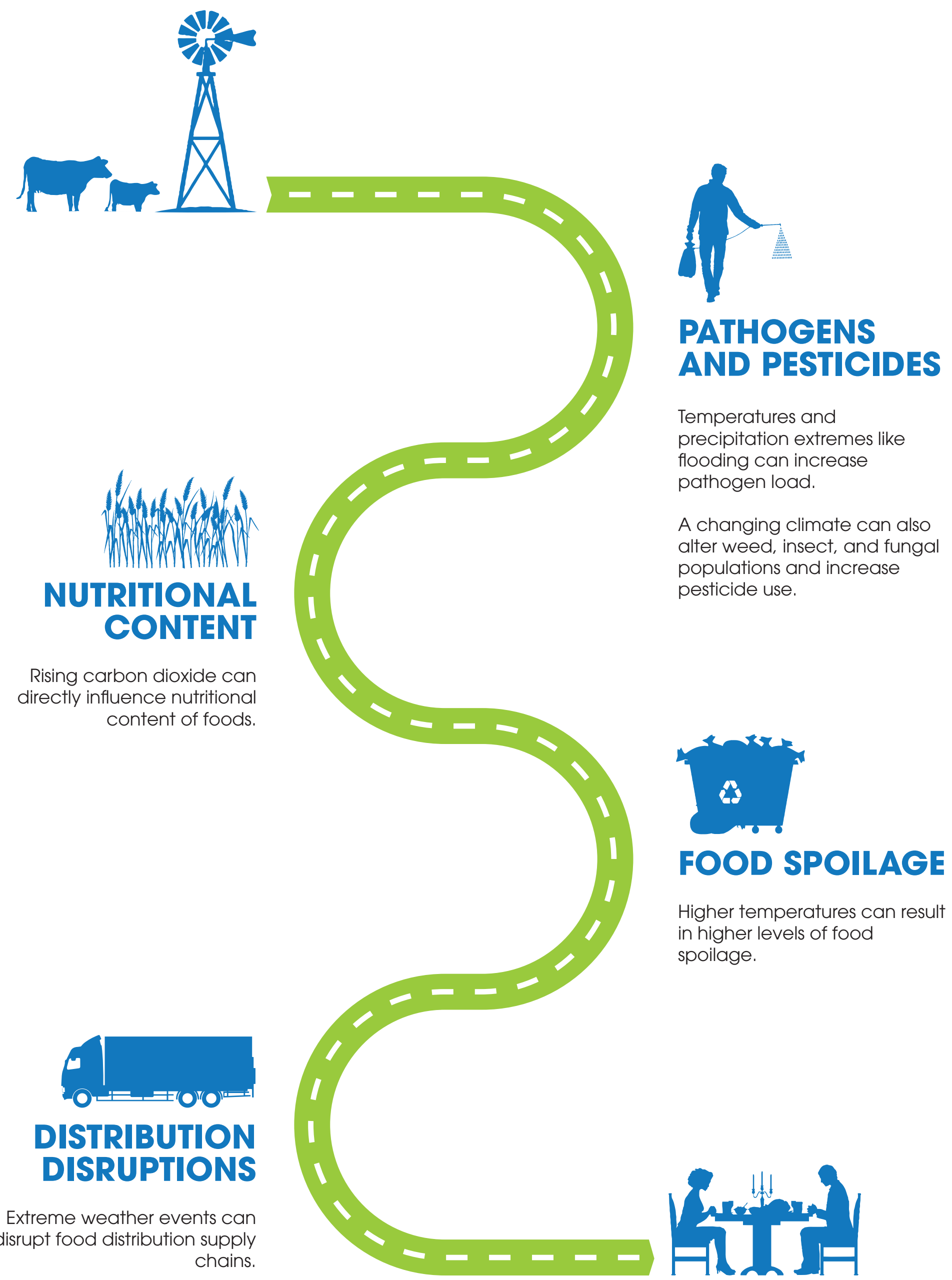


Farm to table describes how the pathways from production to consumption of food involve a network of interactions with our physical and biological environments. Rising CO2 and climate change will affect the quality and distribution of food, with subsequent effects on food safety and nutrition. Authors own based on Crimmins et al. 2016.

INCREASED CARBOHYDRATE INTAKE AND OBESITY

Elevated $CO_2$ and the subsequent increasing carbohydrate-to-protein ratio within staple food crops can adversely affect human metabolism and body composition.[160, 166, 168] The overall effect is an increase in the amount of carbohydrate consumed via plant food sources. This may contribute to increasing rates of overweight and obesity. Additionally, it is possible that people will increase total food and caloric intake in order to offset nutrient scarcity that will result from elevated levels of $CO_2$.[152] This may be further compounded by reduced availability and access to fresh food and vegetables, increasing reliance on the consumption of processed foods which are known to underpin obesity and metabolic syndrome.[169]

POPULATIONS OF CONCERN

Infants and young children, pregnant women, the elderly, low-income populations, agricultural workers, and those with weakened immune systems or who have underlying medical conditions are more susceptible to the effects of climate change on food safety, nutrition, and access. Children may be especially vulnerable because they eat more food by body weight than adults and do so during important stages of physical and mental growth and development. Children are also more susceptible to severe infection or complications from E. coli infections, such as haemolytic uremic syndrome.[160, 170]

Agricultural field workers, especially pesticide applicators, may experience increased exposure as demand for pesticide applications increase with rising pest loads. This could also lead to higher pesticide levels in the children of these field workers (through mechanisms including breast feeding and hand-to-mouth behaviours).[160, 171] People living in low-income urban areas, those with limited access to supermarkets,[160, 172, 173] and the elderly may have difficulty accessing safe and nutritious food after disruptions associated with extreme weather events. Climate change will also affect Indigenous peoples' access to both wild and cultivated traditional foods associated with their nutrition, cultural practices, local economies, and community health.[13, 174]

#### 5. CHANGE IN GEOGRAPHICAL DISTRIBUTION OF VECTOR BORNE DISEASES

The transmission of vector-borne diseases is altered as greenhouse gas concentrations rise. In Australia, increased average temperature, changing precipitation patterns, and a higher frequency of extreme weather events will influence the distribution, abundance, habitat availability, viral reproduction rates and prevalence of mosquitoes that carry infections, such as Ross River virus, Murray Valley encephalitis virus, Barmah Forest virus, Mycobacterium ulcerans and Dengue virus.[151]

In 2015 there were almost 10,000 reported cases of Ross River Virus in Australia. This was the largest annual number of cases ever reported, almost doubling the 2014 numbers, and 21% of cases occurring in South Eastern states.[175] Dengue fever vectorial capacity has been observed to increase 13.7% from the 1950s to 2016.[145] Dengue and other viruses can cause vomiting and diarrhoea in addition to mouth, nose and gastrointestinal bleeding.

#### 6. VULNERABLE SHELTER AND HUMAN SETTLEMENTS AND MIGRATION

Climate change is expected to drive increased scarcity of natural resources, such as food and water. This has the potential to exacerbate conflict and instigate mass migration, as the resources necessary to sustain life become scarce in populated regions. Those already experiencing vulnerability, social exclusion and marginalisation are likely to be most at risk. There is already evidence that children are





particularly vulnerable to the effects of climate change,[72, 176] and women are more vulnerable to the impacts of extreme weather events.[176] There is emerging evidence that climate change, particularly drought has played an important role over the past few decades in severely restricting food access during civil conflicts in sub-Saharan Africa and the Middle East.[72]

Overcrowding and lack of basic sanitary infrastructure including running water and toilets makes refugee and internally displaced person camps prone to infectious disease outbreaks. Waterborne and foodborne diseases, such as cholera, salmonella and trachoma, could become more prevalent as increased water scarcity and lack of food storage lead to increased reliance on contaminated food and water sources. Nutritional deficiencies are commonplace among those forced to move from their homes against their will.

Health risks increase in crowded shelter conditions following natural disasters, which suggests that some low-income groups living in crowded housing (particularly prevalent among low socioeconomic groups, migrants and rural and remote Aboriginal and Torres Strait Islander peoples) may face increased exposure risk to gastrointestinal illnesses and be more at risk from complications.[149]

#### GASTROINTESTINAL HEALTH CO-BENEFITS OF GREENHOUSE GAS MITIGATION

Consuming a plant-based diet can lead to a substantial reduction in greenhouse gas emissions, as well as significant benefits for individual and population health. Research published by Cancer Australia supports that diets rich in plant-based foods, such as fruit, vegetables and unprocessed cereals, reduces the risk of various gastrointestinal and other cancers.[22] Fruits and vegetables (particularly non-starchy vegetables) are thought to protect against mouth, pharyngeal, laryngeal, oesophageal and stomach cancers. Limited suggestive evidence supports a protective effect of non-starchy vegetables and fruits for nasopharyngeal and colorectal cancers. Such plant-based diets also protect against weight gain due to the low energy density of such foods.[22, 165]

The World Cancer Research Fund (WCRF) and American Institute for Cancer Research (AICR) have identified that there is convincing evidence that increased consumption of red meat and processed meat increases the risk of colorectal cancer, and limited evidence suggesting an increased risk of oesophageal, lung, pancreatic and stomach cancers.[22] The WCRF and AICR report recommends limiting consumption of red meat to less than 500 grams per week, with very little if any to be processed meat. Data on meat consumption indicate that Australian males and females eat around 200g and 120g respectively of meat, poultry and game each day.[22] Dose response meta-analyses have indicated a 17% increase in colorectal cancer risk for each 100 gram increase per day in red meat, and an 18% increase in colorectal cancer risk for each 50 grams increase per day in processed meat.

Cancer Australia recommends consuming adequate dietary fibre, including unprocessed cereals (grains) and pulses (legumes), and aiming for five servings of vegetables and two servings of fruit per day. Cancer Australia recommends limiting intake of red meat to less than 500 grams per week and avoiding processed meat to reduce cancer risk.[22] These recommendations are consistent with a diet that is significantly lower in greenhouse gas emissions, with red meat contributing up to 18% of global greenhouse gas footprint.

### Organ System Mapping Block: Neuroscience

This block maps the relationships between climate change consequences and neurological disease and mental illness. The neurological and mental health benefits from mitigating greenhouse gases are also explored. Limited literature search findings were available for sections 3, 4 and 5.

#### 1. INCREASED FREQUENCY OF EXTREME WEATHER EVENTS

Mental health conditions associated with experiencing an extreme weather or disaster event range from acute traumatic stress to more chronic stress-related conditions such as post-traumatic stress disorder (PTSD), complicated grief, depression, anxiety disorders, somatic complaints, poor concentration, sleep difficulties, sexual dysfunction, social avoidance, irritability and drug or alcohol abuse.[31, 177-179]

These mental health impacts can persist for years after the event and may have profound effects on the psychological functioning of individuals or populations.[31, 149]

Risk factors for developing mental illness in the aftermath of a disaster include:[180, 181]

- Magnitude of the traumatic event
- Exposure to the injury or death of a loved one
- Female gender
- Younger age
- Lower socio-economic status
- Lower education
- Minority or ethnic status
- Psychiatric history
- Family instability
- Inadequate social support

It has been estimated that 25% to 50% of people exposed to extreme weather events will experience negative mental health outcomes.[182] Typically, psychological responses are heightened in the first year after the disaster occurs and improve over time.[183] Common initial and immediate responses include hypervigilance, avoidance, anger, flashbacks, guilt, anxiety, emotionality, difficulty concentrating, rumination, preoccupation, and social withdrawal.[181, 184, 185]

PTSD prevalence ranges from 30% to 40% among those directly exposed to a natural disaster.[182] In a systematic review examining the mental health impacts of floods, a greater incidence of PTSD symptoms were observed among victims of extreme flooding compared to the general population.[186] PTSD symptoms are also reported among victims of bushfires.[187]

> "Direct stress from severe climate events predisposes individuals to acute stress disorders and chronic destabilising mental health conditions, particularly anxiety, depression and PTSD."

Slow-developing and prolonged extreme events, such as drought periods are one of the most well-documented climate hazards that can result in chronic psychological distress and a strong association with an increased risk of suicide.[188] A systematic review noted the most prominent causal pathway linking drought and mental health is via its gradual effects, its chronic nature and the perception that it is endless.[189] These were compounded by the economic impacts from land degradation and compromised food and water supplies, along with the sense of despair and isolation, particularly amongst those in rural areas and those dependent on the agricultural sector.[31, 190] Exemplifying this is that in New South Wales, approximately 9% of total deaths in 30-49 year old men are drought-related suicides.[191, 192]

The importance of Country to Aboriginal and Torres Strait Islander peoples involves a deep connection to the environment which provides

#### KEY POINTS

- **Direct stress from severe climate events predisposes individuals to acute stress disorders and chronic destabilising mental health conditions, particularly anxiety, depression and PTSD.**
- **Many psychiatric medications increase the risk of heat related illness: anticholinergic drugs impair sweating and predispose patients to hyperthermia; lithium toxicity is a risk in dehydration states; antipsychotics reduce thirst signals and increase risk of heat stroke through dopamine blockade; antidepressants have a direct effect on increasing core temperature, and sympathomimetic drugs cause cutaneous vasoconstriction and increase hyperthermia risk.**
- **Dehydration decreases central blood volume (hyper-viscosity) which predisposes at-risk individuals to arterial thrombosis and ischemic stroke. This is also augmented by worsening air pollution that triggers vascular dysfunction and atherosclerosis.**
- **The multi-organ-level influence of heat and dehydration predisposes elderly people in particular to higher rates of delirium – which carries associated morbidity and mortality.**
- **Solastalgia defines the way climate change creates a sense of loss, emotion¬al distress and existential anxiety about one's sense of future, place and identity.**

food security, economic opportunity and cultural and spiritual value.[193] Indigenous Australians are known to be more vulnerable to climate change effects due to pre-existing vulnerabilities resulting from historic and ongoing colonisation. The significant socioeconomic disadvantage experienced by some Aboriginal and Torres Strait Islander people places them at increased risk of suffering from environmental health risk factors.[194] For example, there is unequal access to primary health care, safe drinking water, effective sewerage systems, rubbish collection services and healthy housing, all of which are likely to be further undermined during acute rapid or slow-developing extreme weather events.

##### SOLASTALGIA AND ECO-ANXIETY

In recent years, the term eco-anxiety has been used to describe people's experiences when faced with the ecological and existential threats posed by climate change, and other environmental issues.[195]

'Solastalgia' describes the sense of loss people suffer when environmental degradation occurs in one's home and local environment; compromising one's sense of place and identity and often manifesting as hopelessness, anger, sadness and discomfort.[185] The term also defines the way climate change is negatively impacting on quality of life, creating emotional distress and existential anxiety about one's sense of future place and identity.[195-197]

Solastalgia was coined in Australia after observation of chronic drought conditions in New South Wales led to feelings of distress, loss, and bereavement, specifically among farmers, because of isolation and the loss of their livelihood.[197] Solastalgia may also be prominent amongst Aboriginal and Torres Strait Islander people and young people who are deeply concerned by the impacts of climate change.[177]

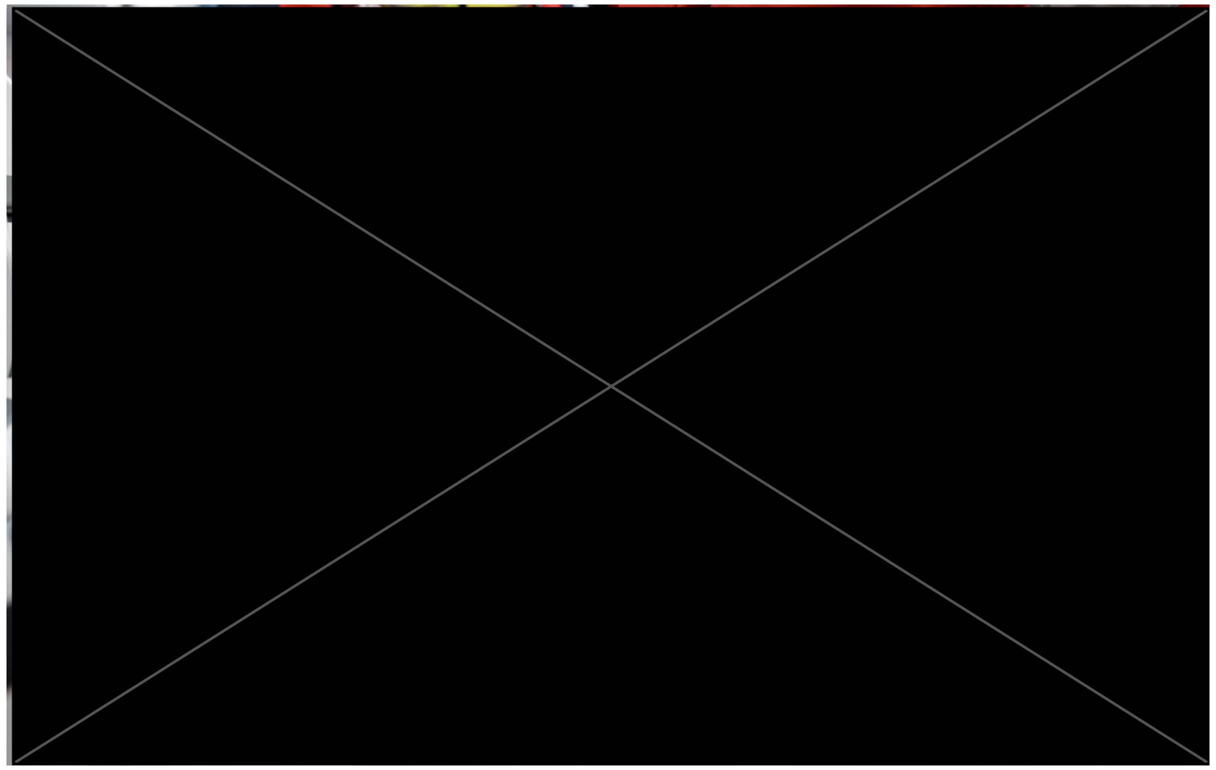

The elderly are highly vulnerable to climate change and more likely to experience multi-organ level impacts with considerable morbidity and mortality.



#### 2. HEAT: HEATWAVES, OZONE AND AEROALLERGENS

Increased temperatures are a well-established contributor to psychological distress.[31, 149, 185, 187, 188] Hotter temperatures are known to increase cortisol release, impair execution of effective behavioural responses, decrease the capacity of both working and short-term memory,[198] reduce sleep quality and disrupt people's physical activity routines.[199] This results in reduced wellbeing and increased psychological distress.[200, 201] In Adelaide, hospital admissions due to mental health have been found to increase by 7% during heatwave events,[202] with similar studies finding an almost 10% increase in admissions due to anxiety, mood (affective) disorders, stress-related and somatoform disorders. Additionally, hospital admissions increased by approximately 17% among people with dementia.[203]

> "The multi-organ-level influence of heat and dehydration predisposes elderly people in particular to higher rates of delirium – which carries associated morbidity and mortality."

The most vulnerable members of society - the very old, the very young, Aboriginal and Torres Strait Islander communities and those who work outdoors are also most vulnerable to exposure to extreme heat and are disproportionately represented amongst mental health statistics during these periods.[187] This is supported by national and international research demonstrating clear spikes in suicide rates during heatwave events, particularly amongst residents of warmer states and territories.[145, 204, 205] In Melbourne, a global average temperature increase of 1.5 degrees is projected to result in 12-17 additional days above 35 degrees per year.[206]

##### PSYCHIATRIC MEDICATION CONSIDERATIONS

Psychiatric medications increase people's vulnerability to heat-related morbidity.[149, 187, 207, 208] The thermoregulatory control centre of the brain, the hypothalamus, contains dopamine, noradrenaline, serotonin, and alpha-adrenergic receptors. Almost all psychotropic medications (except benzodiazepines) affect these neurotransmitters in some way.[207, 209]

For example:

- Anticholinergic drugs inhibit parasympathetic nerve impulses through selective blocking of acetylcholine to its receptor. Acetylcholine functions as the neurotransmitter at both cholinergic sympathetic nerves carrying signals to sweat glands, and at muscarinic receptors in the sweat glands. Consequently, drugs with anticholinergic effects impair sweating and reduce heat elimination, thereby increasing the vulnerability of their users in heatwaves.[208, 210]
- Those taking lithium are at increased risk for toxicity, because heat can lead to dehydration.[210]
- Antipsychotics, such as haloperidol, clozapine, risperidone and olanzapine, have both anticholinergic and central thermoregulatory effects through dopamine

blockade and their use is associated with increased risk of heat stroke.[211] Additionally, many common anti-psychotics have been reported to reduce thirst perception in patients.[207, 210]

- Antidepressants, such as dual dopamine/noradrenaline reuptake inhibitors, have been shown to significantly increase core temperature in exercising humans.[208]

- Sympathomimetic drugs, especially those that act as agonists at the adrenergic α receptor, cause hyperthermia through increased cutaneous vasoconstriction.[208] Some sympathomimetics also increase metabolic heat production by agitation-related expanded muscular activity.[208]

For many people living with dementia, their ability to undertake adaptive behaviours to heat can be impaired.[187] This includes recognition of the need to rehydrate or to remove layers of clothing.

Amongst those with pre-existing medical conditions, psychiatric illness was most closely associated with death during a heatwave.[185] In Wisconsin, United States, more than half of heat-related deaths resulting from a 2012 heat wave occurred in people with at least one mental illness - and half of these people were taking a medication that sensitises people to heat.[212]

##### CEREBROVASCULAR DISEASE

The association between environmental factors and acute risk factor for stroke is debated. Recent literature reviews have concluded that both heatwaves and cold exposures are associated with increased hospital admissions for cerebrovascular accidents.[59, 208, 213, 214]

Amongst the elderly, increased stroke risk occurs through various mechanisms, such as impaired ability to thermoregulate, premorbid conditions such as diabetes mellitus, and medications that modify blood pressure, circulation, and perspiration rates.[214] A decrease in the perception of warmth or cold can reduce the capacity of the elderly to identify when they are experiencing temperature signals and decrease their ability to adapt.[214] In response to heat, this pathway is augmented by increased dehydration, decreased central blood volume and reduced arterial pressure. Red blood cell count, blood viscosity, neutrophils and platelet counts increase with reduced blood volume.[40]

These factors predispose to a marked increase in arterial thrombosis, ischemia and increased mortality in hot weather.[40, 214] Conversely, cold exposure is also associated with increased induction of haemorrhagic and ischaemic stroke.[213-215]

##### NEURODEGENERATIVE DISEASES

Heat is known to trigger or exacerbate symptoms for people with pre-existing neurological conditions (e.g. multiple sclerosis).[216, 217] The elderly with co-existing age-related neurobiological and neuropsychiatric conditions are particularly vulnerable.

##### DELIRIUM

During the major Victorian heatwave of early 2009, emergency department admissions of people over 75 years of age increased by 37% in Victorian hospitals, compared to a 12% increase in other age groups.[60] The combination of dehydration, cardiovascular, renal, psychological distress and altered medication efficacy may all contribute to the increased incidence of delirium during hotter temperatures. Those with access to air conditioning may be able to avoid these effects, however the most marginal or those experiencing power outages or do not have access to air-conditioned facilities are at increased risk.



### Heat, Heat Stroke and Cerebrovascular Disorders

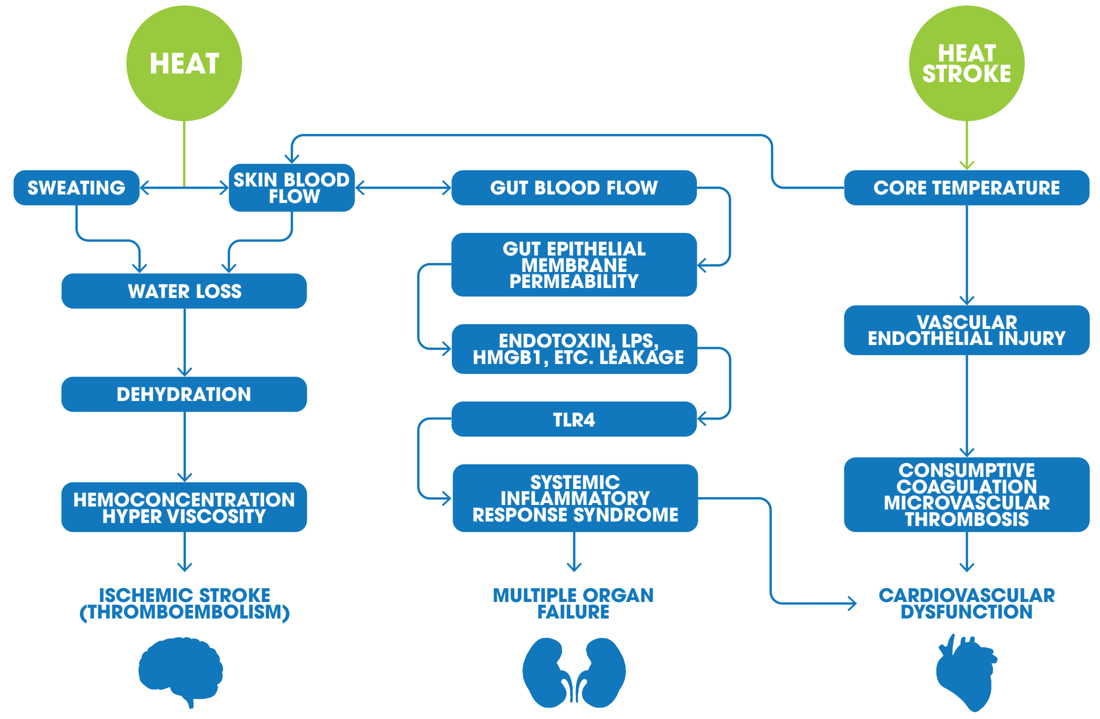


#### Proposed Mechanism

**1. Heat**
**In response to heat exposure, increased skin blood flow (SBF) and sweating lead to water loss and dehydration. The accompanied hemoconcentration and hyperviscosity may cause thromboembolism, leading to increased risk of ischemic stroke.**

**2. Heatstroke**
**In presence of heat stroke, increased core temperature redistributes blood flow to the skin to facilitate heat loss and limit hyperthermia.**

**3. Gut blood flow**
**Consequently, gut blood flow decreases with prolonged reduction increasing gut epithelial membrane permeability, allowing bacteria, its toxic cell wall component lipopolysaccharide (LPS), or HMGB1 (high mobility group box 1) to leak from the gut lumen into the systemic circulation. TLR4 (toll-like receptor 4) recognizes these molecules, stimulating the innate and adaptive immune systems and causing systemic inflammatory response syndrome (SIRS).**

**4. Multiple Organ Failure**
**In combination, the hyperthermia-impaired vascular endothelium induces occlusion of arterioles and capillaries (microvascular thrombosis) or excessive bleeding (consumptive coagulation), leading to multiorgan system failure, including cardiovascular dysfunction.**

Authors own based on Liu et. al 2015 and Lavados et al. 2018.

### Mechanisms for Cold and Cardiovascular Disease

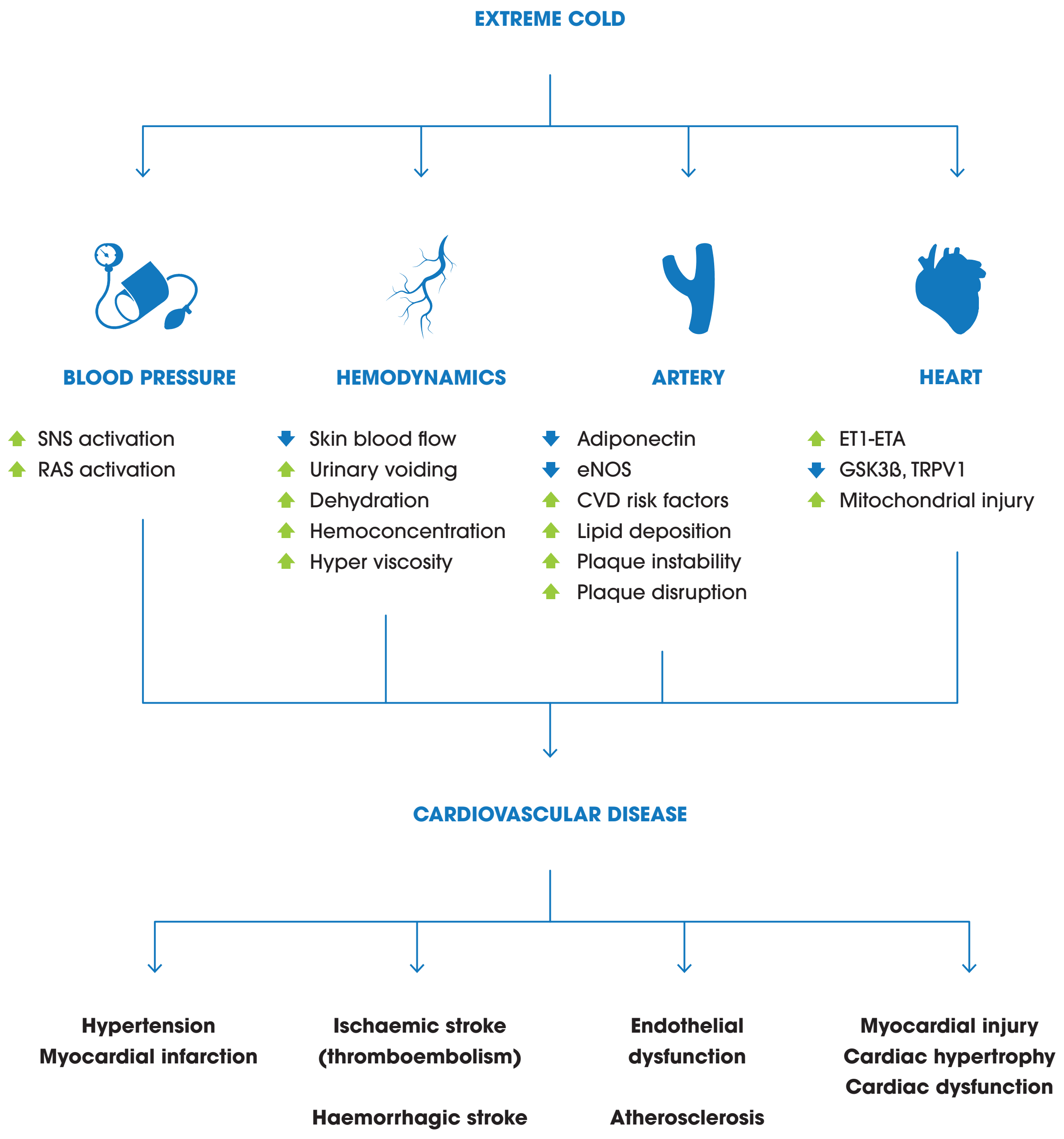


Authors own based on Liu et. al 2015 and Lavados et al. 2018.





#### 6. VULNERABLE SHELTER AND HUMAN SETTLEMENTS AND MIGRATION

Extreme weather events, sea-level rise, destruction of local economies, resource scarcity, and associated conflict due to climate change are predicted to displace millions of people worldwide over the coming century.[196] By causing or contributing to extreme weather events, climate change may result in geographic displacement of populations, damage to property, loss of loved ones and chronic stress, all of which can negatively affect mental health. Long-term drought has also been increasingly linked to conflict and forced migration, which can influence psychosocial outcomes like the propensity for stress, PTSD, anxiety, and trauma.

Particularly amongst Aboriginal and Torres Strait Islander peoples, the strong spiritual and cultural attachment to land predisposes to psychological impacts as a result of continued environmental degradation. In the Torres Strait, Indigenous peoples are subject to sea level rise that is flooding their homes and cultural sites, contributing to a loss of identity and sense of safety in their home.[218, 219]

#### NEUROLOGICAL AND MENTAL HEALTH CO-BENEFITS OF GREENHOUSE GAS MITIGATION

Increased exposure to natural and green space settings, particularly in urban areas has shown to confer multiple benefits for mental and neurological functioning.[31] Adaptation strategies that encourage and allow walkable cities and exercise, increases in green space and urban forestry could all contribute to reductions in neurological deficits.

The HUNT Cohort Study has shown that undertaking regular leisure-time exercise (for example via increasing active transport and reducing reliance on vehicle transport) is associated with reduced incidence of future depression.[28] The majority of this protective effect occurred at low levels of exercise and was observed regardless of intensity.

> "Natural and green space settings, particularly in urban areas, confers multiple benefits for mental and neurological functioning such as reducing air pollutants that are involved in development and progression of Alzheimer's and vascular dementia."

After adjustment for confounders, the population attributable fraction suggests that, assuming the relationship is causal, 12% of future cases of depression could have been prevented if all participants had engaged in at least one hour of physical activity each week. There is a clear co-benefit in reduction of greenhouse gas emissions from increasing exercise prescription. Health benefits may be further compounded by the reduction in air pollution, particularly measures for energy and transport that reduce neurotoxic air pollutants that are known factors in the development or progression of a range of neuro / psychological diseases including Alzheimers and vascular dementia.

### Organ System Mapping Block: Reproduction

This block maps the relationships between climate change consequences and reproductive disease. The reproductive health benefits from mitigating greenhouse gases are also explored.

#### 1. INCREASED FREQUENCY OF EXTREME WEATHER EVENTS

During climate-related disasters, women suffer disproportionate mortality,[220] and female survivors experience decreased life expectancy.[221] Women and girls are at a higher risk of physical, sexual, and domestic violence in the aftermath of disasters[222] and are at a higher risk for mood disorders and poor economic recovery.[223] These impacts are amplified when women have a lower socioeconomic status or belong to marginalised sectors of society.[222, 224, 225]

For women giving birth in the time period following disasters, there is an increased risk of complications, including preeclampsia, bleeding, and low-birthweight infants.[226] These consequences are compounded for women of all ages who are more likely to experience dietary deficiencies, leading to poor physical health and vulnerability to resource shortages ensuing from catastrophes.[227] Further, the risk of leaching from toxic waste sites into floodwaters during extreme weather events is significant and may have effects on development.[31] These include subtle changes such as small reductions in IQ from exposure to lead, changes in onset of puberty from exposure to endocrine disrupting chemicals, birth defects such as cleft palate due to dioxin-like compounds (pesticides and herbicides), and foetal loss through exposure-related spontaneous abortion.

These biophysical impacts on reproductive health are mediated by:[224]

- Poor access to obstetric care during and after disasters.
- Women are often homebound caring for children and elderly while waiting for relatives to return prior to evacuation.
- Poor, single, elderly women, adolescent girls, and women with disabilities are often at greatest risk for abuse because they have fewer personal, family, economic, and educational resources from which to draw protection, assistance, and support.
- Women suffer disproportionate job loss and stagnant personal economic recovery following disasters.





#### 2. HEAT - HEATWAVES, OZONE AND AEROALLERGENS

Increasing extreme heat impacts on women and pregnancy through increased risk of preterm delivery, congenital defects, gestational hypertension, and pre-eclampsia[224, 228-230]

Heat increases production of vasoactive substances, increases blood viscosity, and affects endothelial cell function, which may alter placental blood flow and increase propensity for hypertensive crises and stillbirth.[231] Hyperthermia is teratogenic, disrupting the normal sequence of gene activity during organogenesis.[230] These physiologic and biologic vulnerabilities are amplified by:[224]

- Poor access to healthcare and cooling facilities due to personal safety concerns and lack of access to personal transportation.
- Lack of communication and awareness of women's vulnerabilities to heat among local, national, and even global decision makers and healthcare personnel.
- Scarcity of gender-disaggregated heat-related health data, unknown critical exposure windows.
- Culturally prescribed heavy clothing garments.

Air pollution and increased ground-level ozone from elevated temperatures is associated with stillbirth, intrauterine growth restriction, and congenital defects.[232-234] These adverse reproductive outcomes occur due to air pollutants (e.g., CO, $PM_{2.5}$) crossing the placenta and impacting foetal growth during crucial developmental windows. Additionally, air pollutants exacerbate pre-existing maternal respiratory and cardiovascular health and result in reduced efficiency of placental function with consequent deterioration in foetal development.

##### KEY POINTS

- **Worsening extreme heat increases the risk of preterm delivery, congenital defects, gestational hypertension, and pre-eclampsia.**
- **Acute psychological distress from worsening extreme weather events increases excess foetal glucocorticoid exposure predisposing to intra-uterine growth restriction, preterm birth, low birthweight, and stillbirth.**
- **Increased air pollution from bushfires or vehicles crosses the placenta and damages DNA, impacting foetal growth and epigenetic outcomes during vulnerable periods.**
- **Heat stress and air pollution concentrations exacerbate underlying maternal cardio-respiratory conditions, reducing efficacy of placental function and uterine blood flow associated with preterm labour.**
- **For women exposed to increasing flooding due to climate change, water- and mosquito-borne diseases are particularly dangerous during pregnancy.**
- **Active transport (walking, cycling) and increasing intake of plant-based dietary options benefit maternal and neonatal health and ensure that the health of a child born today is not defined by a changing climate.**

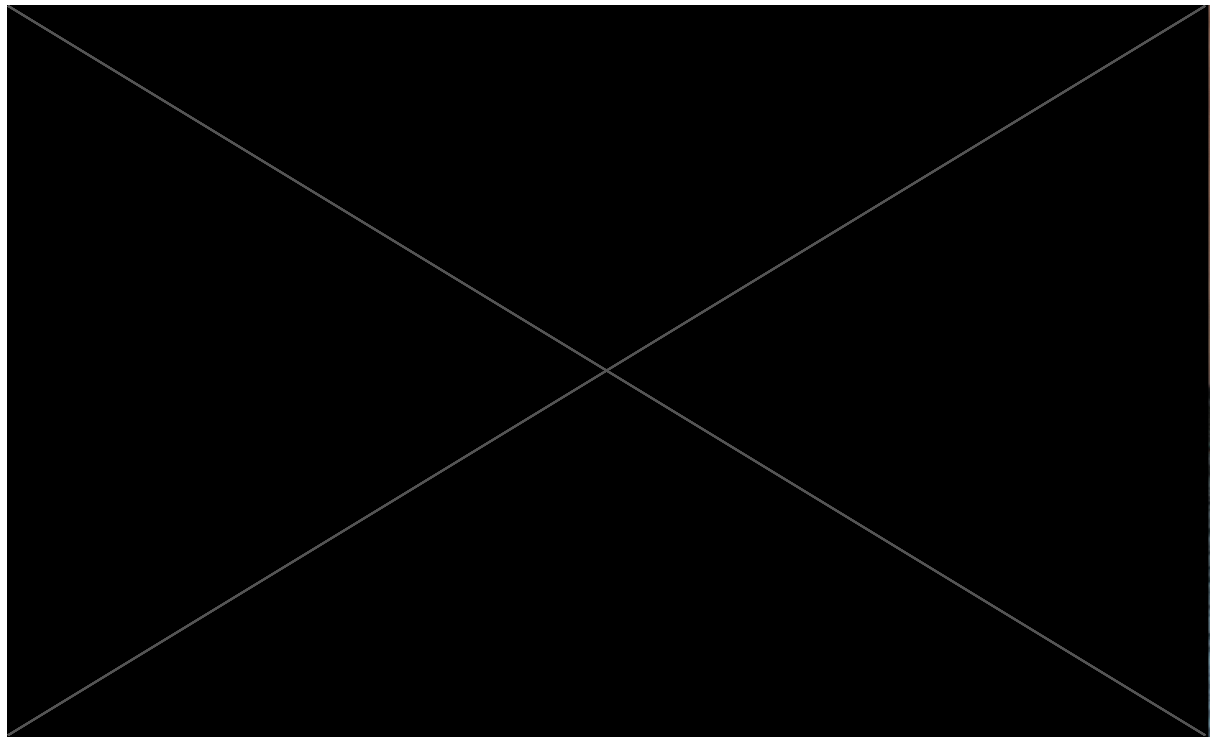

Climate change is impacting upon vulnerable periods during human development with profound consequences on every stage of the lifespan.

Vulnerable periods during human development include preconception (gametogenesis), preimplantation, the foetal period, and early childhood. Environmental exposures during these periods can lead to functional deficits and developmental changes through several mechanisms including genetic mutations and epigenetic change. Some chemicals damage DNA directly, causing mutations in gametes or the developing foetus that can lead to adult disease or conditions that increase disease risks such as obesity.[235]

In the United States a recent literature review covering 30 million births found a significant association in 58 of 68 studies between hotter temperatures, smog and air pollution and increased premature birth, low birthweight and stillbirths.[236]

#### 3. WATER SECURITY AND QUALITY

Shifting rainfall and increased rates of evaporation will lead to a lack of access to water and sanitation creating unsafe conditions for women, especially during reproductive times.[237] Dehydration and waterborne disease are key consequences. Water scarcity forces provision from water sources that may be biologically and toxicologically contaminated, resulting in increased risk of bacterial, viral, and protozoan infections as well as toxin exposure.[238] In most countries women will be forced to travel increasing distances to procure water resulting in increased exposure to heat and at increased risk for physical abuse and harm.[239]

- Dehydration in pregnancy results in decreased uterine blood flow and is associated with preterm labour.[228]
- Infection in pregnancy leads to poor maternal and neonatal outcomes.

These impacts are mediated by:

- Traditional household gender role of providing water for the family; water scarcity equates to more time spent harvesting water and less time spent on other activities of livelihood such as economic gain.
- In some regions, carrying water may use up to 85% of a woman's daily energy intake.[238]

A change in patterns and concentrations of contaminants entering the marine environment will also impact seafood species, many of which provide a major source of protein to global populations. Such contaminants, particularly metals such as mercury and lead that accumulate in fish and seafood, are a special concern for human developmental effects.

> "Acute psychological distress from worsening extreme weather events increases excess foetal glucocorticoid exposure predisposing to intra-uterine growth restriction, preterm birth, low birthweight, and stillbirth."

#### 4. FOOD SECURITY AND MALNUTRITION

Women are particularly sensitive to the effects of food insecurity and resulting nutritional deficiencies due to increased needs during menstruation, pregnancy, and nursing. Shifting rainfall and temperature patterns will impair crop, livestock, and fishery yields, contributing to food insecurity. Globally women already suffer from higher rates of macronutrient and micronutrient deficiencies. This includes higher rates of anaemia, which is associated with cognitive impairments including poor attention span, diminished working memory, emotional regulatory issues, and impaired sensory perception.[240]

Foodborne illness and food insecurity may further lead to malnutrition with negative effects on neonatal outcomes including intrauterine growth restriction and perinatal mortality.[31, 236, 241] These impacts will likely occur as a result of decreased food supplies, and exposure to toxic contaminants and biotoxins resulting from extreme weather events.

Undernutrition in pregnant women is a global cause of low birth weight and other poor birth outcomes that are associated with later developmental deficits. For example, maternal undernutrition may act on the developing foetus to program the risks for adverse health outcomes such as cardiovascular disease, obesity, and metabolic syndrome in adult life. In this way, changes in maternal nutrition and in utero exposure to certain chemicals or biotoxins due to climate change may impact the health of future generations through epigenetic changes before conception and during pregnancy.
In Australia, one in six children live in poverty[242] with increased risk of insufficient food resources and undernutrition that could be made worse by climate change. Therefore, climate change effects on food availability and nutritional content could have a marked, multigenerational effect on human development, resulting in a lifetime of suffering with significant societal costs in terms of resources, medical care, and lost productivity.

Other health impacts in vulnerable populations may be through the paradoxical increase in pesticide use for food production due to food insecurity and increases in harmful algal blooms both leading to harmful foetal exposure to chemicals and biotoxins.

#### 5. CHANGE IN GEOGRAPHICAL DISTRIBUTION OF VECTOR-BORNE DISEASES

Changes in temperature, precipitation, and ecology are altering the geographic distribution of vector-borne diseases that are known to cause harm to maternal and neonatal health. Pregnant women are exceptionally vulnerable to mosquito-borne illnesses including:

- Risk of severe malaria that is three times higher than that of nonpregnant women.[243]
- Zika virus carries devastating foetal impacts, including microcephaly, CNS malformations, and impaired cognitive development.[244]
- Dengue virus is associated with increased risk of caesarean delivery, eclampsia, and growth restriction.[245]

In Australia, dengue infections have historically been isolated events amongst international travellers or in Far North Queensland. However, cases of local infection are now being detected and may pose an increased risk to wider Australia as temperatures alter the geographic pattern of mosquito habitats.[246]

Pregnant women also have particular characteristics that make them and their foetus especially susceptible to mosquito-transmitted diseases:

- Pregnant women produce higher volumes of $CO_2$, a chemoattractant for mosquitos, and have increased peripheral blood flow, the heat from which allows mosquitos to locate hosts.[224]
- Hormonally induced changes in immunologic function during pregnancy lead to decreased immune response, which manifests as higher intensity of viremia and parasitemia.[247, 248]
- Infection during pregnancy can result in anaemia and diminished transplacental nutrient transport resulting in intrauterine growth restriction and increased vulnerability of the mother to haemorrhagic complications of delivery.[249]

Additional factors such as lack of access to prenatal obstetric care and assisted deliveries places women with infections at greater risk of postpartum haemorrhage and poor maternal outcomes, including death.[224]

> "Changes in temperature, precipitation and ecology are altering the geographic distribution of vector-borne diseases, such as dengue, that are known to cause harm to maternal and neonatal health."

#### 6. VULNERABLE SHELTER AND HUMAN SETTLEMENTS AND MIGRATION

Forced migration or "trapped" populations (people unable to move from high-risk areas) are a significant consequence of environmental change. In regard to reproductive health, there are key gender differences that are of significance for women and newborns. These include:[250]



- Forced migration is physiologically and mentally stressful, leading to poor maternal and neonatal health outcomes.
- Women whose partners travel frequently (for work) are at higher risk of HIV infection.
- Lack of basic sanitation and health services is a critical compound factor of health issues for refugees and migrants.

In areas where drought is affecting populations, women are more likely to be displaced as a result.[251] They are at greater risk for human trafficking throughout their migration and have fewer employment opportunities.[250] Therefore, women are often unable to migrate into economically viable and less environmentally vulnerable regions and are forced into less secure circumstances with repercussions on food availability, adequate safe areas for breast feeding or available healthcare for prenatal, antenatal or postnatal care.

Education regarding the gender-specific health threats of climate change is needed within public health, policy, medicine, and general education.[224] These gender-specific health impacts are important as they frame the patient at the centre of their diagnosis and management. Additionally, awareness of these factors may support clinicians considering additional management strategies or referral to services that can contribute to providing quality patient care.

#### REPRODUCTIVE HEALTH CO-BENEFITS OF GREENHOUSE GAS MITIGATION

Pregnancy presents a unique and important time for families to consider behaviours and lifestyle determinants that can promote the health of both mother and baby. Increasing active transport and increasing the proportion of fresh fruit, vegetables and legumes in a mother's diet are beneficial to maternal and foetal health whilst also reducing greenhouse gas emissions. The dual health and environmental wins directly benefit the long-term future for all children and might ensure that the health of a child born today is not defined by a changing climate.[176]

##### CO-BENEFITS OF ACTIVE TRANSPORT

Health professionals who care for pregnant women have the opportunity to promote the potential health benefits of exercise for mothers, their baby and its future. Physical activity in pregnancy has minimal risks and has been shown to benefit most women. Benefits of exercise in pregnancy include reduction in Caesarean section rates, appropriate maternal and foetal weight gain, reduced hypertension in pregnancy, shorter first stage of labour and improved management of gestational diabetes.[252, 253] Even medically minor concerns during pregnancy, such as low back pain experienced by up to 68.5% of pregnant women, are significantly reduced with any amount of land-based exercise.[253]

A meta-analysis of nine randomised controlled trials that included 2059 women with uncomplicated, singleton pregnancy with normal body mass index showed that women who were assigned randomly to 30-60 minutes of aerobic exercise three-to-four times per week had 49% lower incidences of gestational diabetes mellitus, 79% lower incidence of gestational hypertension disorders, 18% lower incidence of caesarean delivery, and a 9% higher rate of vaginal delivery.[254]

These findings are also supported amongst women who may not exercise regularly, with a recent metanalysis focussing on overweight or obese women that included nine randomised controlled trials and 1502 women and showed benefits of exercise in terms of a 38% lower rate of preterm birth and 39% lower rate of gestational diabetes mellitus.[255] There are also well-established benefits to maternal mental health, supporting reduced incidence and severity of postnatal depression and anxiety.[256]

Simply, there is no intervention health providers can recommend to pregnant women as impressive as exercise in its impact on maternal and perinatal outcomes.

Exercise may decrease the incidence of gestational diabetes mellitus by attenuating the increase in insulin resistance that is associated with pregnancy.[255] It may decrease the risk of gestational hypertension disorders by reducing oxidative stress and therefore improving endothelial function.[257] Exercise may decrease the incidence of caesarean delivery (and importantly increase the incidence of vaginal delivery) by improving maternal physical fitness and lowering birthweight.[255, 258]

Maternal exercise can be performed in manners other than by active transport, but for many women, including exercise in their daily commuting is the simplest and most efficient method. By reducing use of fossil fuel power vehicles, increased active travel can reduce greenhouse gas, fine particulate emissions whilst providing significant maternal and neonatal health benefits. Ideally, healthy women should get at least 150 minutes of moderate-intensity aerobic activity per week during pregnancy and the postpartum period.[253]

#### CO-BENEFITS OF PROMOTING PLANT-BASED DIETS

A well-planned plant-based diet can contribute to the mitigation of climate change and protect reproductive health. Animal products have much higher greenhouse gas emissions per gram of protein than plant-based alternatives. For example, beef and lamb have emissions per gram of protein that are about 250 times higher than legumes.[32]

Plant-based diets have increased worldwide in recent decades for numerous reasons, including improved knowledge that they might prevent coronary heart disease, cancer, and type 2 diabetes and for environmental reasons.

In regard to maternal and reproductive health, appropriately planned plant-based diets (fresh fruits, vegetables, nuts and legumes) are healthy and nutritionally adequate for all stages of life including pregnancy, breastfeeding and early childhood. Although mothers consuming plant-based diets must appreciate the risks of nutritional deficiencies such as proteins, iron, vitamin D, calcium, iodine, omega-3, and vitamin B12, the available evidence shows that well planned plant-based diets may be considered safe during pregnancy and lactation, but they require a strong awareness for a balanced intake of key nutrients.[259]

According to the developmental origins of adult disease (Barker) hypothesis,[260] extra-uterine environmental factors and maternal nutritional stimuli during pregnancy are major determinants for the risk of offspring developing chronic disease in adulthood. Therefore, maternal diets that are enriched with fruit, vegetables, and fibre may reduce foetal physiological adaptations that program for chronic adult disease. These benefits include reduced risk of coronary heart disease, obesity, improved lipid profile, and lower blood pressure.[261-263] Additionally, there is association with prevention of cancer (skin, prostate, breast, lung, and liver) and type 2 diabetes.[264, 265]

### Organ Mapping System Block: Intersystem

Epidemiology, trends, and interactions of common co-morbid diseases with climate change in Australia.

| HEALTH CONDITION | CURRENT ESTIMATE | TRENDS | POSSIBLE INTERACTION WITH CLIMATE CHANGE |
|---|---|---|---|
| **Asthma** | Around 2.7 million Australians, or 11% of the total population, had asthma in 2017/18. | Asthma prevalence has increased over the last 10 years. | Worsening outdoor air quality due to increased pollen and particulate matter increases the frequency of acute exacerbations of asthma. |
| **Cardiovascular Disease** | Around 1 in 20 Australians had cardiovascular disease in 2017-18. In 2017, 27% of all deaths in Australia were attributable to cardiovascular disease. | Cardiovascular disease prevalence has remained fairly consistent over time despite improvements in healthcare. | Extreme heat days and bushfire smoke air pollution increase acute cardiovascular events and increase hospital admissions and mortality from cardiovascular disease. |
| **Chronic Obstructive Pulmonary Disease (COPD)** | 464,000 Australians aged 45 years or older have COPD, 2017/18 self-reported national health survey. | Mortality from COPD in Australia has decreased over time, which is partly attributable to a decline in smoking rates. | Extreme heat and air pollution contribute to an increased frequency of exacerbations of COPD, and an increase in respiratory hospital admissions. |
| **Dementia** | In 2018, approximately 436,000 Australians had dementia. | In 2028, an estimated 589,000 Australians will have dementia. Over 1 million Australians are predicted to have dementia by 2058. | In heatwaves, people with dementia are less able to undertake adaptive behaviours and are more likely to be hospitalised. |



| HEALTH CONDITION | CURRENT ESTIMATE | TRENDS | POSSIBLE INTERACTION WITH CLIMATE CHANGE |
|---|---|---|---|
| **Diabetes** | In 2017-18, 1.2 million Australians had diabetes mellitus. In 2017, diabetes caused 4,839 deaths, making it the seventh leading cause of death in Australia. | If there are no changes to the incidence of diabetes, it is estimated that up to 3 million Australians over the age of 25 are expected to have diabetes by 2025 and around 3.5 million by 2033. | Heat waves cause increased deaths related to diabetes in elderly people; individuals with diabetes are more vulnerable to extreme heat; those dependent on insulin are vulnerable in the event of healthcare infrastructure damage in extreme weather events. |
| **Mental Illness** | 20.1% of Australians had a mental or behavioural condition in 2017-18. 13.1% had an anxiety-related condition, and 10.4% had depression or feelings of depression. | Levels of mental illness in Australia appear to be increasing, with a rise of 2.6% in mental or behavioural conditions between 2014-15 and 2017-18.7 45% of Australians aged 16-85 will experience a mental disorder at some point in their life. | Extreme weather events can predispose to negative mental health impacts; climate change can cause environment-related anxiety; mental health hospital admissions increase during heatwaves; medications for mental illness can impair thermal regulation. |
| **Obesity** | 35.6% of Australians aged 18 years and over were found to be overweight, and 31.3% were found to be obese in 2017-18. This is an increase since 1995, from 56.2% of adults being overweight or obese to 67.0%. | Adult obesity prevalence is projected to increase to 35% by 2025, and severe obesity (BMI > 35) is projected to be 13% by 2025. | Obese individuals are more prone to experiencing heat disorders on hot days. |
| **Disability** | In 2016, 4.3 million Australians, or 18% of the population, had a disability. For 79% of people with disability, their main form of disability is physical. For 21%, their main form of disability is behavioural or mental. | Around 13% of people below 65 have some level of disability, increasing to 51% of those aged 65 and older. As we live longer, there will be more Australians living with disabilities. | Individuals with disabilities may find it more difficult to respond to extreme weather events requiring evacuation, or to attend to self-care needs such as temperature regulation and hydration. |

Sources: (266-271)

### Healthcare’s Ecological Footprint

The Australian healthcare sector contributes to the problem of climate change, responsible for approximately 7% of Australia’s national carbon footprint and a considerable amount of Australia’s environmental pollution. As such, many health professionals are unknowingly contributing toward some of the very diseases they seek to manage and treat. It is essential to reflect on the environmental sustainability of healthcare delivery for the benefit of our patients, both now and in the future. All aspects of healthcare delivery should align with our professional commitment to ‘*Do No Harm*’.

#### HEALTHCARE EMISSIONS AND POLLUTION

Internationally, healthcare contributes to 4.4% of all greenhouse gas emissions, 2.8% of PM, 3.4% of $NO_2$ and 3.6% of SO2 emissions.[272]

The Australian Medical Association, Doctors for the Environment Australia (and others) have called on the Australian healthcare sector to reduce its carbon emissions to net zero by 2040, with an interim emission target of 80% by 2030 to address this situation. These targets are consistent with the 1.5 °C Paris Agreement.[273, 274] The National Health Service (NHS) in the UK are already showing leadership setting a net zero emissions healthcare target for 2040.[275]

Australian healthcare accounts for approximately 7% of Australia’s total carbon footprint, a larger contributor to national emissions than the activity of a population similar to South Australia.[8] Of the 7% of CO2e emissions (from health care), hospitals are responsible for 44%, pharmaceuticals 19%, capital expenditure 8%, community and public health 6% and general practice 4%.[8] Beyond carbon emissions, healthcare is a significant contributor to waste products and natural resource consumption, threatening our present and future health. There are significant areas for change. For example, only 2.3% of all Australian public hospital energy use is from renewable sources (above the grid background renewable level).[276]

Recognising the carbon harm from healthcare is pertinent to every Australian clinician:[277, 278]

- During one average working day an individual anaesthetist, administering N2O or desflurane can contribute CO2 equivalent to more than 1,000km of car driving.
- Every day of the year operating room staff will deposit into landfill more than 1,000 tons of rubbish.



#### AREAS FOR IMPROVEMENT

By understanding what contributes to emissions, medical students have an opportunity to reflect on effective opportunities for sustainable practice and on areas for leadership and advocacy.

At an organisational level, embedding environmental sustainability into institutional mission, vision and values, in accordance with obtaining a strong executive level champion, are key strategies consistently employed by global leaders in healthcare sustainability.[279, 280] Additionally, advocating for sustainable models of care, increasing renewable electricity, reducing consumption and waste (i.e. Choosing Wisely) and improving design of healthcare infrastructure will further reduce health care’s footprint.[5]

#### Carbon Emissions from Healthcare

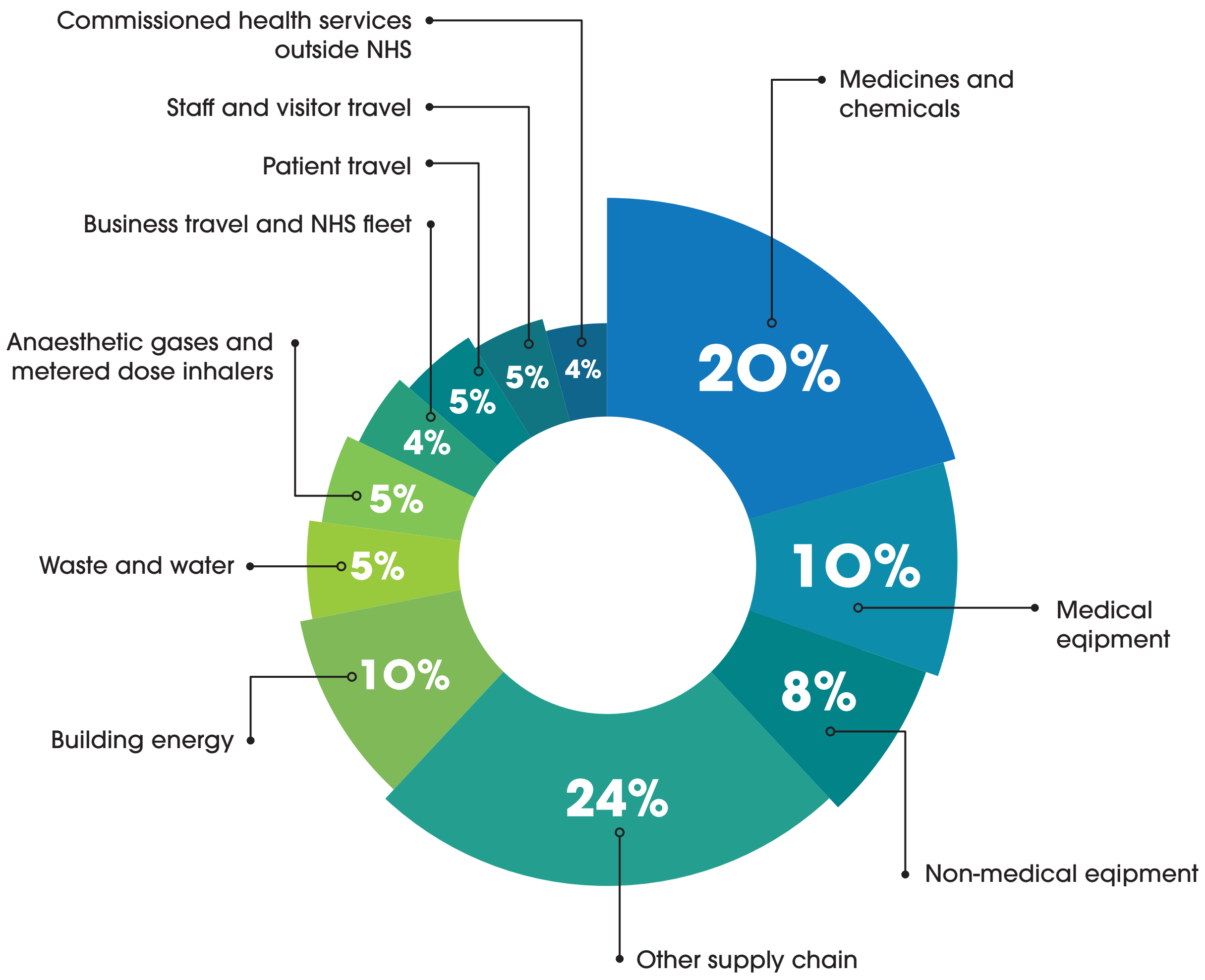


Authors own based on the National Health Service 2020.

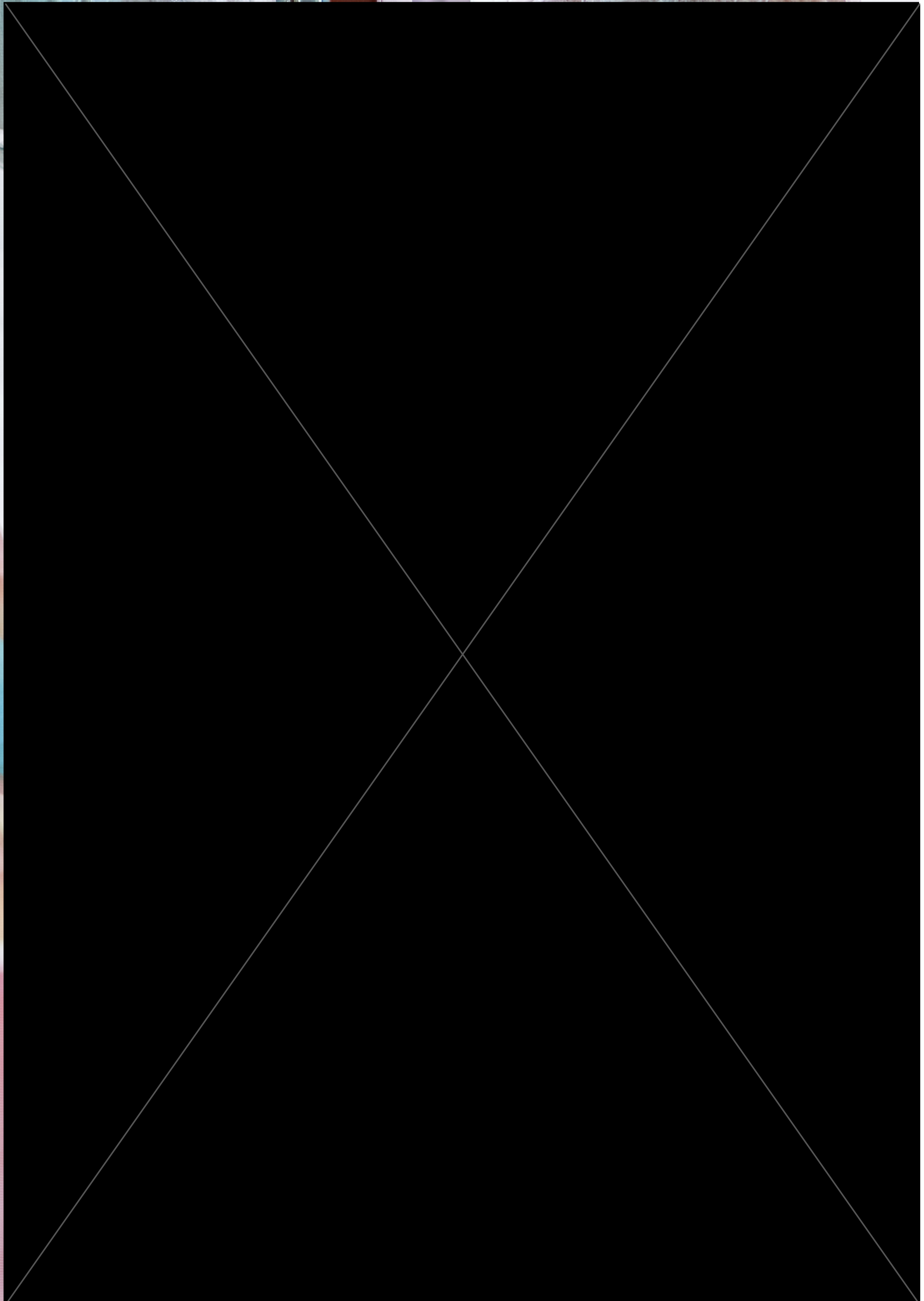

| | SOURCES OF EMISSIONS | MANAGEMENT SOLUTIONS |
|---|---|---|
| **Direct Energy Consumption** | Predominantly coal-fired energy used for:<br>• Lighting<br>• Heating/cooling<br>• Water heating<br>• Ventilation<br>• Powering medical and technological equipment<br>• Cooking and cleaning appliances | • Transition towards renewable energy sources (e.g. on-site solar power)<br>• Optimisation of energy use (e.g. energy-efficient lighting or reducing thermostat temperature in winter) |
| **Waste and Procurement** | • Disposal of biohazardous clinical waste (incineration, autoclaving, chemical disinfection)<br>• Single-use clinical equipment (including PPE)<br>• Excessive packaging<br>• Food waste<br>• Pharmaceuticals | • Reducing misclassification of biohazardous waste<br>• Transition towards reusable equipment with sterilisation<br>• Recycling programs<br>• Legislation that enables facilities to procure from sustainable suppliers |
| **Models of Care** | Inefficiencies and resource waste:<br>• Duplications in care provided<br>• Unnecessary diagnostic testing<br>• Unnecessary interventions and prescriptions<br>• Excess use of clinical testing | Ensuring:<br>• Integration and coordination between healthcare areas and providers<br>• Increasing diagnostic accuracy<br>• Implementing effective communication procedures<br>• Adoption of technological innovations (e.g. telehealth) |
| **Preventive Health Care** | Increasing health care utilisation (higher incidence/severity of chronic and infectious disease). | Public health initiatives (e.g. promoting active transport and plant-based food options). |
| **Infrastructure and Planning** | • Building, upgrading, and extending facilities<br>• Energy loss due to inefficiencies in infrastructure | • Preservation of local habitat<br>• Maximise natural light and ventilation<br>• Use of reflective materials to reduce need for electric cooling<br>• Using sustainable or recycled building materials<br>• Green spaces for exercise/health promotion<br>• Safe bicycle and pedestrian access |

Adapted from AMA Position Statement: Environmental Sustainability in Healthcare (5)

### The Role of Medical Students

Medical Students can have key roles in identifying, reducing and managing adverse health effects of climate change on Australians and the international community.

Within expected MD1 Principles of Practice Learning Outcomes and each of the domains of the MD Graduate Student Attributes, future doctors should be equipped with the knowledge, skills, values, competence and confidence they need to sustainably promote the health, human rights and wellbeing of current and future generations, while protecting the health of the planet.

#### MD1 PCP FINAL LEARNING OUTCOMES

- **LO6.** Demonstrate a sophisticated understanding of the determinants of health (physical, psychological, social, cultural, economic, environmental, gender) and their impact on health and health behaviours in individuals, communities and populations. CILO: 1, 3, 4, 6, 7, 8
- **LO7.** Recognise the impact of planetary health and climate change on human health at the individual, community and societal level. CILO: 5, 6
- **LO8.** Examine and expand on the core ethical, legal and moral principles which guide medical practice, with emphasis on consent, confidentiality, different communication media and advocacy. CILO: 4, 5, 6, 7, 9
- **LO18.** Identify and examine the suitability of a variety of resources, using these resources to determine the efficacy and effectiveness of current health care practices. CILO: 2,3,4,6,7,8
- **LO24.** Advocate for the advancement of the health and well-being of Australia's First Nations peoples. CILO: All (1-12)

#### MD2 GRADUATE STUDENT ATTRIBUTES DEVELOPED

Australian Medical Council accreditation dictates *40 Graduate Student Attributes*

##### DOMAIN: SELF

- **Attribute 2:** an understanding of the principles of reflective practice, the ability to apply them, and a recognition of their importance in health care.
- **Attribute 3:** an understanding of the principles of self-awareness .
- **Attribute 6:** the ability to manage uncertainty.

##### DOMAIN: KNOWLEDGE

- **Attribute 1:** an understanding of the scientific method relevant to biological, behavioural and social science.
- **Attribute 6:** an understanding of the factors that might disturb normal structure, function and development.
- **Attribute 7:** an understanding of the aetiology, pathology, symptoms and signs, natural history and prognosis of important physical and mental illnesses in all stages of life.
- **Attribute 8:** an understanding of the management (pharmacological, physical, nutritional, behavioural and psychological) of important medical conditions.
- **Attribute 9:** the ability to access new knowledge from all sources, to analyse and interpret it in a critical manner, and to apply it appropriately to their provision of health care.
- **Attribute 10:** the ability to learn from patients, health professionals and the community in a broad range of settings.
- **Attribute 11:** an appreciation of the responsibility to contribute towards the generation of new knowledge.

##### DOMAIN: PATIENT

- **Attribute 3:** the ability to advocate appropriately on behalf of the patient.
- **Attribute 4:** an understanding of factors affecting human relationships and the psychological, cultural and spiritual wellbeing of patients.
- **Attribute 7:** an understanding of chronic illness and disability and its impact on the patient, their carers and communities.
- **Attribute 11:** the ability to select and interpret the most appropriate and cost-effective diagnostic procedures.
- **Attribute 12:** the ability to formulate an evidence-based and cost-effective management plan in collaboration with the patient.
- **Attribute 14:** a recognition that it is not always in the interests of the patient to do everything that is technically possible to make a precise diagnosis or to attempt to modify the course of an illness.

> "Medical Students can have key roles in identifying, reducing and managing adverse health effects of climate change."

##### DOMAIN: MEDICAL PROFESSION

- **Attribute 2:** an understanding of the potential conflicts of interest that may confront doctors.
- **Attribute 3:** an understanding of and ability to apply the principles of ethics in the provision of health care and research.
- **Attribute 4:** an understanding of organisational governance, the ability to be an active participant in professional organisations, and an appreciation of the benefits of this participation.
- **Attribute 7:** an understanding of educational theory and practice and the ability to teach.

##### DOMAIN: SYSTEMS OF HEALTH CARE

- **Attribute 9:** an understanding of the structure of the Australian health care system and health care systems globally.
- **Attribute 10:** an understanding of the principles of efficient and equitable allocation and use of finite resources in health care systems, locally and globally.
- **Attribute 11:** an understanding of the role of political systems in shaping health care systems locally, nationally and internationally.

##### DOMAIN: SOCIETY

- **Attribute 1:** an understanding of the interactions between humans and their social and physical environment.
- **Attribute 2:** an understanding of the determinants of a well society and the economic, political, psychological, social and cultural factors that contribute to the development and persistence of health and illness.
- **Attribute 3:** an understanding of the principles of health promotion including primary and secondary prevention.
- **Attribute 4:** an understanding of the health of Indigenous Australians including their history, cultural development and the impact of colonisation and the ongoing health disparities of Indigenous people in this country and globally.
- **Attribute 5:** an understanding of the burden of disease in differing populations and geographic locations.
- **Attribute 9:** the ability to consider local, regional, national and global ramifications of health care issues.
- **Attribute 10:** the ability and a willingness to contribute to the community.
- **Attribute 11:** a commitment to contribute to the resolution of health inequities locally and globally.
- **Attribute 12.** an understanding of the relationship between environmental issues and the health of local communities and society.
- **Attribute 13:** a commitment to practise medicine in an environmentally responsible way.

### Opportunities for Applied Skills and Behaviours

Action by medical students as a trusted group of current and emerging leaders is important for modelling, education and facilitation of community-wide action.

##### 1. COMMUNICATION SKILLS AND THE DOCTOR–PATIENT RELATIONSHIP

- Identifying patients who are particularly vulnerable to heat, and ensuring that they take precautions and are monitored.[18]
- Ensuring that patients and the local community have access to and respond to public health advice, such as disaster and weather warnings from health departments and emergency services.[18]
- Recognising that climate change exacerbates health inequities – for example, through the unequal impacts of extreme weather events – and seeking opportunities to promote health and social equality.[17]

##### 2. APPLIED PROFESSIONAL KNOWLEDGE AND SKILLS

- Promoting urgent action to mitigate climate change through individual, practice-based, social and population-based initiatives.[281]
- Identifying co-benefits of action to reduce climate change in clinical consultations and hospital placement – for example, encouraging active transport, promoting low-energy diets including less meat and processed food consumption, preventing unwanted pregnancy,[282, 283] and promoting energy-efficient homes and buildings.[281]

##### 3. POPULATION HEALTH AND THE CONTEXT OF MEDICAL PRACTICE

- Understanding the response to the burden of non-communicable diseases that are the main cause of morbidity and mortality in Australia today, such as mental illness.
- Undertaking and supporting ongoing education for themselves, other health students and professionals, patients and the wider community about climate change and its impact on individual and population health.

> “Action by medical students as a trusted group of current and emerging leaders is important for modelling, education and facilitation of community-wide action.”

##### 4. PROFESSIONAL AND ETHICAL ROLE

- Taking personal action to mitigate climate change and improve health and equity. Examples of actions with both health and climate benefits include using active transport, minimising air travel, reducing highly processed food consumption, reducing meat consumption, and encouraging use of smaller cars driven less often.[281]
- Supporting community action – for example, community gardens for local food production, public open space for outdoor recreation and physical activity, safe walking and cycle ways, high-quality public transport systems.[281]
- Working with other professionals to strengthen individual, community and social action, through government, business and community organisations.
- Using medical expertise and the developing professional position as trusted community leaders to advocate on behalf of patients for effective climate change policy and action.[281]

##### 5. ORGANISATIONAL DIMENSIONS

- Investigating opportunities to reduce energy usage and other environmental impacts, minimising waste, improving efficiency, investing savings in further energy reductions[11] and addressing opportunities for new technologies within medical school and clinical school placements.[281, 284]



145. Zhang Y, Beggs PJ, Bambrick H, Berry HL, Linnenluecke MK, Trueck S, et al. The MJA–Lancet Countdown on health and climate change: Australian policy inaction threatens lives. Medical Journal of Australia. 2018;209(11):474-. Available from: https://onlinelibrary.wiley.com/doi/abs/10.5694/mja18.00789
146. Hall GV, Hanigan IC, Dear KBG, Vally H. The influence of weather on community gastroenteritis in Australia. Epidemiology and Infection. 2011;139(6):927-36. Available from: https://www.cambridge.org/core/journals/epidemiology-and-infection/article/influence-of-weather-on-community-gastroenteritis-in-australia/4FE582A7F05BB12F442BB4C98834E0ED
147. Lam LT. The association between climatic factors and childhood illnesses presented to hospital emergency among young children. International Journal of Environmental Health Research. 2007;17(1):1-8. Available from: https://doi.org/10.1080/09603120601124264
148. Schlenker W, Roberts MJ. Nonlinear temperature effects indicate severe damages to U.S. crop yields under climate change. Proceedings of the National Academy of Sciences. 2009;106(37):15594. Available from: http://www.pnas.org/content/106/37/15594.abstract
149. Crimmins A, J. Balbus, J.L. Gamble, C.B. Beard, J.E. Bell, D. Dodgen, et al. The Impacts of Climate Change on Human Health in the United States: A Scientiffic Assessment. GlobalChange.gov: U.S Global Change Research Program (USGCRP); 2016. Available from: http://dx.doi.org/10.7930/J0R49N
150. Jofre J, Blanch A, Lucena F. Water-Borne Infectious Disease Outbreaks Associated with Water Scarcity and Rainfall Events. 82009. p. 147-59.
151. Wu X, Lu Y, Zhou S, Chen L, Xu B. Impact of climate change on human infectious diseases: Empirical evidence and human adaptation. Environment International. 2016;86:14-23. Available from: http://www.sciencedirect.com/science/article/pii/S0160412015300489
152. Smith MR, Myers SS. Impact of anthropogenic CO2 emissions on global human nutrition. Nature Climate Change. 2018;8(9):834-9. Available from: https://doi.org/10.1038/s41558-018-0253-3
153. Davis J. Graincorp, Australia's largest grain handler records $59 million half-year loss due to drought Online: ABC News; 2019 [Available from: https://www.abc.net.au/news/rural/2019-05-09/graincorp-takes-59-million-hit-due-to-drought/11096030
154. Boffetta P, Couto E, Wichmann J, Ferrari P, Trichopoulos D, Bueno-de-Mesquita HB, et al. Fruit and Vegetable Intake and Overall Cancer Risk in the European Prospective Investigation Into Cancer and Nutrition (EPIC). Journal of the National Cancer Institute. 2010;102:529-37. Available from: https://academic.oup.com/jnci/article/102/8/529/2516544
155. Colditz GA, Wolin KY, Gehlert S. Applying what we know to accelerate cancer prevention. Sci Transl Med. 2012;4(127):127rv4-rv4. Available from: https://www.ncbi.nlm.nih.gov/pmc/articles/PMC3343638/
156. Martin-Moreno JM, Soerjomataram I, Magnusson G. Cancer causes and prevention: A condensed appraisal in Europe in 2008. European Journal of Cancer. 2008;44(10):1390-403. Available from: http://www.sciencedirect.com/science/article/pii/S0959804908000907
157. Stewart BW. Priorities for cancer prevention: lifestyle choices versus unavoidable exposures. The Lancet Oncology. 2012;13(3):e126-e33. Available from: http://www.sciencedirect.com/science/article/pii/S1470204511702212
158. Gregory PJ, Ingram JS, Brklacich M. Climate change and food security. Philosophical Transactions of the Royal Society B: Biological Sciences. 2005;360(1463):2139-48. Available from: http://dx.doi.org/10.1098/rstb.2005.1745
159. Delcour I, Spanoghe P, Uyttendaele M. Literature review: Impact of climate change on pesticide use. Food Research International. 2015;68:7-15. Available from: http://dx.doi.org/10.1016/j.foodres.2014.09.030
160. Ziska L, Crimmins A, Auclair A, DeGrasse S, Garofalo JF, Khan AS, et al. Ch. 7: Food Safety, Nutrition, and Distribution. The Impacts of Climate Change on Human Health in the United States: A Scientific Assessment. Washington, DC: U.S. Global Change Research Program; 2016. p. 189–216.
161. Liu Y, Wu F. Global burden of aflatoxin-induced hepatocellular carcinoma: a risk assessment. Environmental health perspectives. 2010;118(6):818-24. Available from: https://pubmed.ncbi.nlm.nih.gov/20172840
162. Marx MA, Rodriguez CV, Greenko J, Das D, Heffernan R, Karpati AM, et al. Diarrheal illness detected through syndromic surveillance after a massive power outage: New York City, August 2003. American Journal of Public Health. 2006;96(3):547-53. Available from: http://dx.doi.org/10.2105/ajph.2004.061358
163. Conroy JP. Influence of elevated atmospheric CO2 concentrations on plant nutrition. Australian Journal of Botany. 1992;40(5):445-56. Available from: http://dx.doi.org/10.1071/bt9920445
164. Myers SS, Zanobetti A, Kloog I, Huybers P, Leakey AD, Bloom AJ, et al. Increasing CO2 threatens human nutrition. Nature. 2014;510(7503):139-42. Available from: http://dx.doi.org/10.1038/nature13179
165. Mozaffarian D, Hao T, Rimm EB, Willett WC, Hu FB. Changes in Diet and Lifestyle and Long-Term Weight Gain in Women and Men. New England Journal of Medicine. 2011;364(25):2392-404. Available from: https://doi.org/10.1056/NEJMoa1014296
166. Raubenheimer D, Machovsky-Capuska GE, Gosby AK, Simpson S. Nutritional ecology of obesity: from humans to companion animals. British Journal of Nutrition. 2015;113(S1):S26-S39. Available from: http://dx.doi.org/10.1017/s0007114514002323
167. Worm B, Barbier EB, Beaumont N, Duffy JE, Folke C, Halpern BS, et al. Impacts of Biodiversity Loss on Ocean Ecosystem Services. Science. 2006;314(5800):787. Available from: http://science.sciencemag.org/content/314/5800/787.abstract
168. Layman DK, Boileau RA, Erickson DJ, Painter JE, Shiue H, Sather C, et al. A reduced ratio of dietary carbohydrate to protein improves body composition and blood lipid profiles during weight loss in adult women. The Journal of nutrition. 2003;133(2):411-7. Available from: http://jn.nutrition.org/content/133/2/411.full.pdf+html
169. Myers SS, Zanobetti A, Kloog I, Huybers P, Leakey ADB, Bloom AJ, et al. Increasing CO2 threatens human nutrition. Nature. 2014;510(7503):139-42. Available from: https://doi.org/10.1038/nature13179
170. Wong CS, Mooney JC, Brandt JR, Staples AO, Jelacic S, Boster DR, et al. Risk factors for the hemolytic uremic syndrome in children infected with Escherichia coli O157: H7: a multivariable analysis. Clinical Infectious Diseases. 2012;55(1):33-41. Available from: http://dx.doi.org/10.1093/cid/cis299
171. Eskenazi B, Rosas LG, Marks AR, Bradman A, Harley K, Holland N, et al. Pesticide toxicity and the developing brain. Basic & clinical pharmacology & toxicology. 2008;102(2):228-36. Available from: http://dx.doi.org/10.1111/j.1742-7843.2007.00171.x
172. Furey S, Strugnell C, McIlveen MH. An investigation of the potential existence of``food deserts''in rural and urban areas of Northern Ireland. Agriculture and Human Values. 2001;18(4):447-57. Available from: http://dx.doi.org/10.1023/a:1015218502547
173. Hendrickson D, Smith C, Eikenberry N. Fruit and vegetable access in four low-income food deserts communities in Minnesota. Agriculture and Human Values. 2006;23(3):371-83. Available from: http://dx.doi.org/10.1007/s10460-006-9002-8
174. Green D, Minchin L. Living on Climate-Changed Country: Indigenous Health, Well-Being and Climate Change in Remote Australian Communities. EcoHealth. 2014;11(2):263-72.
175. RACP. Climate change contributes to growth in mosquito populations [press release]: Royal Australian College of Physicians; 2016 [Available from: https://www.racp.edu.au/docs/default-source/default-document-library/media-releases/media-release---climate-change-contributes-to-growth-in-mosquito-populations.pdf?sfvrsn=0
176. Watts N, Amann M, Arnell N, Ayeb-Karlsson S, Belesova K, Boykoff M, et al. The 2019 report of The Lancet Countdown on health and climate change: ensuring that the health of a child born today is not defined by a changing climate. The Lancet. 2019;394(10211):1836-78. Available from: https://doi.org/10.1016/S0140-6736(19)32596-6
177. Berry H, Kelly B, Hanigan I, Coates J, McMichael A, Welsh J, et al. Rural mental health impacts of climate change. 2008. Available from: http://www.garnautreview.org.au/CA25734E0016A131/WebObj/03-DMentalhealth/$File/03-D%20Mental%20health.pdf
178. CSIRO, Bureau of Meterology. Climate Change in Australia Information for Australia's Natural Resource Management Regions: Technical Report. CSIRO and Bureau of Meterology, Australia; 2015. Available from: https://www.climatechangeinaustralia.gov.au/en/publications-library/technical-report/
179. Padhy SK, Sarkar S, Panigrahi M, Paul S. Mental health effects of climate change. Indian J Occup Environ Med. 2015;19(1):3-7. Available from: https://www.ncbi.nlm.nih.gov/pmc/articles/PMC4446935/
180. Neria Y, Shultz JM. Mental health effects of Hurricane Sandy: characteristics, potential aftermath, and response. JAMA. 2012;308(24):2571-2. Available from: https://www.ncbi.nlm.nih.gov/pmc/articles/PMC3638199/
181. Scurfield R, Halpern J, Tramontin M. Book Review: Disaster mental health: Theory and practice. Traumatology. 2007;13(2):37-8. Available from: https://journals.sagepub.com/doi/abs/10.1177/1534765607302930
182. Shukla J. Extreme Weather Events and Mental Health: Tackling the Psychosocial Challenge. ISRN Public Health. 2013. Available from: http://dx.doi.org/10.1155/2013/127365
183. Goldmann E, Galea S. Mental Health Consequences of Disasters. Annual Review of Public Health. 2014;35(1):169-83. Available from: https://doi.org/10.1146/annurev-publhealth-032013-182435
184. Forbes D, Alkemade N, Waters E, Gibbs L, Gallagher C, Pattison P, et al. The role of anger and ongoing stressors in mental health following a natural disaster. Australian & New Zealand Journal of Psychiatry. 2015;49(8):706-13. Available from: https://doi.org/10.1177/0004867414565478
185. Trombley J, Chalupka S, Anderko L. Climate Change and Mental Health: An evidence-based review of the emotional health risks associated with a changing climate. AJN. 2017;117(4):44-52. Available from: https://qualaxia.org/wp-content/uploads/trombley-2017-climate-change-and-mental-health.pdf
186. Fernandez A, Black J, Jones M, Wilson L, Salvador-Carulla L, Astell-Burt T, et al. Flooding and mental health: a systematic mapping review. PloS one. 2015;10(4):e0119929. Available from: https://doi.org/10.1371/journal.pone.0119929
187. Steffen W, Hughes L, Perkins S. Heatwaves: Hotter, longer, more often Climate Council of Australia 2014. Available from: https://www.climatecouncil.org.au/resources/heatwaves-report
188. Zhang Y, Beggs PJ. The Lancet Countdown down under: tracking progress on health and climate change in Australia. Med J Aust. 2018(208):285-6.
189. Hayes K, Blashki G, Wiseman J, Burke S, Reifels L. Climate change and mental health: Risks, impacts and priority actions. International journal of mental health systems. 2018;12(1):28. Available from: https://ijmhs.biomedcentral.com/articles/10.1186/s13033-018-0210-6
190. Coêlho AEL, Adair JG, Mocellin JSP. Psychological responses to drought in northeastern Brazil. Revista Interamericana de Psicologia/Interamerican Journal of Psychology. 2004;38(1). Available from: https://doi.org/10.30849/rip/ijp.v38i1.845
191. Hanigan IC, Butler CD, Kokic PN, Hutchinson MF. Suicide and drought in new South Wales, Australia, 1970–2007. Proceedings of the National Academy of Sciences. 2012;109(35):13950-5. Available from: https://doi.org/10.1073/pnas.1112965109
192. Hanna EG, McIver LJ. Climate change: a brief overview of the science and health impacts for Australia. Medical Journal of Australia. 2018;208(7):311-5. Available from: https://doi.org/10.5694/mja17.00640
193. Christopher P Burgess, Fay H Johnston, Helen L Berry, Joseph McDonnell, Dean Yibarbuk, Charlie Gunabarra, et al. Healthy country, healthy people: the relationship between Indigenous health status and "caring for country". MJA. 2009;190(10):567-72. Available from: https://doi.org/10.5694/j.1326-5377.2009.tb02566.x

194. Calma T, Dick D. Social determinants and the health of Indigenous peoples in Australia—A human rights based approach International Symposium on the Social Determinants of Indigenous Health. Adelaide2007 [updated 2007. Available from: https://www.humanrights.gov.au/about/news/speeches/social-determinants-and-health-indigenous-peoples-australia-human-rights-based
195. Warsini S, Mills J, Usher K. Solastalgia: living with the environmental damage caused by natural disasters. Prehospital and disaster medicine. 2014;29(1):87-90. Available from: https://doi.org/10.1017/S1049023X13009266
196. Fritze JG, Blashki GA, Burke S, Wiseman J. Hope, despair and transformation: climate change and the promotion of mental health and wellbeing. International journal of mental health systems. 2008;2(1):13. Available from: https://ijmhs.biomedcentral.com/articles/10.1186/1752-4458-2-13
197. Albrecht G, Sartore G-M, Connor L, Higginbotham N, Freeman S, Kelly B, et al. Solastalgia: the distress caused by environmental change. Australasian psychiatry. 2007;15(sup1):S95-S8. Available from: https://www.tandfonline.com/doi/abs/10.1080/10398560701701288
198. Sun G, Qian S, Jiang Q, Liu K, Li B, Li M, et al. Hyperthermia-induced disruption of functional connectivity in the human brain network. PloS one. 2013;8(4). Available from: 10.1371/journal.pone.0061157
199. American Physiological Society. Stress hormones spike as the temperature rises: Study surprisingly finds higher cortisol levels in summer than in winter. 2018. Available from: www.sciencedaily.com/releases/2018/04/180425131906.htm
200. Okamoto-Mizuno K, Mizuno K. Effects of thermal environment on sleep and circadian rhythm. Journal of physiological anthropology. 2012;31(1):14. Available from: https://link.springer.com/article/10.1186/1880-6805-31-14
201. Simister J, Cooper C. Thermal stress in the USA: effects on violence and on employee behaviour. Stress and Health: Journal of the International Society for the Investigation of Stress. 2005;21(1):3-15. Available from: https://doi.org/10.1002/smi.1029
202. Nitschke M, Tucker GR, Bi P. Morbidity and mortality during heatwaves in metropolitan Adelaide. Medical journal of Australia. 2007;187(11-12):662-5. Available from: https://doi.org/10.5694/j.1326-5377.2007.tb01466.x
203. Hansen A, Bi P, Nitschke M, Ryan P, Pisaniello D, Tucker G. The effect of heat waves on mental health in a temperate Australian city. Environmental health perspectives. 2008;116(10):1369-75. Available from: https://doi.org/10.1289/ehp.11339
204. Burke M, González F, Baylis P, Heft-Neal S, Baysan C, Basu S, et al. Higher temperatures increase suicide rates in the United States and Mexico. Nature climate change. 2018;8(8):723-9. Available from: https://doi.org/10.1038/s41558-018-0222-x
205. Page LA, Hajat S, Kovats RS. Relationship between daily suicide counts and temperature in England and Wales. The British Journal of Psychiatry. 2007;191(2):106-12. Available from: https://doi.org/10.1192/bjp.bp.106.031948
206. Department of Environment Land Water and Planning. Independent expert advice on interim targets Victorian Parliament: Victorian State Government; 2019 [Available from: https://www.climatechange.vic.gov.au/reducing-emissions/interim-targets
207. Stöllberger C, Lutz W, Finsterer J. Heat-related side-effects of neurological and non-neurological medication may increase heatwave fatalities. European journal of neurology. 2009;16(7):879-82. Available from: https://doi.org/10.1111/j.1468-1331.2009.02581.x
208. Lõhmus M. Possible biological mechanisms linking mental health and heat—a contemplative review. Int J Environ Res Public Health. 2018;15(7):1515. Available from: https://doi.org/10.3390/ijerph15071515
209. Vida S, Durocher M, Ouarda TBMJ, Gosselin P. Relationship between ambient temperature and humidity and visits to mental health emergency departments in Québec. Psychiatric Services. 2012;63(11):1150-3. Available from: https://doi.org/10.1176/appi.ps.201100485
210. Martin-Latry K, Goumy M-P, Latry P, Gabinski C, Bégaud B, Faure I, et al. Psychotropic drugs use and risk of heat-related hospitalisation. European Psychiatry. 2007;22(6):335-8. Available from: https://doi.org/10.1016/j.eurpsy.2007.03.007
211. Kilbourne EM, Choi K, Jones TS, Thacker SB. Risk factors for heatstroke: a case-control study. Jama. 1982;247(24):3332-6. Available from: https://jamanetwork.com/journals/jama/article-abstract/374849
212. Christenson ML, Geiger SD, Anderson HA. Heat-related fatalities in Wisconsin during the summer of 2012. WMJ. 2013;112(5):219-23. Available from: https://pubmed.ncbi.nlm.nih.gov/24734418/
213. Lian H, Ruan Y, Liang R, Liu X, Fan Z. Short-term effect of ambient temperature and the risk of stroke: a systematic review and meta-analysis. Int J Environ Res Public Health. 2015;12(8):9068-88. Available from: https://doi.org/10.3390/ijerph120809068
214. Lavados PM, Olavarría VV, Hoffmeister L. Ambient temperature and stroke risk: evidence supporting a short-term effect at a population level from acute environmental exposures. Stroke. 2018;49(1):255-61. Available from: https://doi.org/10.1161/STROKEAHA.117.017838
215. Zhang Y, Li S, Pan X, Tong S, Jaakkola JJK, Gasparrini A, et al. The effects of ambient temperature on cerebrovascular mortality: an epidemiologic study in four climatic zones in China. Environmental Health. 2014;13(1):24. Available from: https://ehjournal.biomedcentral.com/articles/10.1186/1476-069X-13-24
216. Davis SL, Wilson TE, White AT, Frohman EM. Thermoregulation in multiple sclerosis. Journal of Applied Physiology. 2010;109(5):1531-7. Available from: https://pubmed.ncbi.nlm.nih.gov/20671034/
217. Marino FE. Heat reactions in multiple sclerosis: an overlooked paradigm in the study of comparative fatigue. International Journal of Hyperthermia. 2009;25(1):34-40. Available from: https://doi.org/10.1080/02656730802294020



218. Cordes-Holland O. The sinking of the Strait: the implications of climate change for Torres Strait Islanders' human rights protected by the ICCPR. Melb J Int'l L. 2008;9:405. Available from: https://heinonline.org/HOL/LandingPage?handle=hein.journals/meljil9&div=21&id=&page=
219. Norman C. Torres Strait islanders take legal action against Australian government over failure to act on climate change TVNZ. One News2019 [Available from: https://www.tvnz.co.nz/one-news/world/torres-strait-islanders-take-legal-action-against-australian-government-over-failure-act-climate-change
220. World Health Organization. Gender, Climate Change and Health. 2014. Available from: https://apps.who.int/iris/bitstream/handle/10665/144781/9789241508186_eng.pdf;sequence=1
221. Neumayer E, Plümper T. The gendered nature of natural disasters: The impact of catastrophic events on the gender gap in life expectancy, 1981–2002. Annals of the Association of American Geographers. 2007;97(3):551-66. Available from: https://doi.org/10.1111/j.1467-8306.2007.00563.x
222. International Federation of Red Cross, Red Crescent Societies. World Disasters Report: Focus On Discrimination. 2007. Available from: https://www.ifrc.org/PageFiles/99876/WDR2007-English.pdf
223. Norris FH, Friedman MJ, Watson PJ, Byrne CM, Diaz E, Kaniasty K. 60,000 disaster victims speak: Part I. An empirical review of the empirical literature, 1981–2001. Psychiatry: Interpersonal and biological processes. 2002;65(3):207-39. Available from: https://doi.org/10.1521/psyc.65.3.207.20173
224. Sorensen C, Murray V, Lemery J, Balbus J. Climate change and women's health: Impacts and policy directions. PLOS Medicine. 2018;15(7):e1002603. Available from: https://doi.org/10.1371/journal.pmed.1002603
225. United Nations Division for the Advancement of Women. Environmental Management and the Mitigation of Natural Disasters: a Gender Perspective. 2001. Available from: https://www.un.org/womenwatch/daw/csw/env_manage/documents/EGM-Turkey-final-report.pdf
226. Tong VT, Zotti ME, Hsia J. Impact of the Red River catastrophic flood on women giving birth in North Dakota, 1994–2000. Maternal and child health journal. 2011;15(3):281-8. Available from: https://link.springer.com/article/10.1007/s10995-010-0576-9
227. Rahman MS. Climate change, disaster and gender vulnerability: A study on two divisions of Bangladesh. American Journal of Human Ecology. 2013;2(2):72-82. Available from: https://doi.org/10.11634/216796221504315
228. Kuehn L, McCormick S. Heat exposure and maternal health in the face of climate change. Int J Environ Res Public Health. 2017;14(8):853. Available from: https://doi.org/10.3390/ijerph14080853
229. Makhseed Ma, Musini VM, Ahmed MA, Monem RA. Influence of seasonal variation on pregnancy-induced hypertension and/or preeclampsia. Australian and New Zealand journal of obstetrics and gynaecology. 1999;39(2):196-9. Available from: https://doi.org/10.1111/j.1479-828X.1999.tb03372.x
230. Van Zutphen AR, Lin S, Fletcher BA, Hwang S-A. A population-based case–control study of extreme summer temperature and birth defects. Environmental health perspectives. 2012;120(10):1443-9. Available from: https://doi.org/10.1289/ehp.1104671
231. Ha S, Liu D, Zhu Y, Soo Kim S, Sherman S, Grantz KL, et al. Ambient temperature and stillbirth: a multi-center retrospective cohort study. Environmental health perspectives. 2017;125(6):067011. Available from: https://doi.org/10.1289/EHP945
232. Glinianaia SV, Rankin J, Bell R, Pless-Mulloli T, Howel D. Particulate air pollution and fetal health: a systematic review of the epidemiologic evidence. Epidemiology. 2004;15(1):36-45. Available from: www.jstor.org/stable/20485838
233. Pope DP, Mishra V, Thompson L, Siddiqui AR, Rehfuess EA, Weber M, et al. Risk of low birth weight and stillbirth associated with indoor air pollution from solid fuel use in developing countries. Epidemiologic reviews. 2010;32(1):70-81. Available from: https://doi.org/10.1093/epirev/mxq005
234. Šrám RJ, Binková B, Dejmek J, Bobak M. Ambient air pollution and pregnancy outcomes: a review of the literature. Environmental health perspectives. 2005;113(4):375-82. Available from: https://doi.org/10.1289/ehp.6362
235. Wadhwa PD, Buss C, Entringer S, Swanson JM. Developmental origins of health and disease: brief history of the approach and current focus on epigenetic mechanisms.27:358-68. Available from: https://www.thieme-connect.com/products/ejournals/html/10.1055/s-0029-1237424
236. DeNicola NG, Bekkar B, Pacheco S, Basu R. A scoping review of climate-change related exposures on obstetrics outcomes [18G]. Obstetrics & Gynecology. 2019;133:78S. Available from: https://journals.lww.com/greenjournal/Abstract/2019/05001/A_Scoping_Review_of_Climate_Change_Related.270.aspx
237. Birch EL, Meleis A, Wachter S. The Urban Water Transition: Why We Must Address the New Reality of Urbanization, Women, Water, and Sanitation in Sustainable Development. wH2O: The Journal of Gender and Water. 2012;1(1):1. Available from: https://repository.upenn.edu/cgi/viewcontent.cgi?article=1002&context=wh2ojournal
238. Duncan K. Global climate change, air pollution, and women's health. WIT Transactions on Ecology and the Environment. 2006;99. Available from: https://www.witpress.com/elibrary/wit-transactions-on-ecology-and-the-environment/99/17218
239. Shiva V, Jalees K. Water & women: A report by research foundation for science, technology, and ecology for national commission for women. Navdanya/RFSTE; 2005. Available from: https://catalog.hathitrust.org/Record/005628031
240. Jáuregui-Lobera I. Iron deficiency and cognitive functions. Neuropsychiatric disease and treatment. 2014;10:2087. Available from: https://www.ncbi.nlm.nih.gov/pmc/articles/PMC4235202/
241. Food Agriculture Organization. The state of food insecurity in the world 2002. Food insecurity: When people must live with hunger and fear starvation. Food and Agriculture Organization of the United Nations; 2002. Available from: http://www.fao.org/3/y7352e/y7352e00.htm



266. Australian Bureau of Statistics. National health survey: first results, 2017-18. Cat. no. 4364.0.55.001 Canberra: ABS; 2018. Available from: https://www.abs.gov.au/ausstats/abs@.nsf/mf/4364.0.55.001
267. Australian Institute of Health and Welfare. Chronic Obstructive Pulmonary Disease (COPD). Cat. no. ACM 35. Canberra: AIHW; 2019. Available from: https://www.aihw.gov.au/reports/chronic-respiratory-conditions/copd/contents/copd
268. Australian Institute of Health and Welfare. People with disability in Australia. Cat. no. DIS 72. Canberra: AIHW; 2019. Available from: https://www.aihw.gov.au/reports/disability/people-with-disability-in-australia/personal-factors/prevalence-of-disability
269. Dementia Australia. Dementia prevalence data 2018-2058. Canberra: NATSEM, University of Canberra; 2018. Available from: https://www.dementia.org.au/sites/default/files/documents/2018-2058%20Prevalence%20CED_AUSTRALIA_alpha.pdf
270. Hayes AJ, Lung TWC, Bauman A, Howard K. Modelling obesity trends in Australia: unravelling the past and predicting the future. International journal of obesity. 2017;41(1):178-85.
271. Magliano DJ, Peeters A, Vos T, Sicree R, Shaw J, Sindall C, et al. Projecting the burden of diabetes in Australia–what is the size of the matter? Australian and New Zealand journal of public health. 2009;33(6):540-3.
272. Lenzen M, Malik A, Li M, Fry J, Weisz H, Pichler P-P, et al. The environmental footprint of health care: a global assessment. The Lancet Planetary Health. 2020;4(7):e271-e9.
273. Doctors for the Environment Australia. Net zero carbon emissions: responsibilities, pathways and opportunities for Australia's healthcare sector 2020 [Available from: https://www.dea.org.au/wp-content/uploads/2020/12/DEA-Net-Zero-report_v11.pdf
274. Australian Medical Association Limited, Doctors for the Environment Australia. Joint statement - medical professionals call for emissions reduction in healthcare 2021 [Available from: https://ama.com.au/sites/default/files/2021-03/170321%20-%20Joint%20statement%20-%20Medical%20Professionals%20call%20for%20emissions%20reduction%20in%20health%20care.pdf
275. National Health Service. Delivering a 'net zero' National Health Service webpage NHS; 2020 [cited 2020 03 October]. Available from: https://www.england.nhs.uk/greenernhs/wp-content/uploads/sites/51/2020/10/delivering-a-net-zero-national-health-service.pdf
276. Burch H, McGain F. Renewable energy use in Australian public hospitals. Med J Aust. 2021.
277. McGain F, Story D, Kayak E, Kashima Y, McAlister S. Workplace Sustainability: The "Cradle to Grave" View of What We Do. Anesthesia & Analgesia. 2012;114(5). Available from: https://journals.lww.com/anesthesia-analgesia/Fulltext/2012/05000/Workplace_Sustainability__The__Cradle_to_Grave_.33.aspx
278. Vergunst F, Berry HL, Rugkåsa J, Burns T, Molodynski A, Maughan DL. Applying the triple bottom line of sustainability to healthcare research—a feasibility study. International Journal for Quality in Health Care. 2020;32(1):48-53.
279. Block DJ. Health Care Sustainability: Managing Natural Resources in Value-Based Care. Physician Leadership Journal. 2016;3(3):18-21. Available from: https://pubmed.ncbi.nlm.nih.gov/27295740/
280. Pencheon D, Rissel CE, Hadfield G, Madden DL. Health sector leadership in mitigating climate change: experience from the UK and NSW. New South Wales public health bulletin. 2010;20(12):173-6.
281. Xie E, Barros E, Abelsohn A, Stein A, Haines A. Challenges and opportunities in planetary health for primary care providers. The Lancet Planetary Health. 2018;2:e185-e7.
282. Campbell-Lendrum D. Taking the heat out of the population and climate debate. Bulletin of the World Health Organization. 2009;87:807.
283. Gipson J, Koenig M, Hindin M. The Effects of Unintended Pregnancy on Infant, Child, and Parental Health: A Review of the Literature. Studies in family planning. 2008;39:18-38.
284. Council of Presidents of Medical Colleges. Managing and responding to climate risks in healthcare. https://cpmc.edu.au/communique/managing-and-responding-to-climate-risks-in-healthcare: Deakin, ACT: CPMC; 2018.

### About Doctors for the Environment Australia

Doctors for the Environment Australia (DEA) is an independent, self-funded, non-governmental organisation of medical doctors and students in all Australian states and territories. We are supported by a distinguished Scientific Advisory Committee.

We work to address the public health impacts from damage to our natural environment such as climate change, which will increasingly undermine our health and our healthcare services if we fail to act.
A key focus of DEA's work is raising awareness of the healthcare sector's responsibility to reduce its sizeable carbon footprint and to ensuring adequate measures are instigated.

**DEA's Scientific Advisory Committee:**

Prof Stephen Boyden AM, Prof Emeritus Chris Burrell AO, Prof Colin Butler, Prof Peter Doherty AC, Prof Michael Kidd AM, Prof David de Kretser AC, Prof Stephen Leeder AO, Prof Ian Lowe AO, Prof Robyn McDermott, Prof Lidia Morawska, Prof Peter Newman AO, Prof Emeritus Sir Gustav Nossal AC, Prof Hugh Possingham, Prof Lawrie Powell AC, Prof Fiona Stanley AC, Dr Rosemary Stanton OAM, Dr Norman Swan

Suite 3 Ground Floor
60 Leicester Street, Carlton VIC 3053 0422 974 857
ABN 80 178 870 373

www.dea.org.au





**We acknowledge the Traditional Custodians of the many Lands we call Australia, and pay respects to Elders Past and Present, as well as emerging Aboriginal and Torres Strait Islander leaders.**

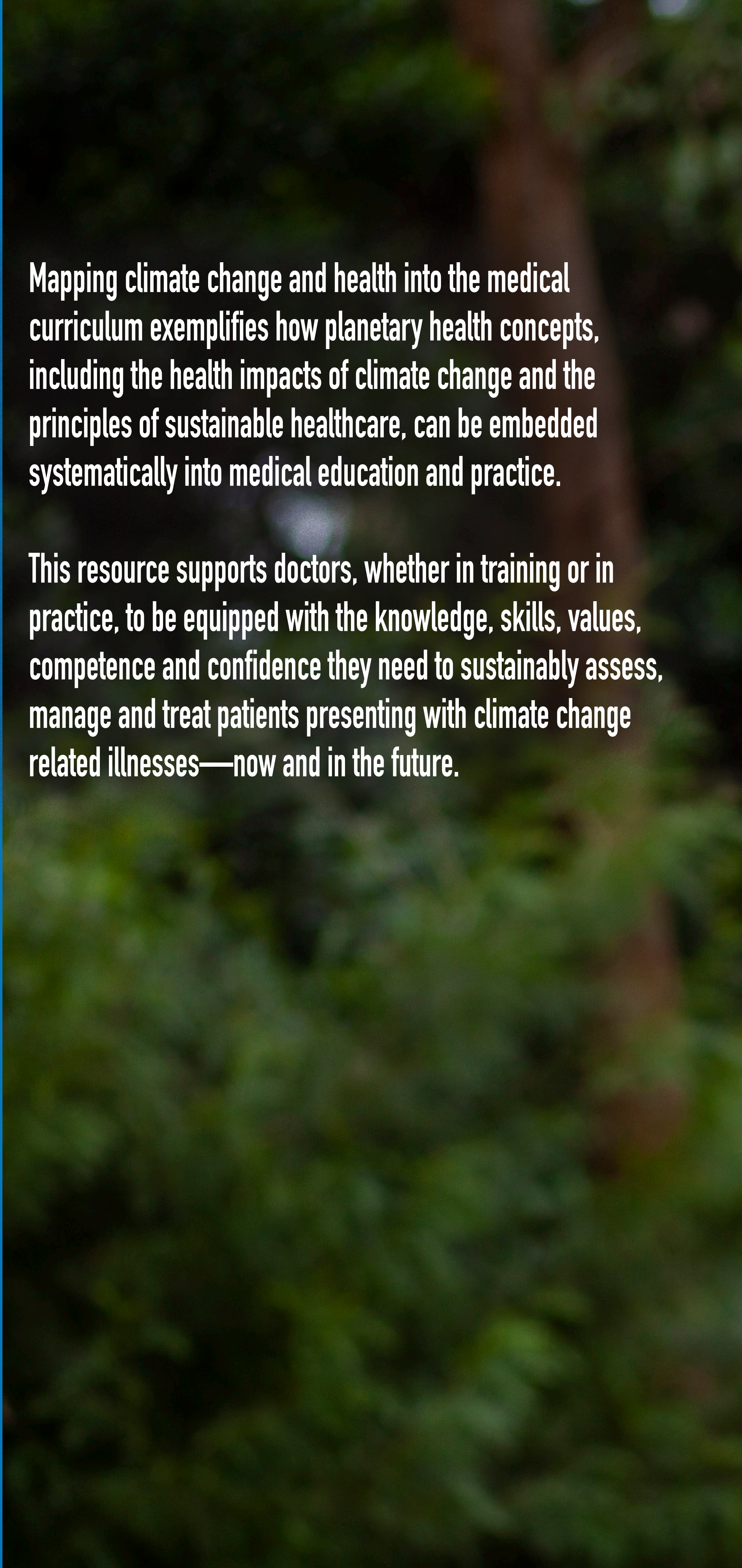

Mapping climate change and health into the medical curriculum exemplifies how planetary health concepts, including the health impacts of climate change and the principles of sustainable healthcare, can be embedded systematically into medical education and practice.

This resource supports doctors, whether in training or in practice, to be equipped with the knowledge, skills, values, competence and confidence they need to sustainably assess, manage and treat patients presenting with climate change related illnesses—now and in the future.

**Doctors for the Environment Australia (DEA) is an independent, self-funded, non-governmental organisation of medical doctors and students in all Australian states and territories. We are supported by a distinguished Scientific Advisory Committee.**

**We work to address the public health impacts from damage to our natural environment such as climate change, which will increasingly undermine our health and our healthcare services if we fail to act.**

**A key focus of DEA's work is raising awareness of the healthcare sector's responsibility to reduce its sizeable carbon footprint and to ensuring adequate measures are instigated.**

Cover image by Jesse Thompson

Doctors for the Environment Australia
